# Clinical Study to Evaluate the Possible Efficacy and Safety of Antibodies Combination (casirivimab and imdevimab) versus standard antiviral therapy as antiviral agent against Corona virus 2 infection in hospitalized COVID-19 patients

**DOI:** 10.1101/2022.08.20.22279020

**Authors:** Ahmed H. Hassan, Sahar K. Hegazy, Samar T. Radwan

## Abstract

**Introduction:** Corona Virus-induced disease – 2019 (COVID-19) pandemic stimulates research works to find a solution to this crisis from starting 2020 year up to now. With ending of the 2021 year, various advances in pharmacotherapy against COVID-19 have emerged.

Regarding antiviral therapy, Casirivimab and imdevimab antibody combination is a type of new immunotherapy against COVID-19. Standard antiviral therapy against COVID-19 includes Remdesivir and Favipravir.

**Aim of Study:** To compare the efficacy and safety of antibodies cocktail (casirivimab and imdevimab), Remdesivir, and Favipravir in the COVID-19 patients

**Patients and Population:** 265 hospitalized COVID-19 patients were used to represent the COVID-19 population and were assigned into three groups in a ratio of (1:2:2) respectively, Group (A) received REGN3048-3051(Antibodies cocktail (casirivimab and imdevimab), group (B) received remdesivir, and group (C) received favipravir.

**Methods:** The study design is a single-blind non-Randomized Controlled Trial (non-RCT). The drugs of the study are owned by Mansoura University Hospital (MUH) and prescribed by chest diseases lectures of the faculty of medicine-Mansoura University. The duration of the study is about 6 months after ethical approval.

**Results and discussion:** Casirivimab and imdevimab achieve less 28-day mortality rate, less mortality at hospital discharge, more negative swab cases, less need for O2 therapy and IMV, less duration of this need, less hospital and ICU stay, less case progression as presented by lower World Health Organization (WHO) scale and better multi-organ functions as presented by lower Sequential Organ Function Assessment (SOFA) score than Remdesivir and Favipravir.

**Conclusion:** From all of these results, it is concluded that Group A (Casirivimab & imdevimab) has more favorable clinical outcomes than B (remdesivir) & C (favipravir) intervention groups.

Clinical Trial Registration: NCT05502081, 16/08/2022, Clinicaltrials.gov, retrospectively registered

## I. Introduction

### 1.1. COVID-19 overview and classification

COVID-19 is an infectious viral disease caused by sever acute respiratory syndrome-corona virus 2 (SARS CoV-2) that has affected large number of people all over the world with high mortality rate (Okonji et al., 2021). COVID-19 infection has been classified(NIH, 2021) as: as Mild Illness in which signs and symptoms of COVID-19 exist (e.g., cough, fever, sore throat, headache, loss of taste and smell muscle pain, vomiting, diarrhea) but without dyspnea, shortness of breath, or abnormal chest imaging. Moderate Illness in which evidence of lower respiratory disease exists with an oxygen saturation (SpO2) ≥94% on room air. Severe Illness in which SpO2 <94% on room air, respiratory frequency >30 breaths/min, a ratio of arterial partial pressure of oxygen to fraction of inspired oxygen (PaO2/FiO2) <300 mm Hg, or lung infiltrates >50%. Critical Illness in which respiratory failure, septic shock, and/or multiple organ dysfunctions may occur.

COVID-19 pandemic stimulates research works to find a solution to this crisis from the start of the 2020-year up to now. With the end of the year 2021, various advances in pharmacotherapy against COVID-19 have emerged. (Umakanthan et al., 2021).

### 1.2. Standard and controversial antivirals used in treatment of COVID-19 (Remdesivir and Favipravir)

Regarding antiviral drugs used in treatment of COVID-19, Remdesivir is a standard antiviral against COVID-19 and has been approved by Food and drug administration (FDA) for treatment of mild, moderate, sever and critical hospitalized COVID-19 patients (Aleem and Kothadia, 2021). Other drugs have shown controversial antiviral activity include: favipravir, ivermectin, nitazoxanide, hydroxychloroquine, ribavirin. Favipravir became a standard antiviral which has been used for treatment of mild and moderate COVID-19 outpatients (de Almeida et al., 2020).

### 1.3. Advances in immunotherapy for treatment of COVID-19

Recently with the end of 2020, immunotherapy to target virus antigen has developed (Owji et al., 2020).Figure 1 shows two types of immunotherapies include active and passive immunotherapy. Active immunotherapy is to enhance body to produce antibodies against virus as by vaccination. Passive immunotherapy involves administration of products containing antibodies like plasma, or direct administration of prepared antibodies acting specifically against virus (Owji et al., 2020).

**Fig. 1.**
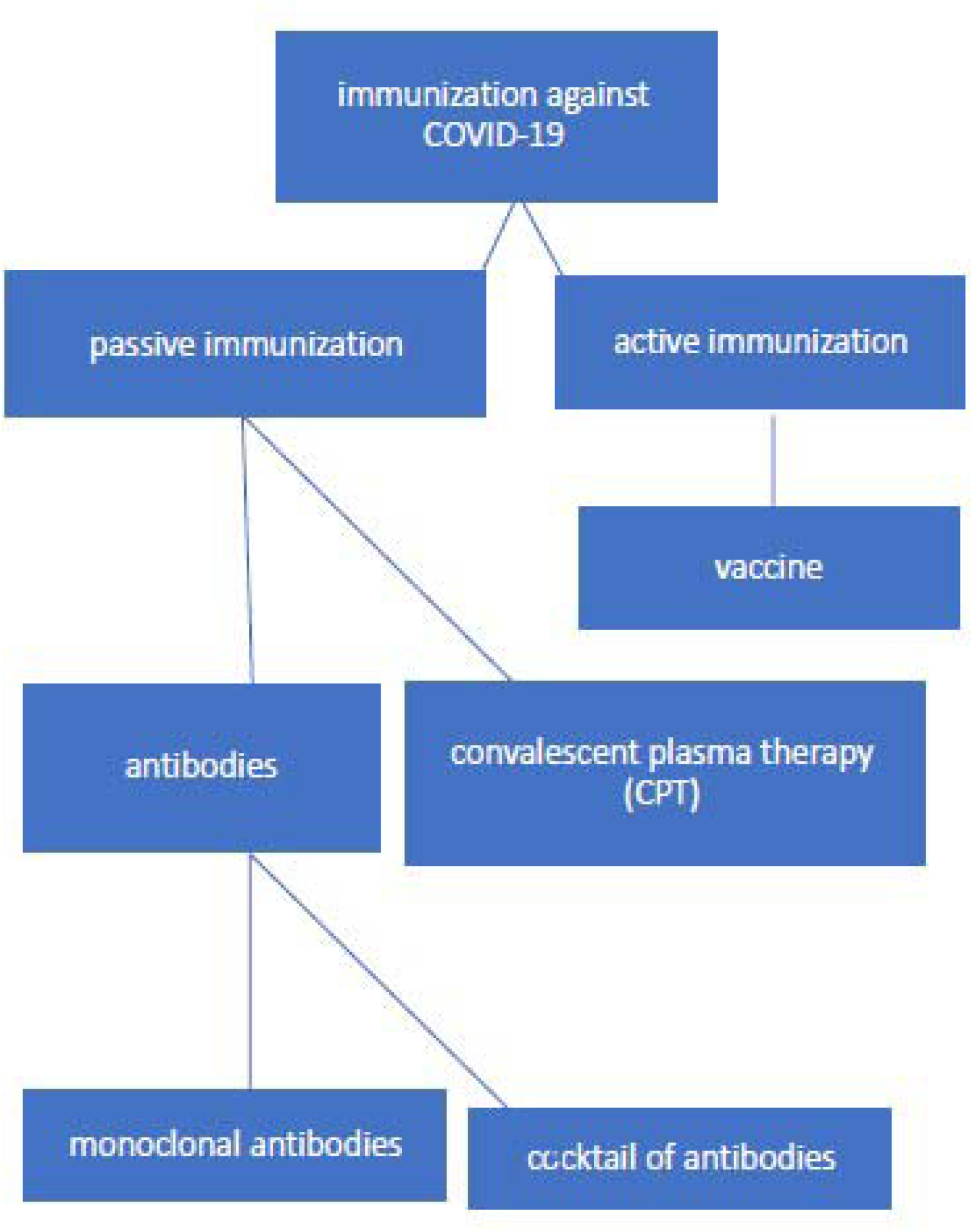
Immunization approaches against COVID-19. (Owji et al., 2020)

In this study, the point of research is antibodies cocktail including REGN3048-3051(casirivimab and imdevimab). There are three targets for these antibodies to work as antiviral including: antibodies that prevent the virus attachment and entry, antibodies that inhibit the virus replication and transcription, and antibodies that hinder various steps of the immune system response

Table 1 includes various types of antibodies under investigation for treatment of COVID-19 and their targets (Owji et al., 2020).

**Table 1.** shows antibodies candidate against SARS-CoV-2 under investigation by pharmaceutical companies. (Owji et al., 2020).

| **Antibody** | **Mechanism** | **Company** | **Stage of study/identification method** |
| --- | --- | --- | --- |
| VIR-7831/VIR-7832 | -Neutralize highly conserved epitope in S protein<br>-Induce NK-mediated antibody-dependent cell cytotoxicity | VIR biotechnology and GSK | Designed based on S309 (isolated from SARS-Cov patients) |
| SAB | Anti SARS-CoV-2 fully human polyclonal antibodies | SAB Biotherapeutics | -Antibodies produced in genetically engineered cattle will enter clinical trial by early summer<br>-SAB-301 against MERS passes phase 1 of clinical trial and entered phase II/III |
| — | Target multiple viral S epitope | ImmunoPrecise | Using B cell Select® and Deep Display® technology |
| COVID-HIG and COVID-EIG | Hyperimmune polyclonal antibody derived from human plasma or immunized horse | Emergent BioSolutions | Enter clinical trial within 4–5 months |
| Rcig | Recombinant anti SARS-CoV-2 hyperimmune gammaglobulin, polyclonal antibodies | GigaGen | Preclinical stage-<br>-Aimed for COVID19 hospitalized patients and prophylaxis in high risk individuals |
| Antibody cocktail including REGN3048-3051 | Fully human multivalent antibodies against the spike protein isolated from genetically modified mice or recovered COVID-19 patients | Regeneron | -Phase 1 clinical trial for Middle East Respiratory Syndrome (MERS) completed last year<br>-Clinical trial for SARS-CoV-2 starts by early summer |

**Table 2.** The three intervention groups of the study.

|  |  | Frequency | Percent | Valid Percent | Cumulative Percent |
| --- | --- | --- | --- | --- | --- |
| Valid | A | 53 | 20.0 | 20.0 | 20.0 |
|  | B | 106 | 40.0 | 40.0 | 60.0 |
|  | C | 106 | 40.0 | 40.0 | 100.0 |
|  | Total | 265 | 100.0 | 100.0 |  |

### 1.4. Casirivimab and Imdevimab as antibodies cocktail against COVID-19

Antibodies cocktail including REGN3048-3051(casirivimab and imdevimab) are human monoclonal antibodies that target the spike glycoprotein on surface of viral particles to prevent viral entry into human cells by the angiotensin-converting enzyme 2(ACE2) receptor (Baum et al., 2020, Hansen et al., 2020), and showed an antiviral activity and needs for further research to prove their efficacy in COVID patients.

## II. Aim of the study

To evaluate the efficacy of antibodies cocktail (casirivimab and imdevimab) compared to standard antiviral therapy in reducing 28-day mortality in hospitalized patients with moderate, severe or critical COVID19, as well to examine its safety by monitoring hypersensitivity and infusion related reactions or other significant adverse effects

## III. PATIENTS AND POPULATION

265hospitalized COVID-19 patients are used to represent COVID-19 population and was assigned into 3 groups in a ratio of (1:2:2) respectively, group (A) received REGN3048-3051(Antibodies cocktail (casirivimab and imdevimab)), group (B) received remdesivir, and group (C) received favipravir Populations in this study are the COVID-19 patients admitted to isolation hospital-Mansoura University. An electronic file containing a written informed consent from included patients will be provided. Paper will not be a tool for providing agreement by the patients or their relatives to avoid transmission of infection

Inclusion criteria include age more than 12 years old, weight not less than 40 kg, moderate, sever or critical COVID-19 disease as defined by WHO, and PCR-confirmed patients to be Positive before inclusion.

Exclusion criteria include history of hypersensitivity or infusion related reactions after administration of monoclonal antibodies, prior use of standard antiviral therapy (remdesivir or favipravir), Current use of controversial antiviral therapy (ivermectin, hydroxychloroquine, oseltamivir, nitazoxanide, ribavirin, lopinavir/ritonavir, daclatasvir, sofosbuvir, semipirvir, acyclovir, azithromycin), and patients expected to die within 48 hours.

## IV. INTERVENTIONS

Population included in this study will be assigned into 3 groups with 1:2:2 ratios to receive either antibodies cocktail or standard antiviral therapy (remdesvir, favipravir) as shown in table2 and figures 2,3.

**Figure 2.**
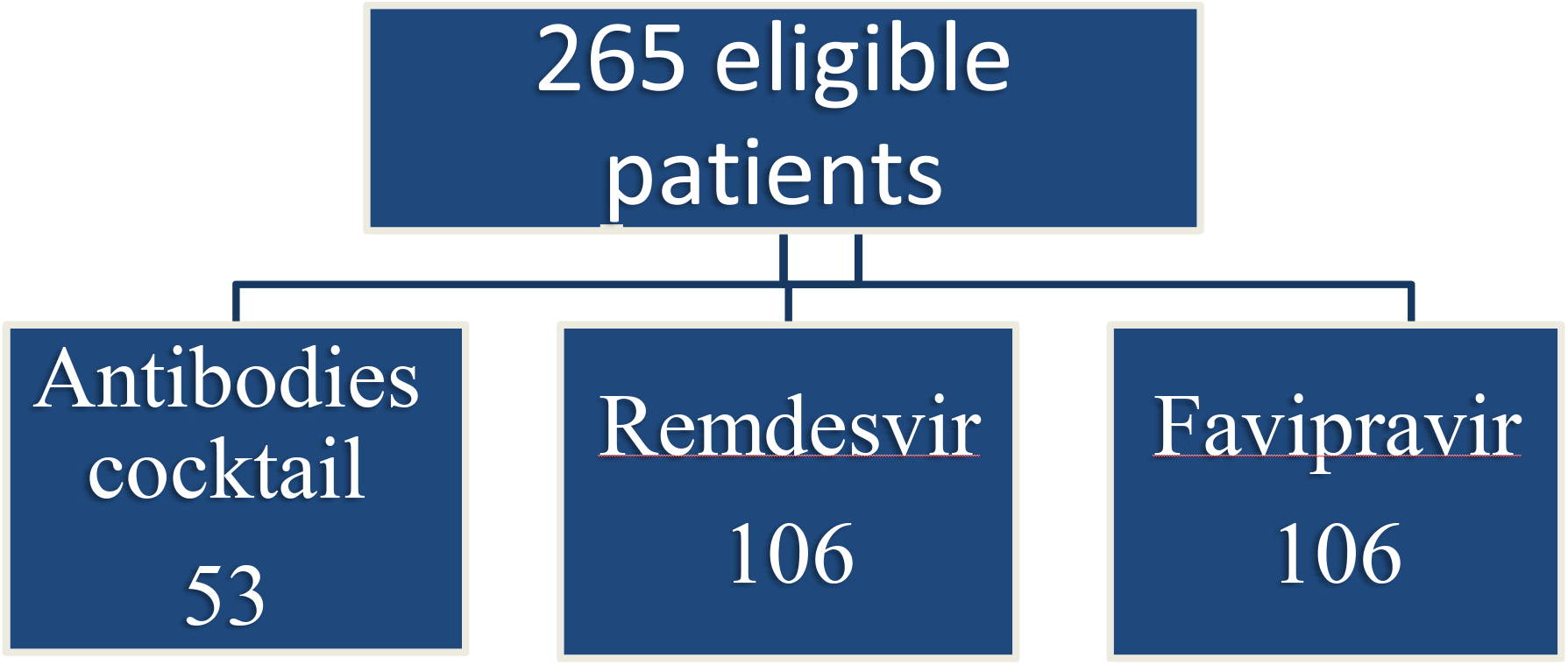
Assignment of the included COVID cases at their groups.

**Figure 3.**
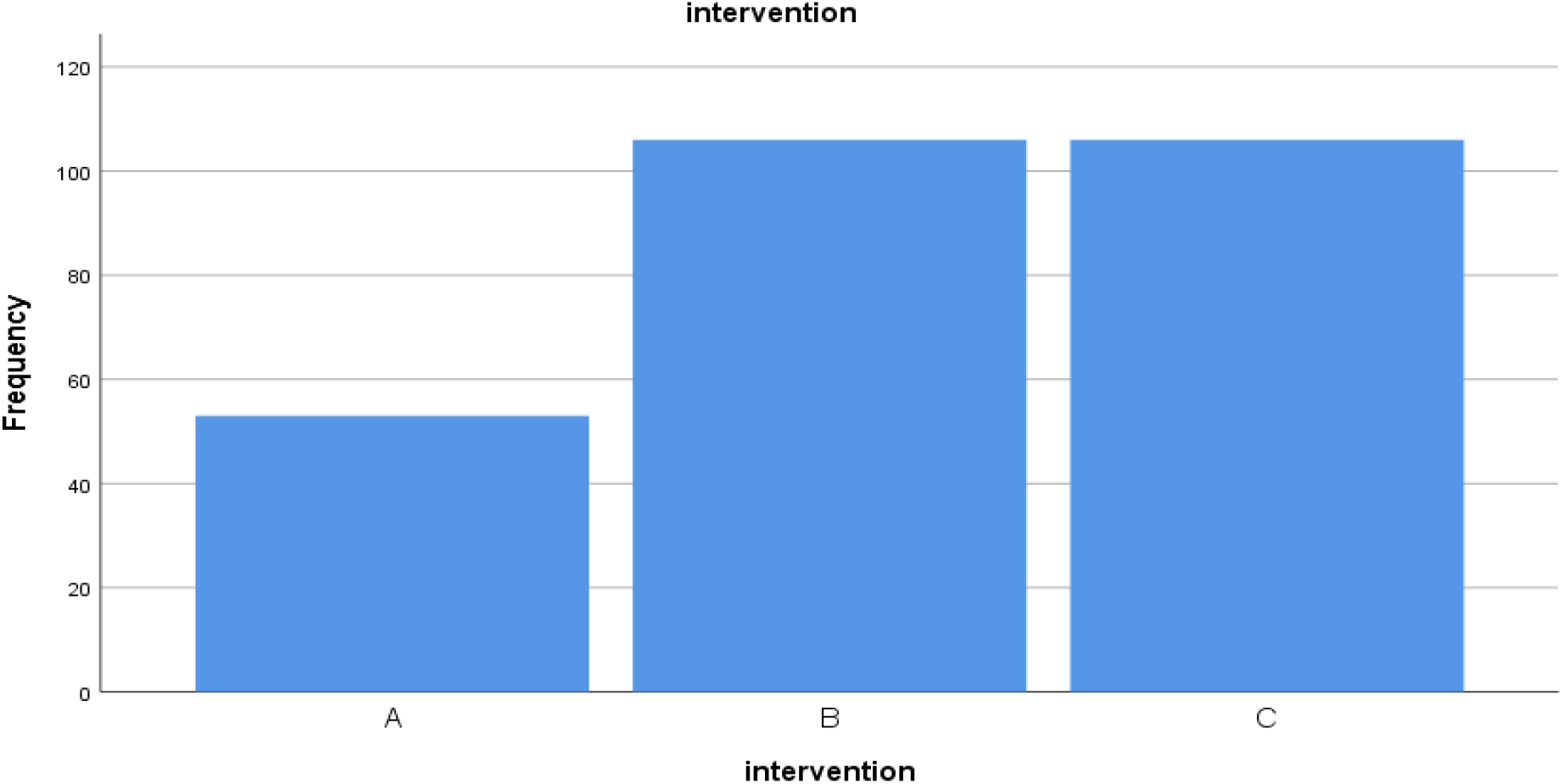
Frequency of interventions in included patients.

Group A patients will receive REGN3048-3051(Antibodies cocktail (casirivimab and imdevimab)) in low-dose regimen 1.2 gm (1200 mg of combined antibodies) diluted in 250 ml 0.9% sodium chloride solution as single I.V infusion over 30-60 minutes.

Group B patients will receive Remdesivir :

Day1 (loading dose): 200 mg (two 100mg vials) diluted in 500ml 0.9% sodium chloride solution infused I.V over 60 minutes

Day 2-5 or Day 2-10 (maintenance dose): 100 mg (one 100mg vial) in 250 ml 0.9% sodium chloride solution infused I.V over 30 minutes

Group C patients will receive Favipravir

Day 1 (loading dose): 1600 mg (8 tablets) or 1800 mg (9 tablets) orally or in Ryle tube / 12 hours

Day 2-5 or day 2-10 (maintenance dose): 600 mg (3 tablets) or 800 mg (4 tablets) orally or in Ryle tube / 12 hours

Patients will be received standard of care by Physicians, Clinical pharmacist, Nurses and as guided by Egyptian COVID-19 treatment protocol.

## V. METHOD

The type of this study is single blind non-RCT and is considered a Phase IV Clinical trial (post-marketing study) to report efficacy and safety of new medicine.

We use PubMed search tool to find clinical studies that performed to test efficacy and safety of developed immunotherapy in treatment of COVID-19 with about 4,000 results with focusing on antibodies developed as antiviral against COVID-19 obtaining only 70 results from which REGN-COV2, a Neutralizing Antibody Cocktail is selected with its only one clinical study up to now (REGN-COV2, a Neutralizing Antibody Cocktail, in Outpatients with Covid-19) which is published in New England Journal of Medicine on January 21, 2021.

Another resource used to obtain data is Fact Sheet for Health Care Providers-EUA OF casirivimab and imdevimab which provides clinical data about the use of this antibodies cocktail. Endnote citation software is used for citation of references.

## VI. Outcomes

Parameters that will be assessed during hospitalization at day 0(baseline), day 3, 7, 14, 28 include:

- C-reactive protein (CRP)
- ferritin
- lactate dehydrogenase (LDH)
- D-dimer
- serum creatinine (S.Cr) and estimated creatinine clearance (CrCl)
- alanine aminotransferase) (ALT), aspartate aminotransferase (AST), albumin, bilirubin

Clinical outcomes measured before & during intervention

### 1) Primary outcomes

1. 28-days mortality rate (efficacy).
2. PCR test results at end of hospital visit (efficacy).
3. precentage of patients who developed infusion related or hypersensitivity reactions during and after the end of drug infusion and reporting of any Serious and unexpected adverse events may occur that have not been previously reported with REGEN-COV use that may cause drug discontinuation (Safety).

### 2) Secondary outcomes

1. Need for invasive mechanical ventilation (IMV)
2. Invasive mechanical ventilation and oxygen support duration (days).
3. Time to clinical improvement(Garibaldi et al., 2021) (defined as 2 points reduction in the WHO disease ordinal progression scale(Plaçais et al., 2022) or discharge, whatever happens first)
4. ICU and hospitalization length of stay (days).
5. SOFA score(Yang et al., 2021) on day 0,3,7,14, and 28. ]
6. COVID19 WHO disease progression score(Plaçais et al., 2022) from day 0 to day 28.
7. Inflammatory markers including CRP, ferritin, LDH
8. liver and kidney functions

In addition to clinical outcomes measured before and during intervention, Vital signs, glasgow coma score (GCS),complete blood count (CBC), artieral blood gas (ABG) and prothrombin time (PT) is observed daily and recorded at day0.patients’ charactrestics(age,gender) and relevant medical and medication history and current COVID-19 treatment drugs will be recorded on admission

Duration of research will be about 6 months from Novmber 2021 to April 2022

## VII. Statistical analysis and Sample Size

### Statistical analysis

Intention-to-treat strategy will be used in this study. Statistical analysis will be achieved with SPSS, version 26.

Categorical variables will be presented as proportion and percent. Continuous variables will be presented as mean (standard deviation) for parametric data or as a median (25^th^-75^th^ percentile) for non-parametric data.

Regarding baseline characteristics, Kruskal-Wallis or ANOVA test (depending on type of data and the continous data distrubation (normal or not)) will be used to compare these characteristics between the study groups. We will report the P-value for our statistical tests with level of statistical significance will be P-value ≤ 0.05.

In case of existiing differences in some basline characteristics, logestic regression will be performed. This allows studying the effect of these variables on the primary outcomes of the study to exclude the effect of these confounding vaiables and to ensure the effect on the outcomes is due to interventions.

we will compare the 28-day all-, Regarding the outcomes result of PCR test at hospital discharge and, cause mortality rate incidence of infusion related or hypersensitivity reactions during and after the end of drug infusion (primary outcome) using the .Kruskal-Wallis test with reporting the P-value icu, While the secondary outcomes (hospital stay duration stay duration and others) are compared using Kruskal-Wallis or ANOVA test depending on type of data and the continous data distrubation (normal or not).

### Sample Size

A total sample sizes of 246 patients would achieve at least 80 % power to detect a risk difference of 0.2 (20%) in the 28-day all-cause mortality (primary outcome) with a significance level (α) of 0.05 and 95% confidence level using the ANOVA or Kruskal-Wallis test of independent proportion in G*Power software. To compensate for the estimated loss-to-follow-up and increase the study power, we will increase the sample size in both remdesivir and favipravir groups to be 106 patients compared to 53 patients in Antibodies cocktail Group As Antibodies cocktail product is available for only about 50 COVID-19 patients. In addition,the ratio (1:2:2) is the clostest to reality according to number of patients who recieve each drug.

The mortality data was estimated from the average mortality in Augest, Septmper, and Octobar 2021 at the Mansoura University Isolation Hospital among all hospitalized patients. Mortality rate is found to be about 360 cases in these 3 months (120 cases / month). The online system has been used to obtain mortality rate in these three monthes.

The current admission rate at the Mansoura University – Isolation Hospital is 250 cases per month on average; our needed sample is about 250 cases.

## RESULTS

After statstical analysis using SPSS software, all continous data shows no normal distrubation. So Kruskal-Wallis Test is used to compare non normally distrubated continous, catagorical and nominal variables between the three groups.

### 8.1. Regarding baseline characterestics

Table 3 shows the significance of difference between the three groups and also includes a pairwise comparison between every two groups in baseline characterestics if they show statistically significant difference between the three groups. Figures (4-23) show distrubuations and frequencies of baseline characterestics between the three groups.

**Table 3.**
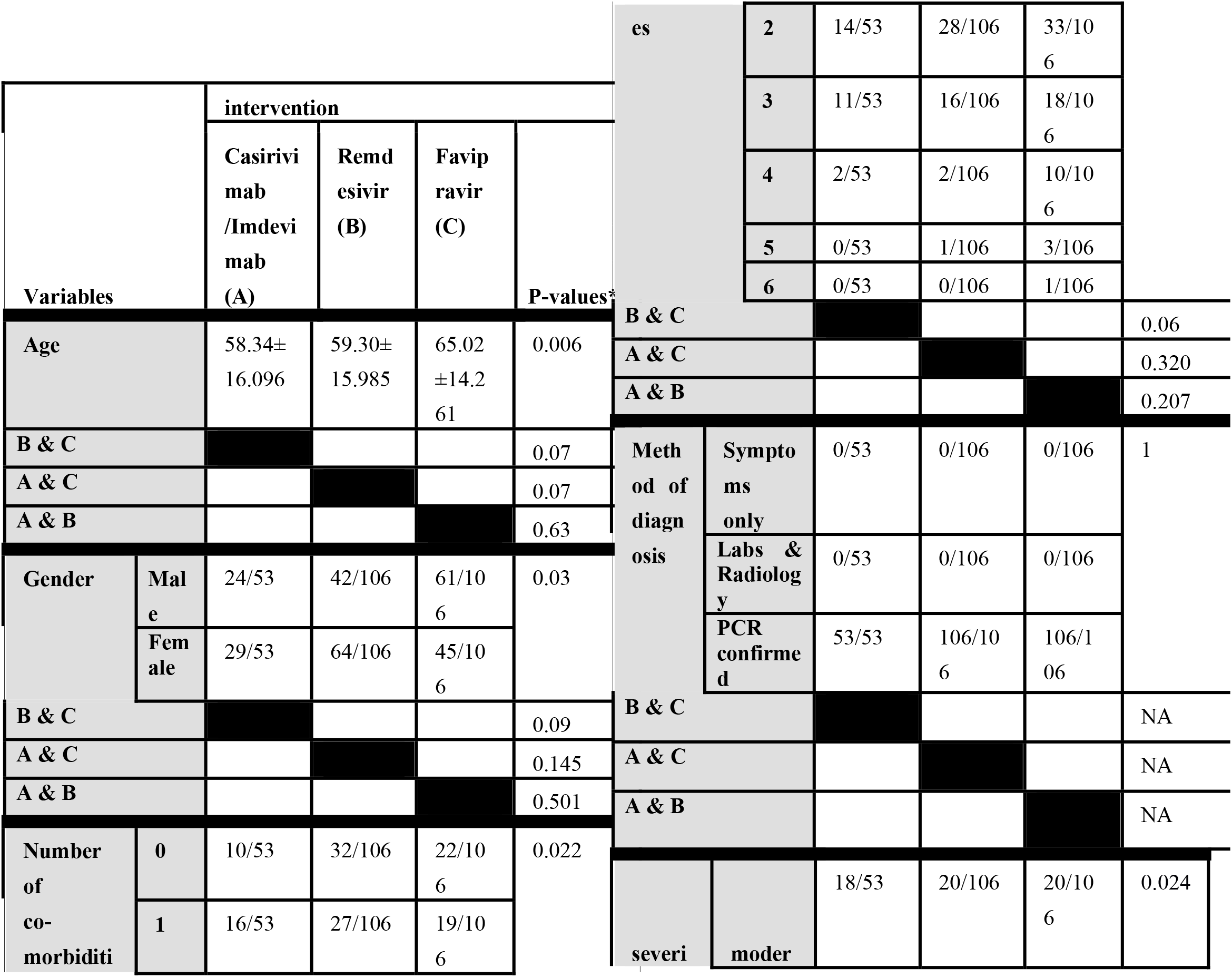
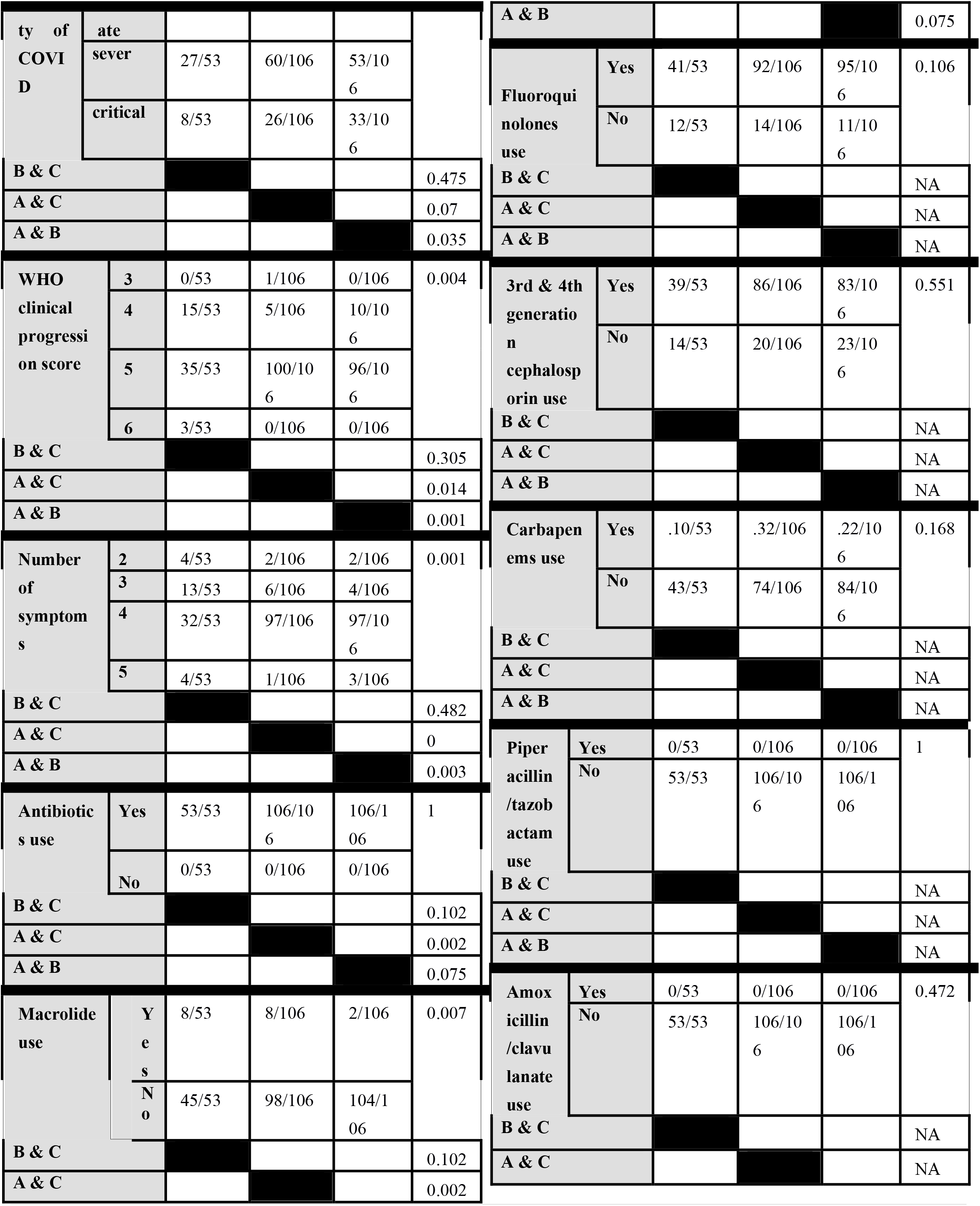
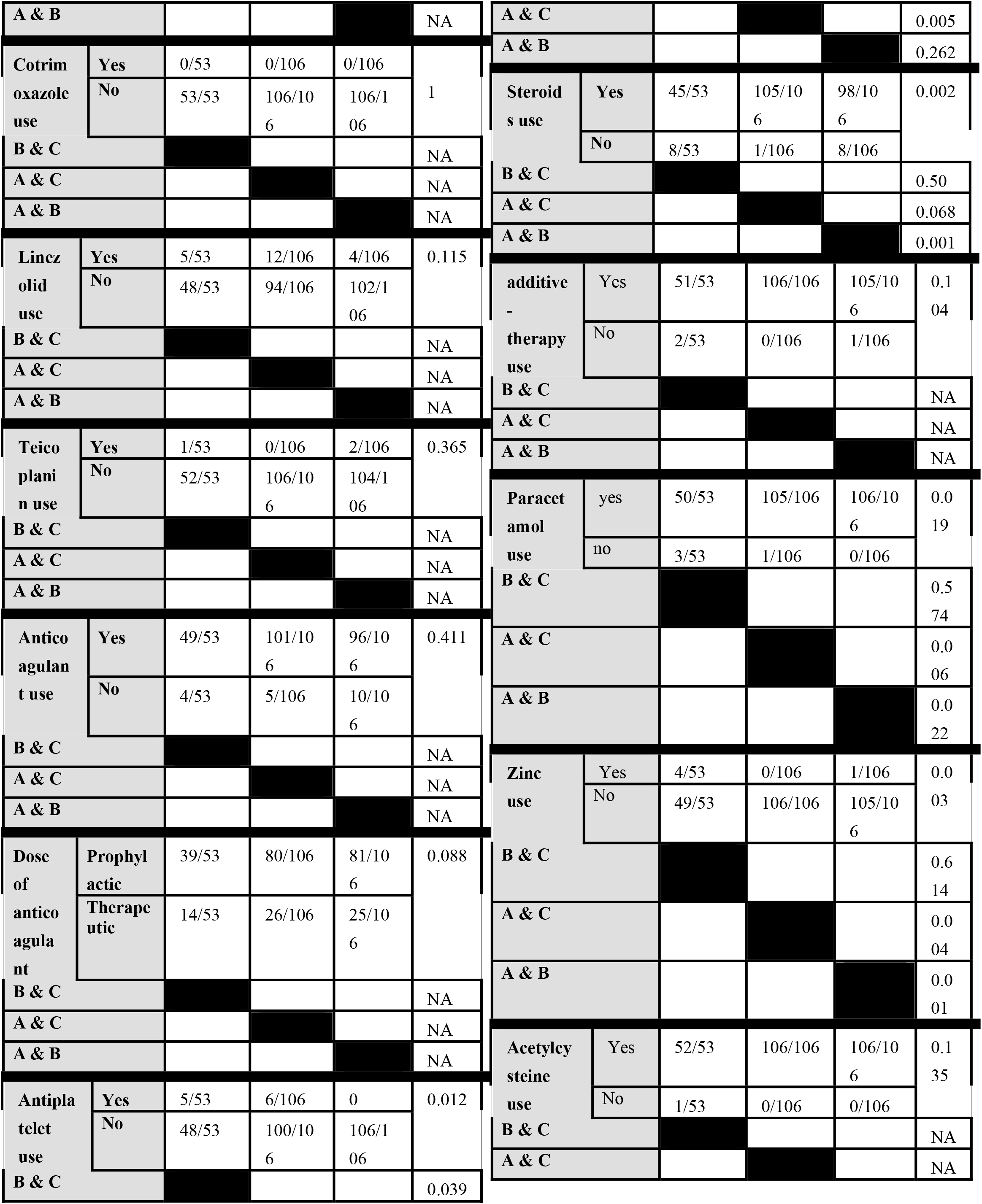
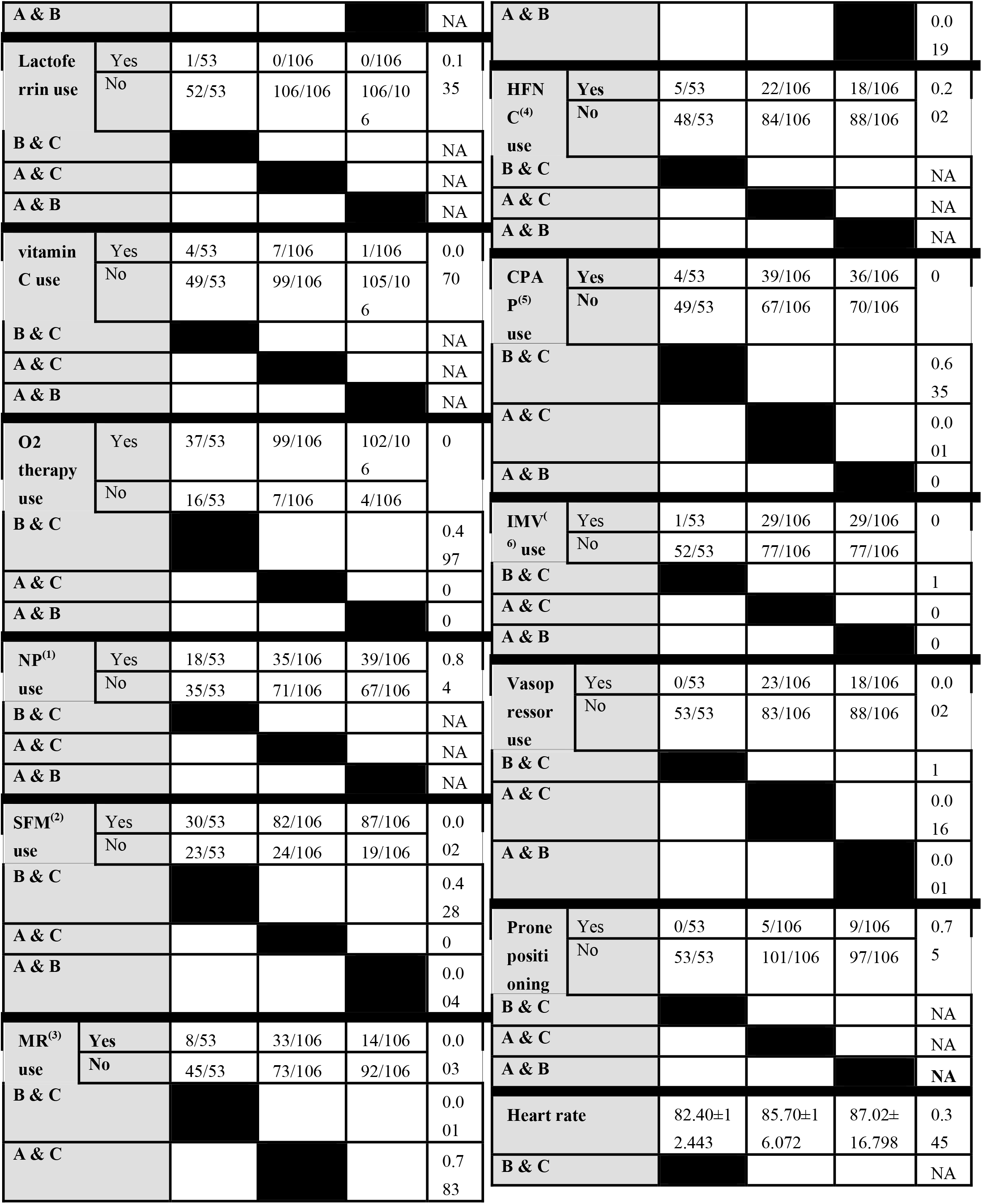
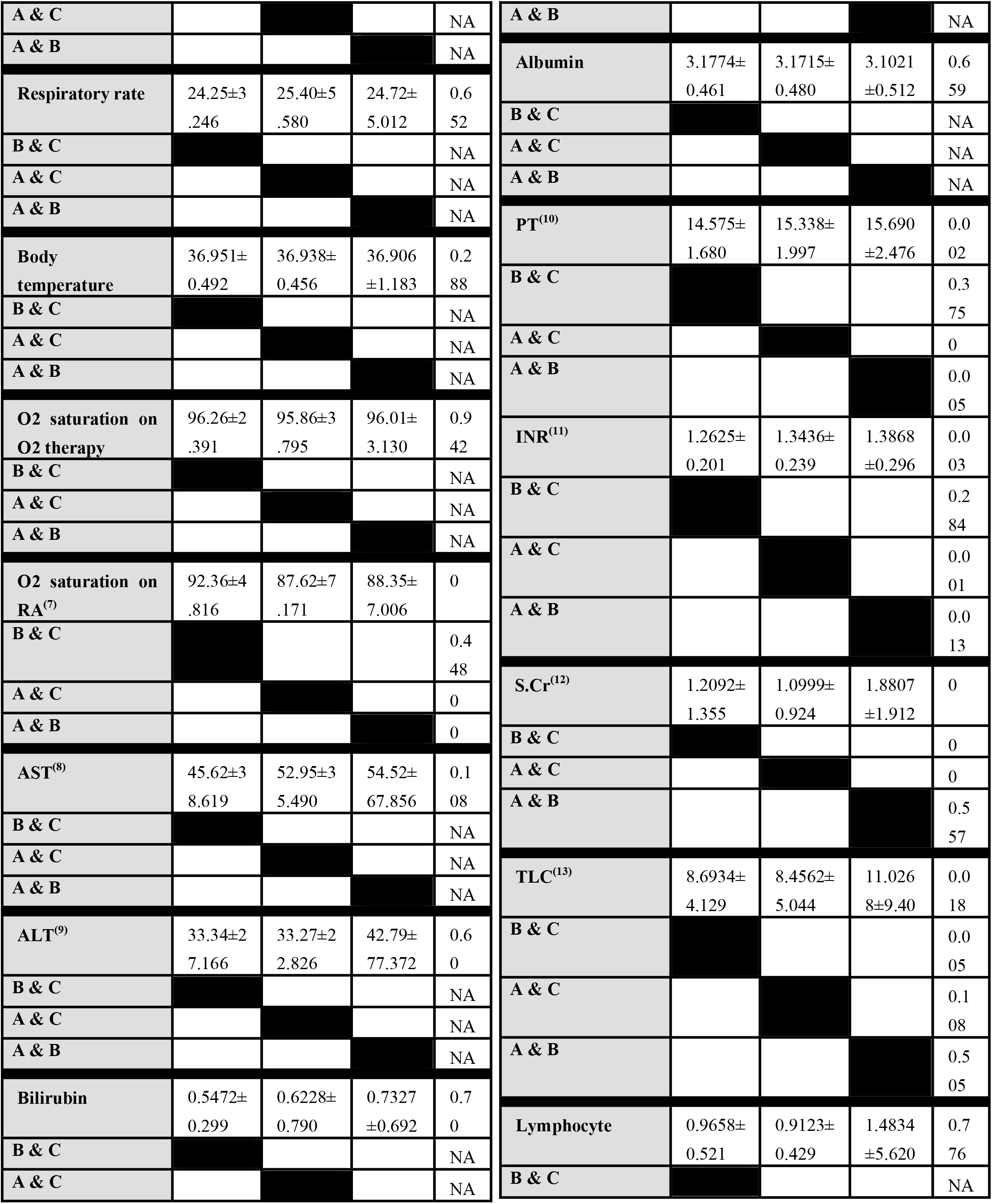
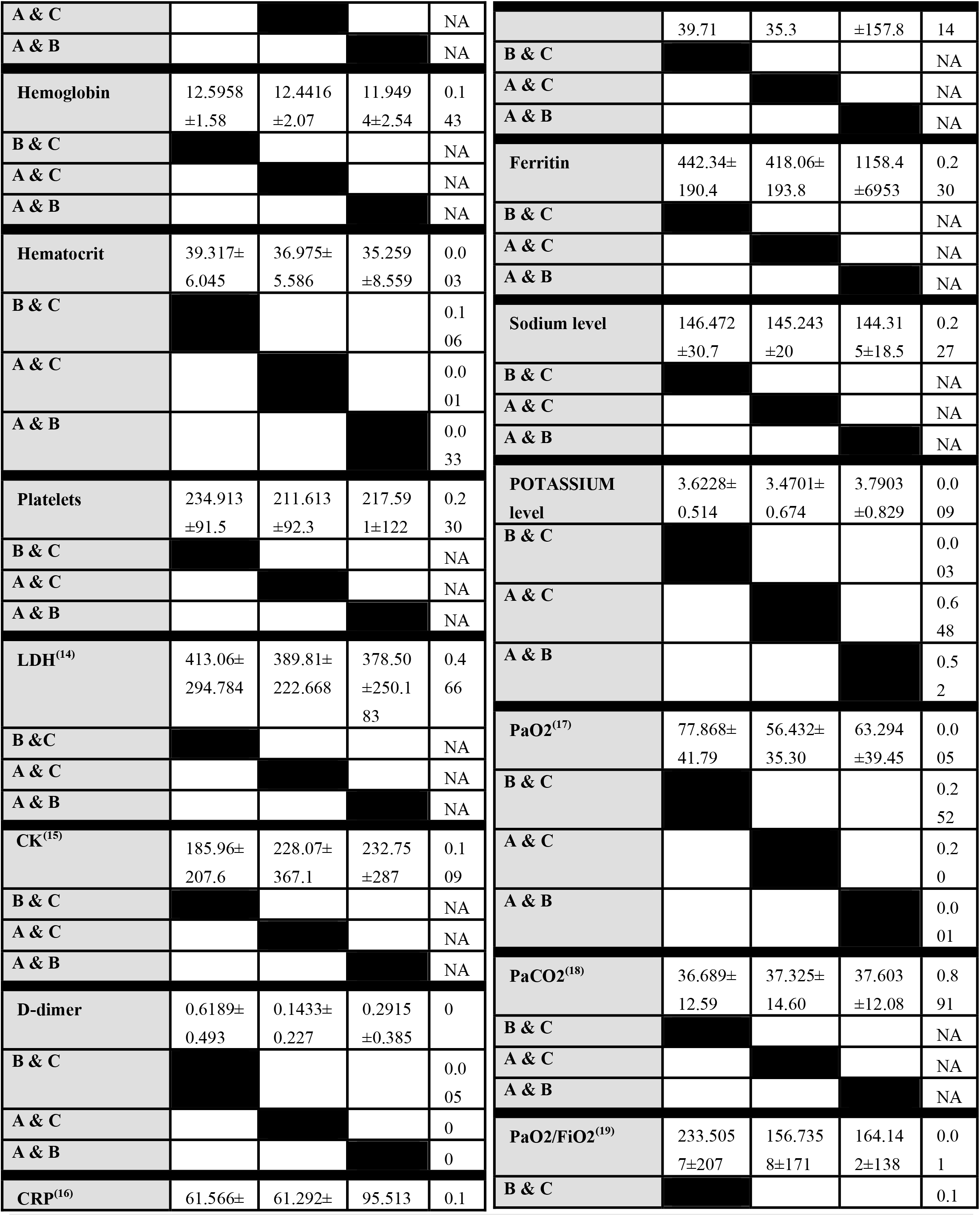
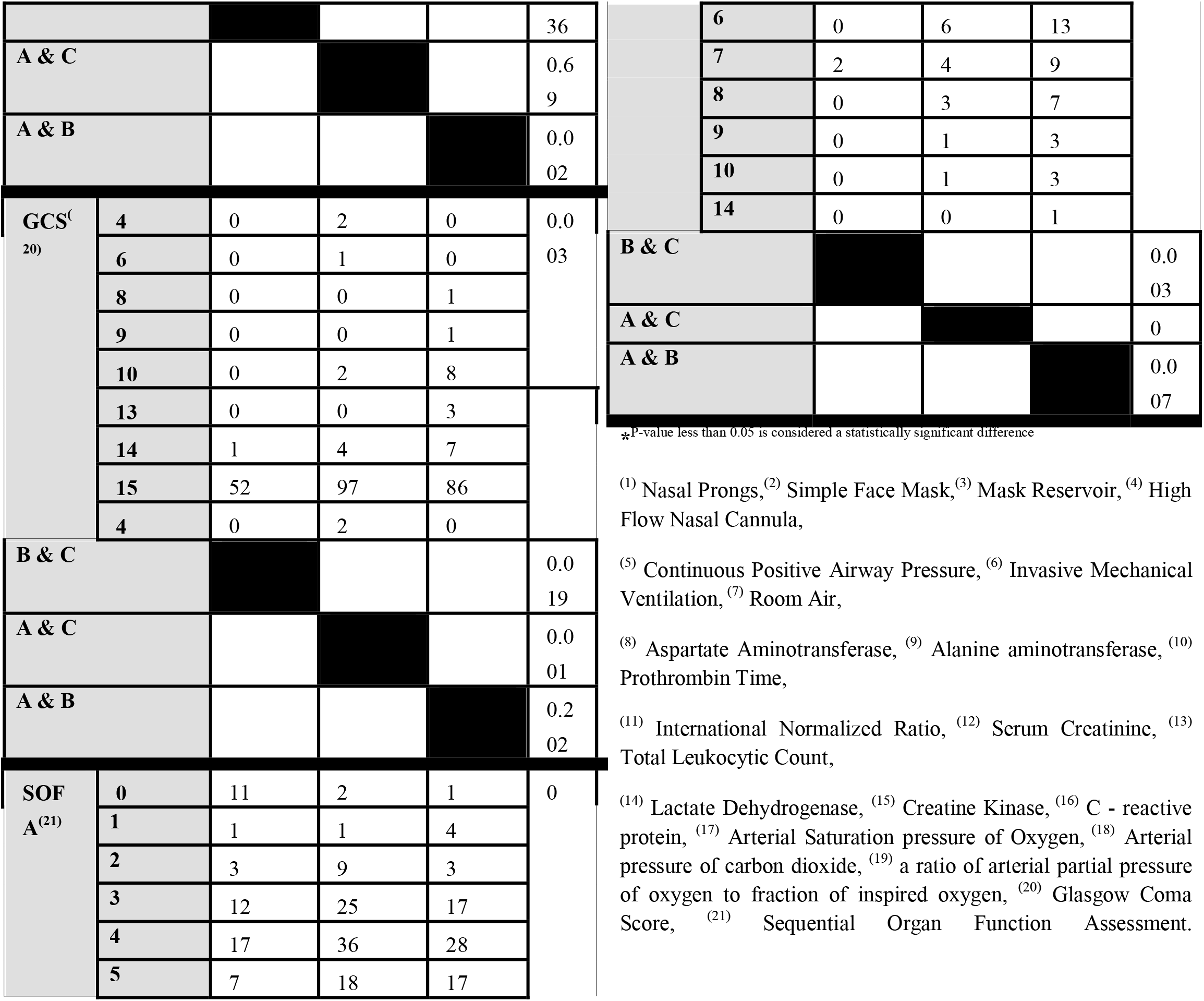
the Significance of differences in baseline characteristics between the three groups

**Figure 4.**
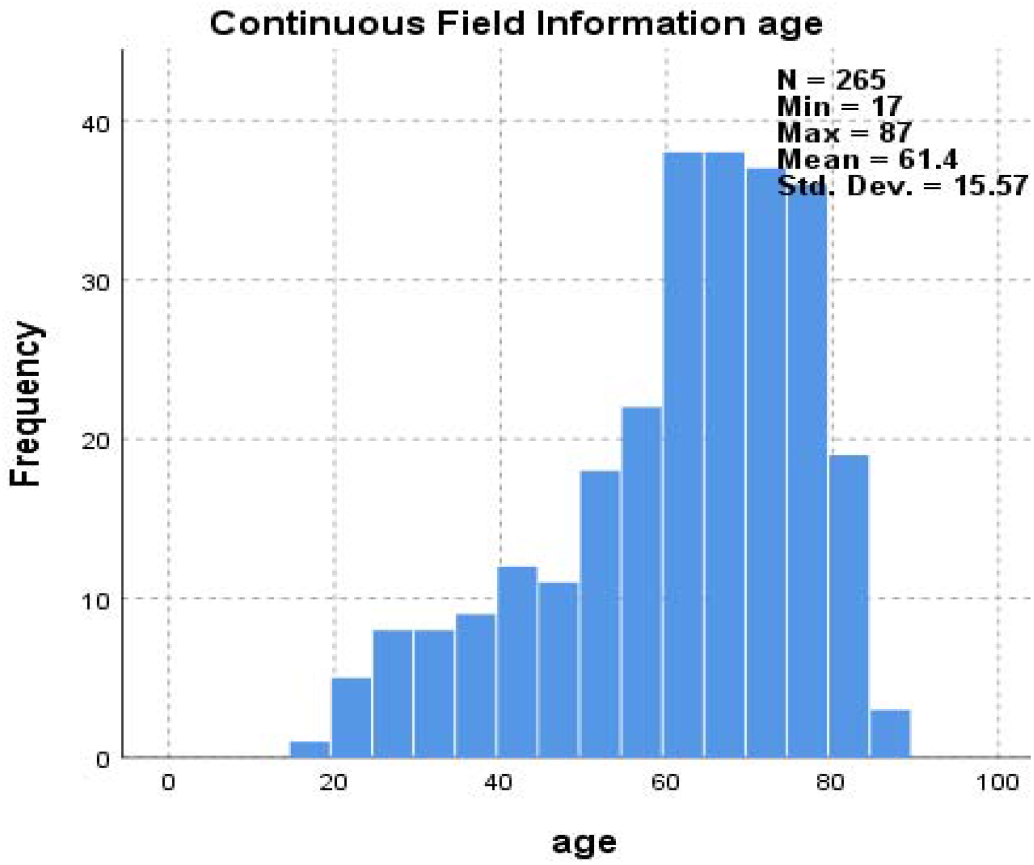
Frequency of Age in included patients.

**Figure 5.**
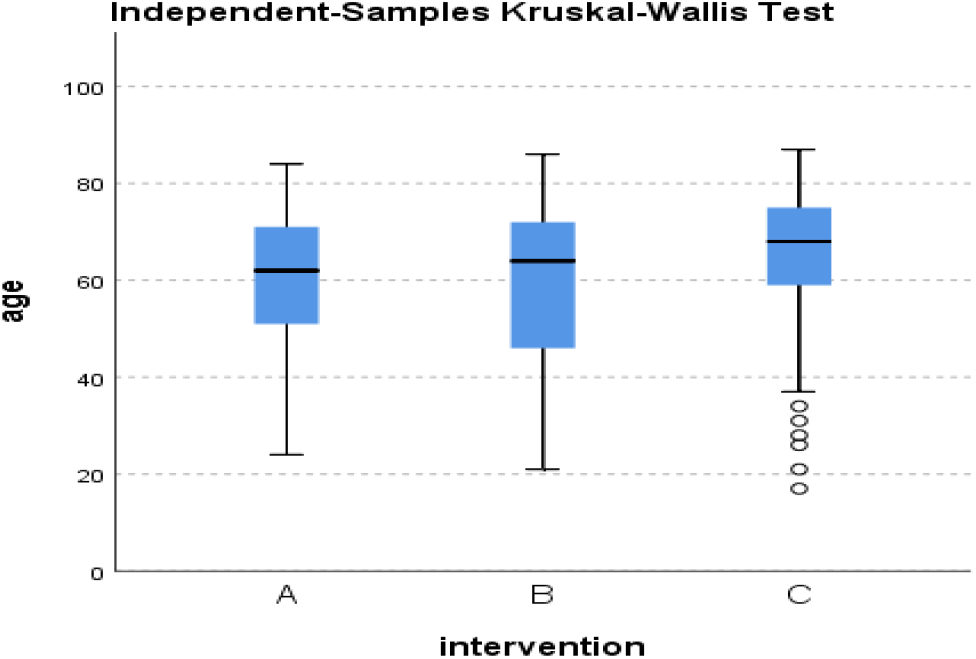
Distribution of Age across the three groups.

**Figure 6.**
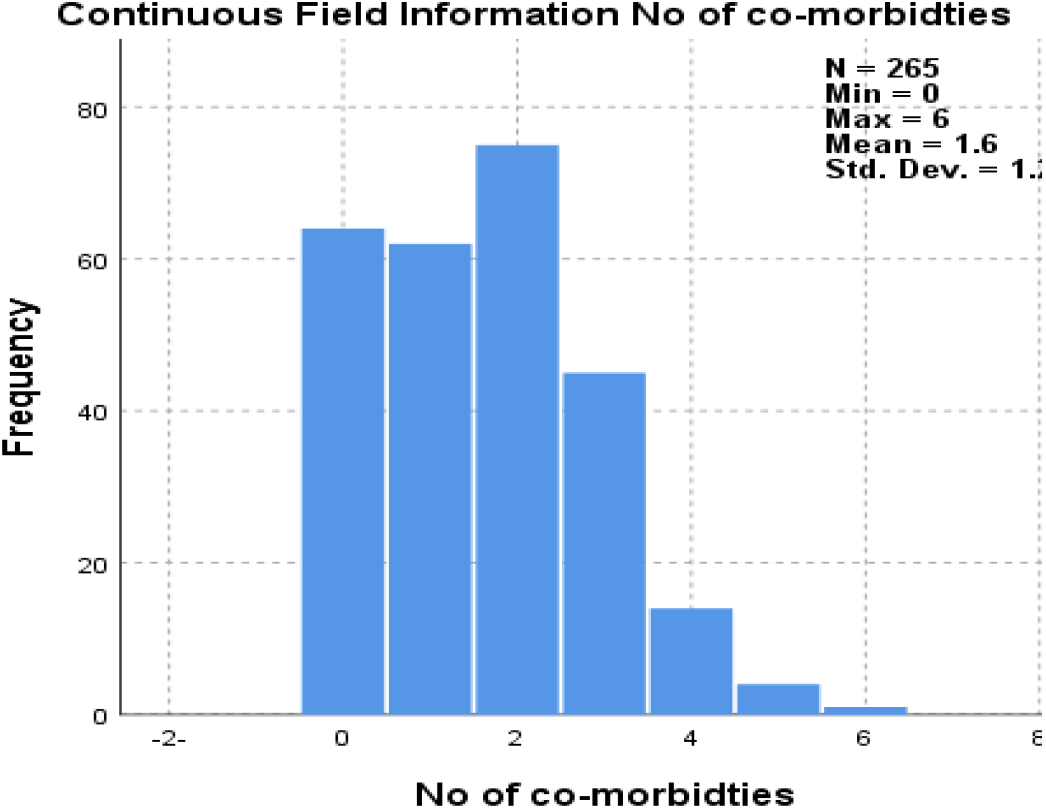
Frequency of co-morbidities number in included patients.

**Figure 7.**
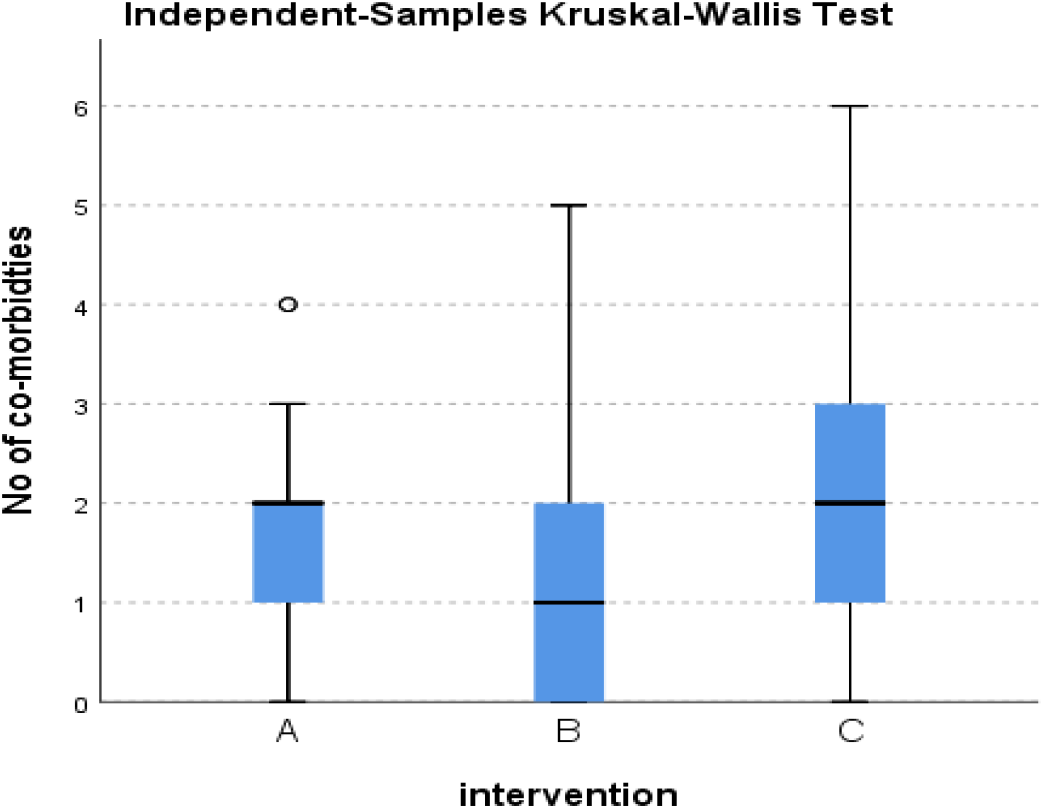
Distribution of comorbidities number across the three groups.

**Figure 8.**
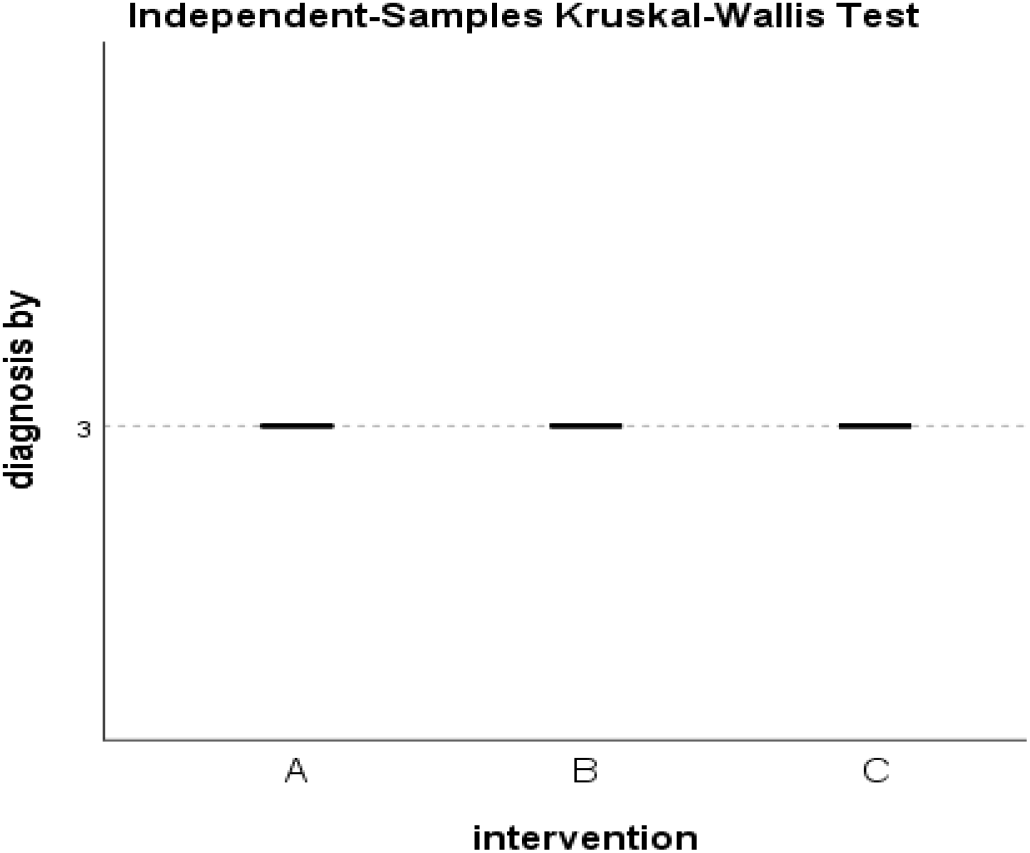
Distribution of Method of diagnosis across the three groups.

**Figure 9.**
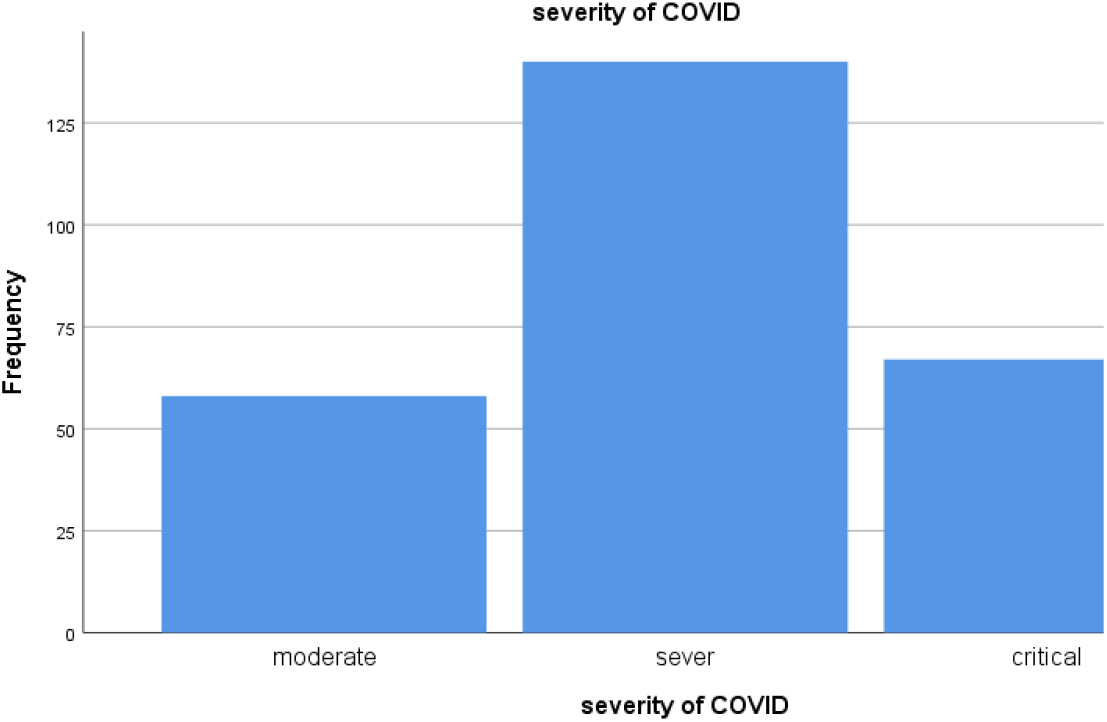
Frequency of COVID severity in included COVID-19 patients.

**Figure 10.**
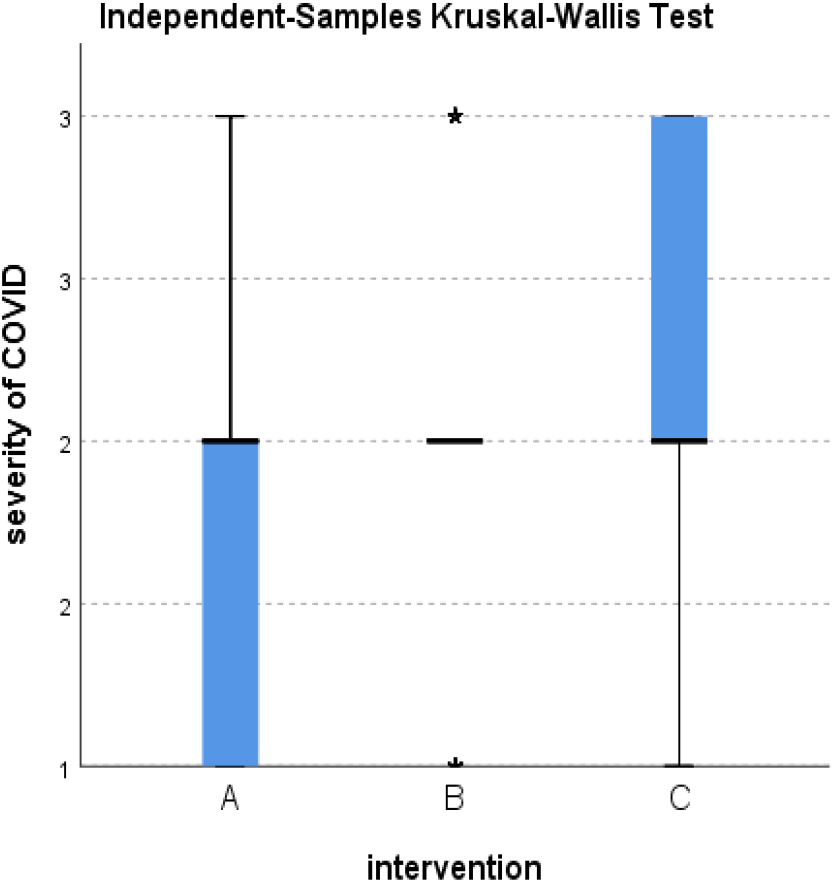
Distribution of COVID-19 severity across the three groups.

**Figure 11.**
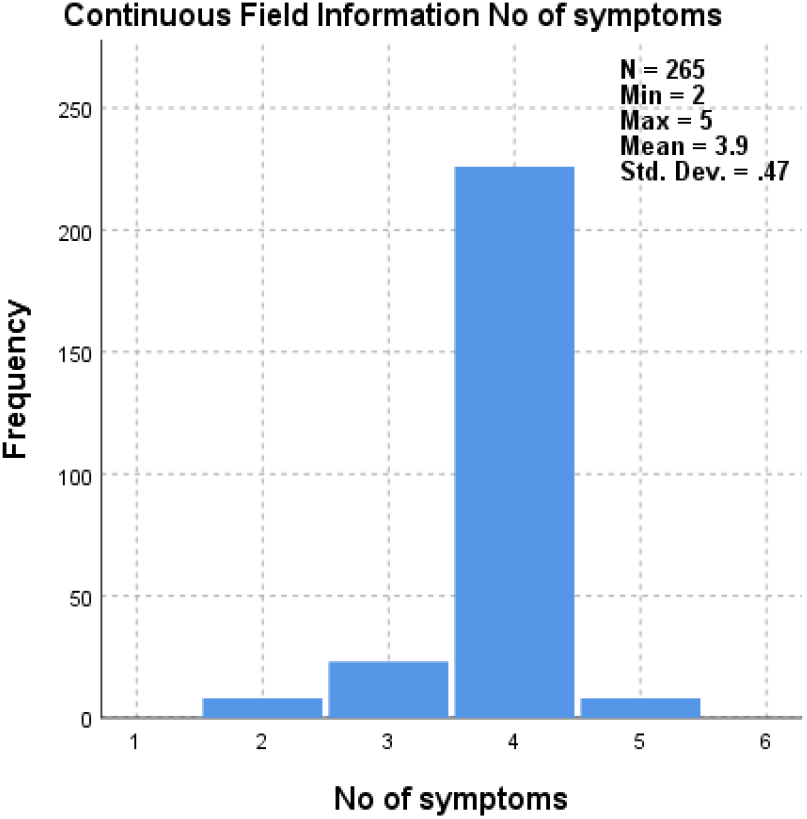
Frequency of number of symptoms in included patients.

**Figure 12.**
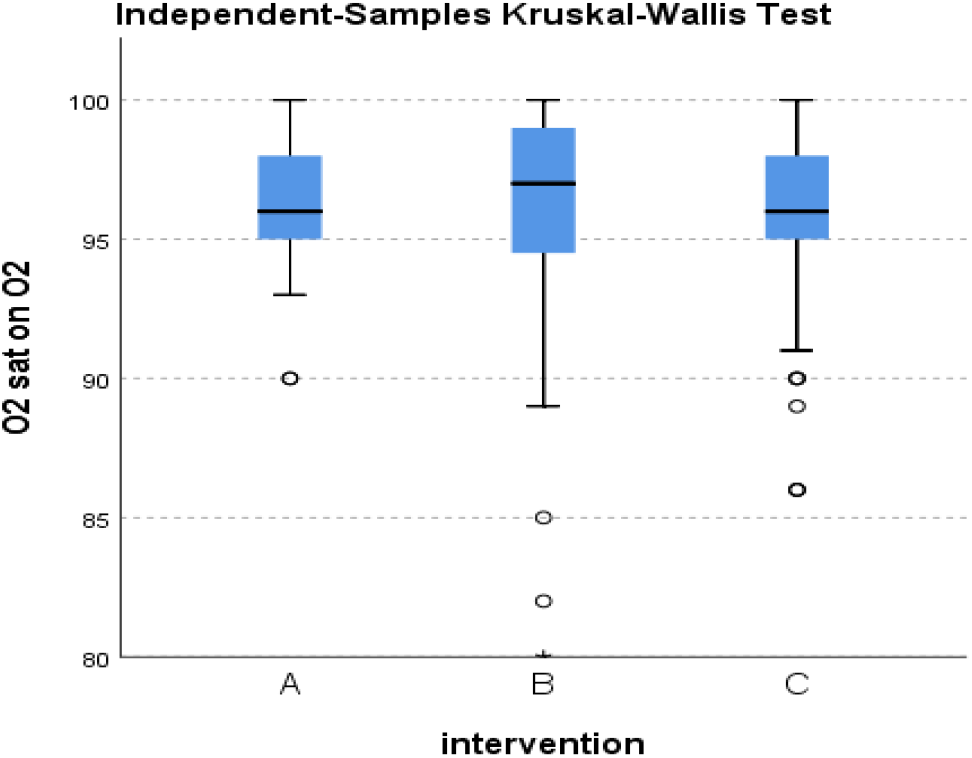
Distribution O2 saturation (on O2 therapy) across the three groups.

**Figure 13.**
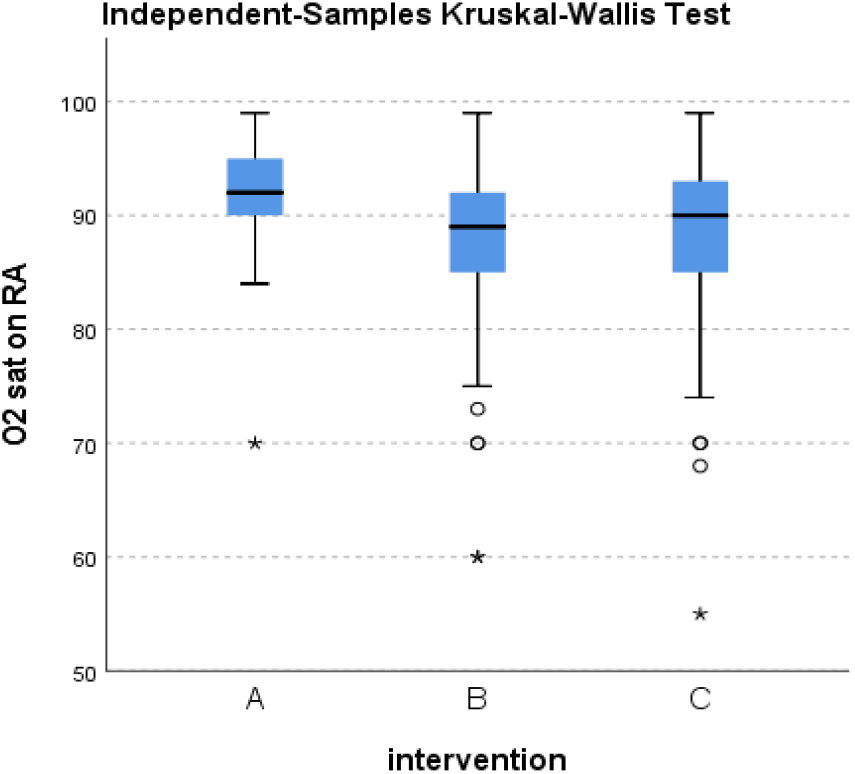
Distribution of O2 saturation (on RA) across the three groups.

**Figure 14.**
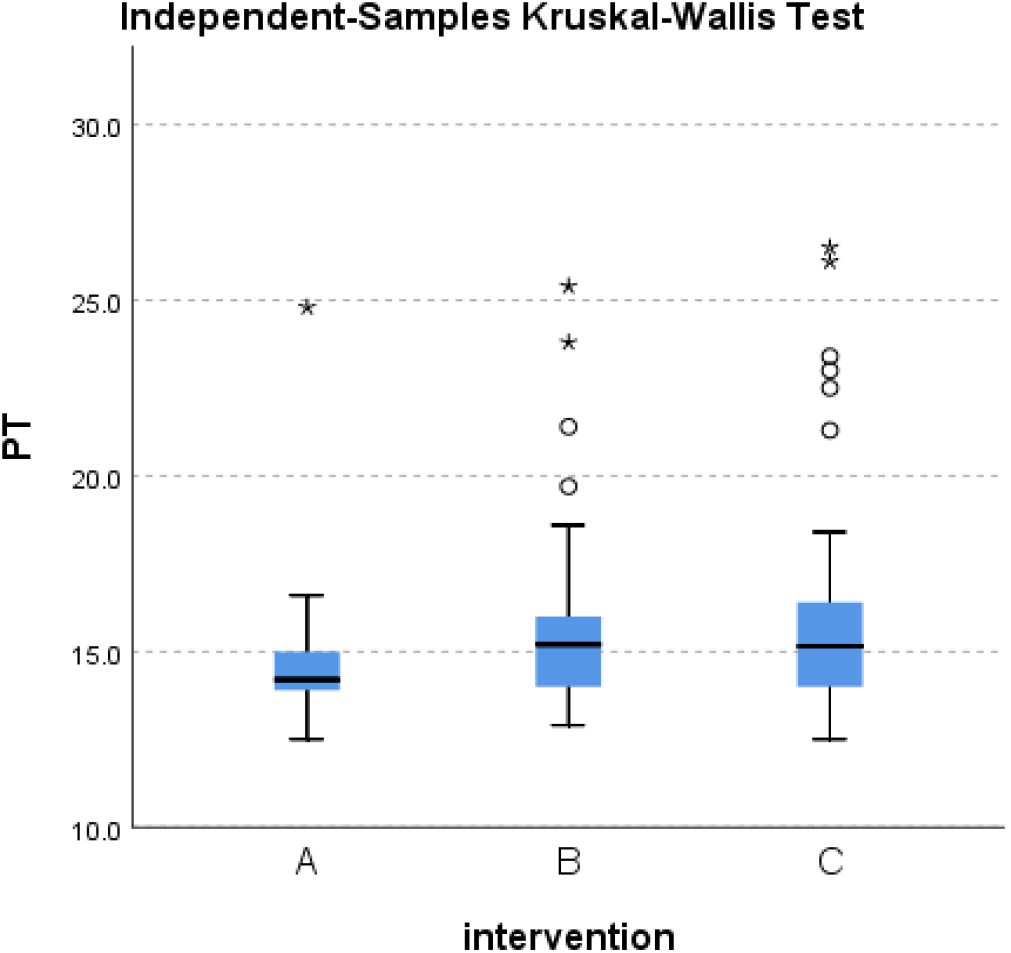
Distribution of PT across the three groups.

**Figure 15.**
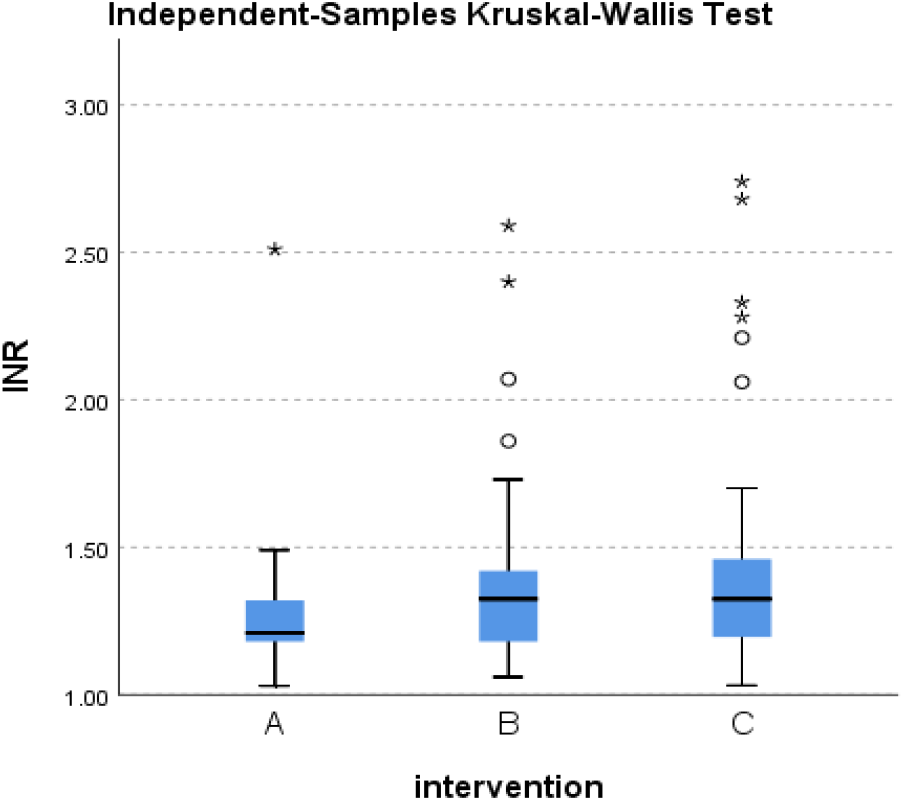
Distribution of INR across the three groups.

**Figure 16.**
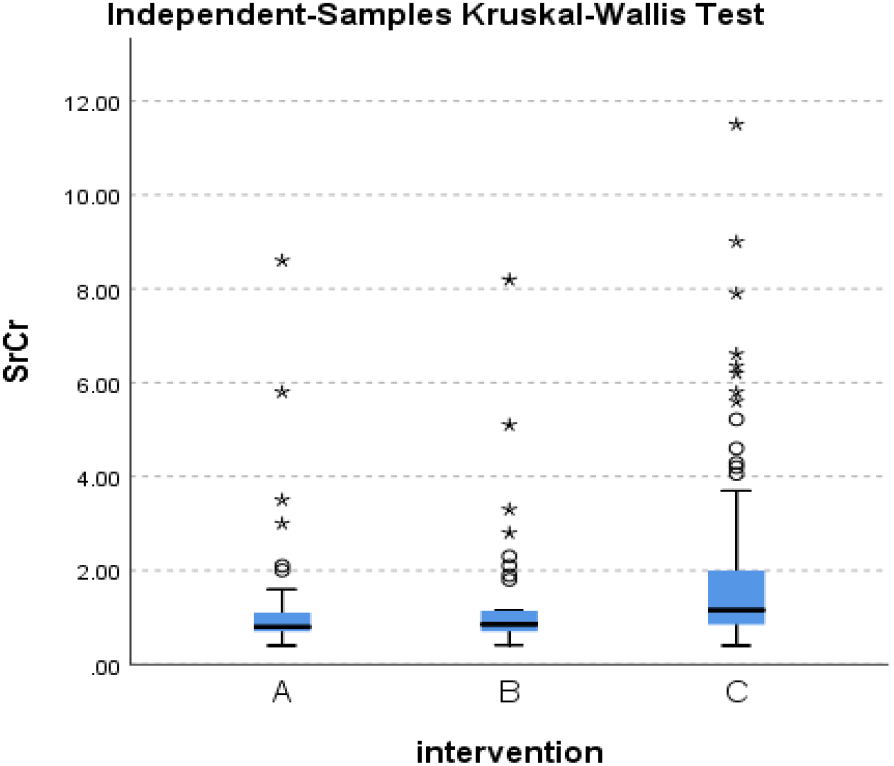
Distribution of S.Cr across the three groups.

**Figure 17.**
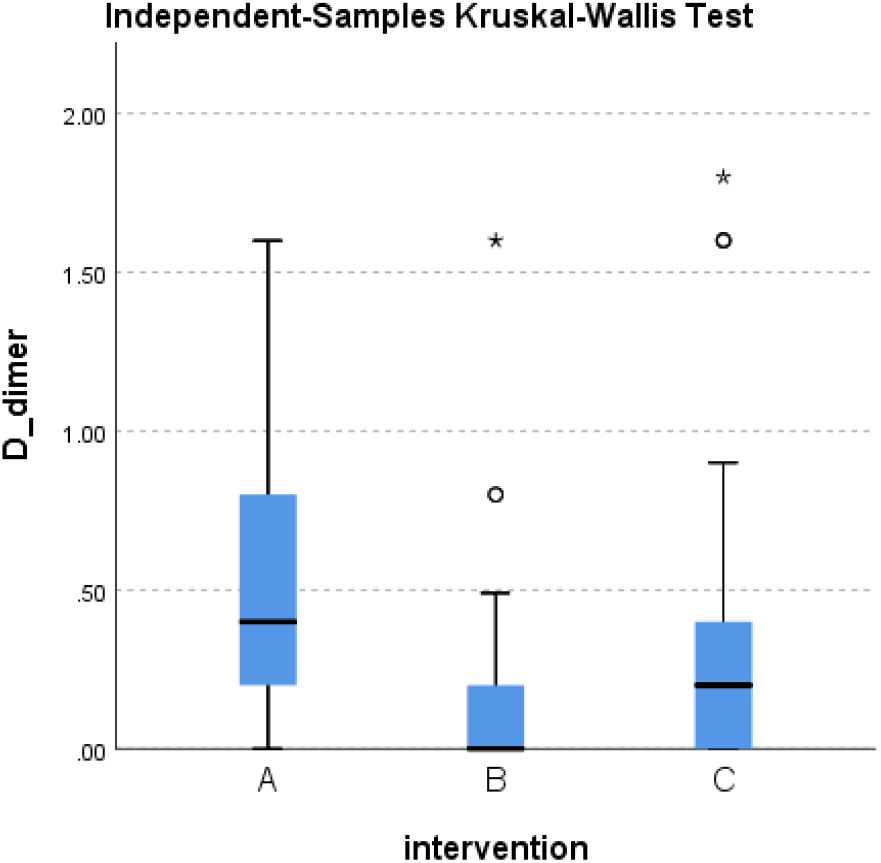
Distribution of D-dimer across the three groups.

**Figure 18.**
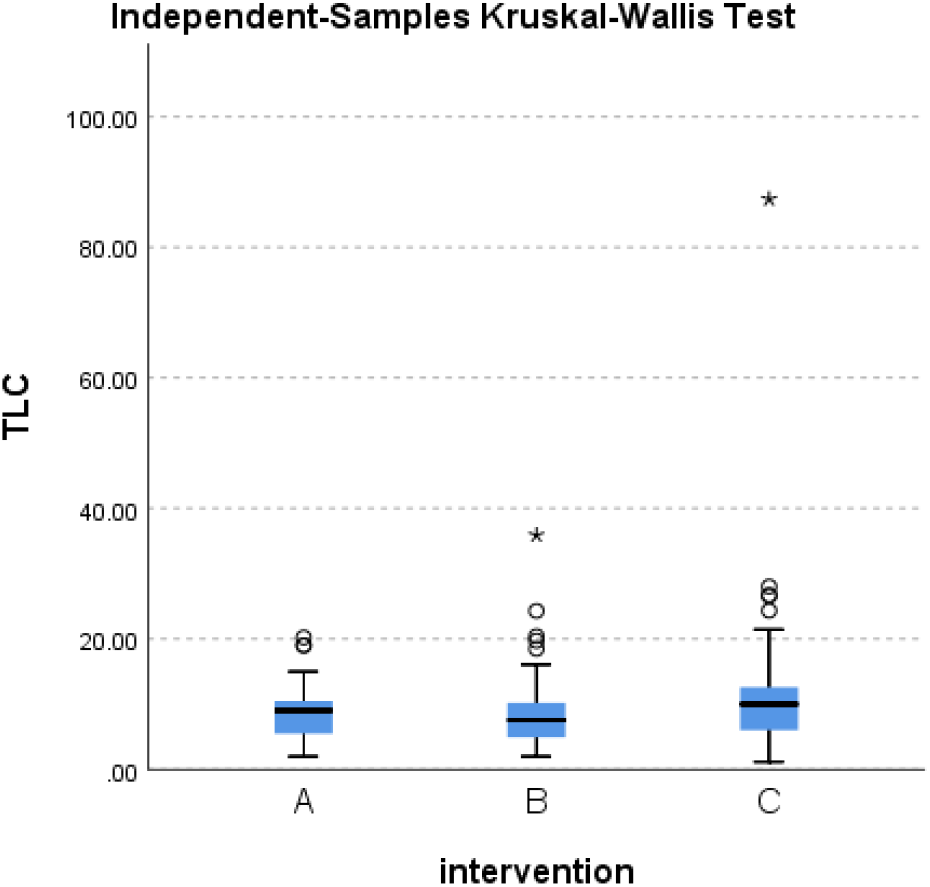
Distribution of TLC across the three groups.

**Figure 19.**
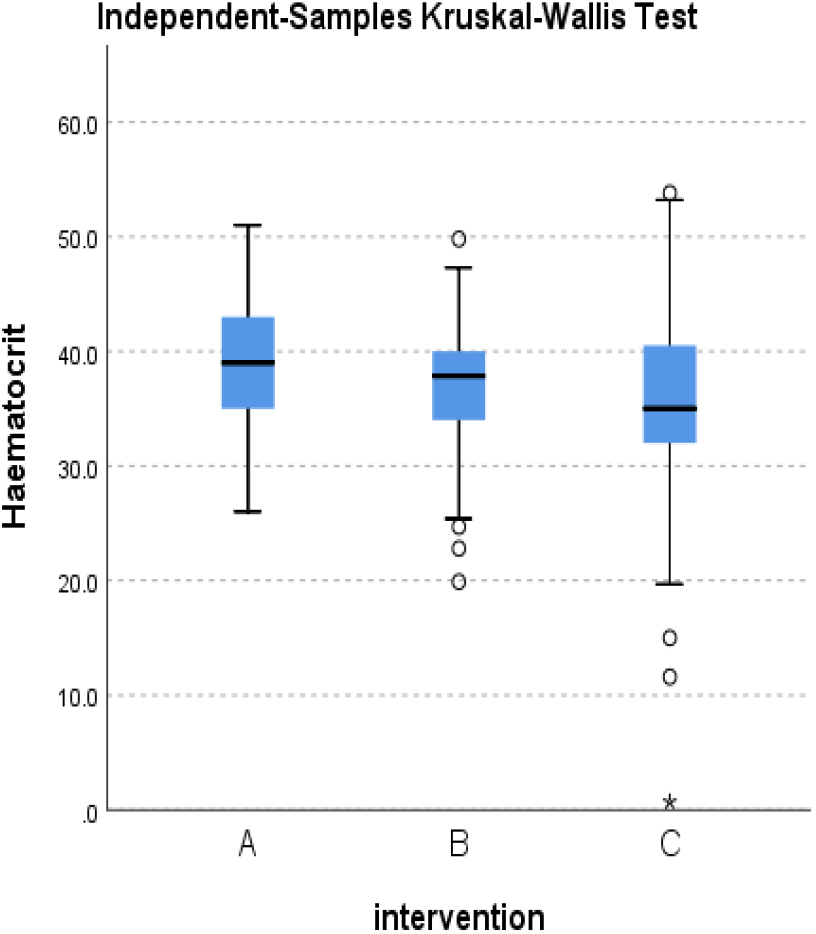
Distribution of hematocrit across the three groups.

**Figure 20.**
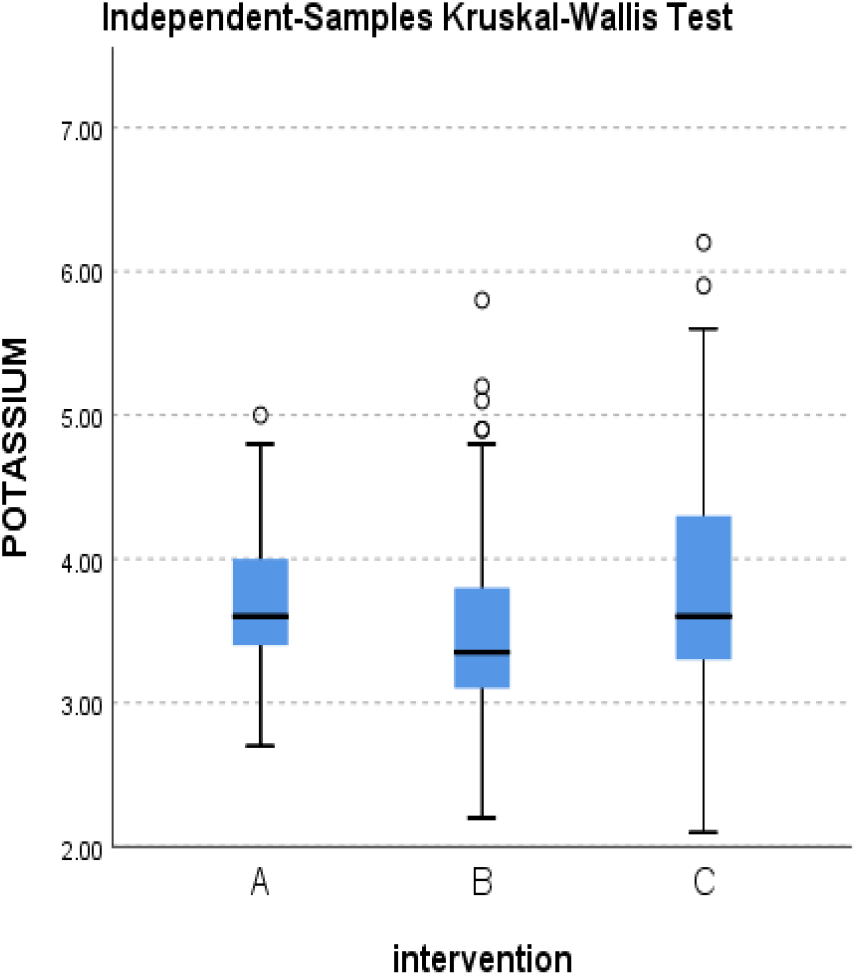
Distribution of Potassium level across the three groups.

**Figure 21.**
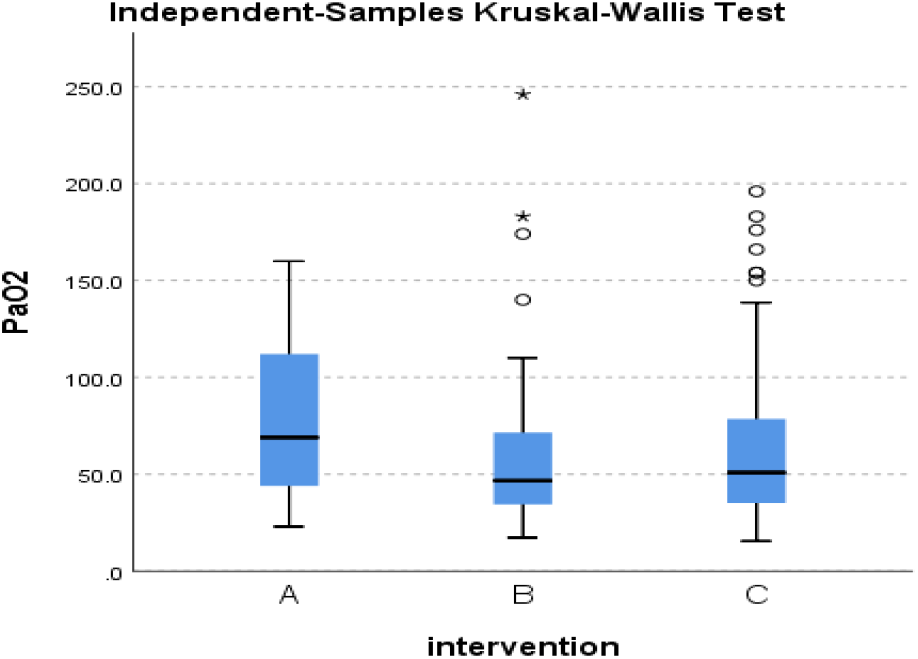
Distribution of PaO2 level across the three groups.

**Figure 22.**
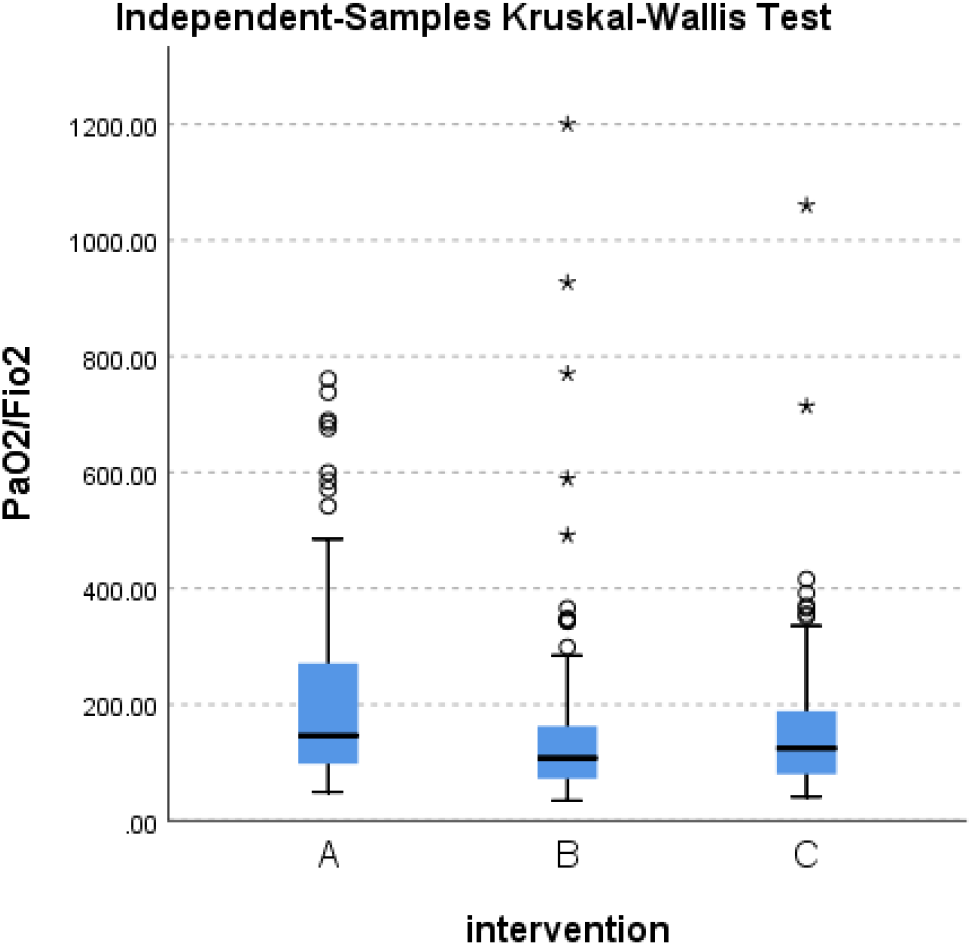
Distribution of PaO2/FiO2 across the three groups.

**Figure 23.**
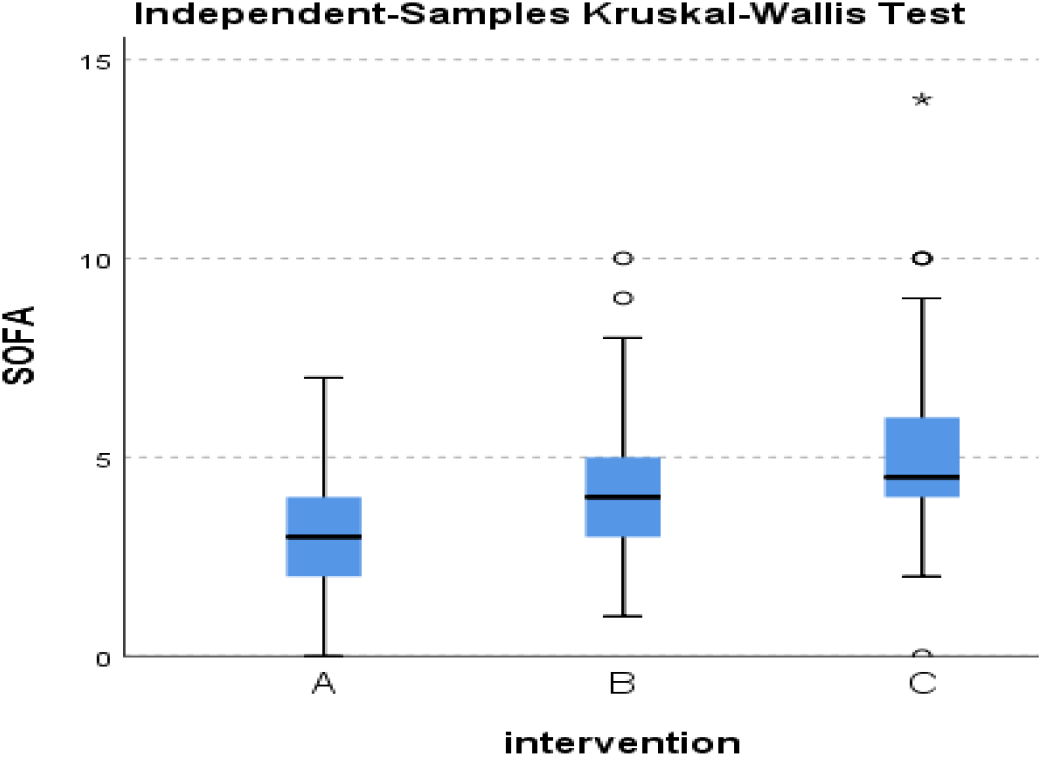
Distribution of SOFA score across the three groups.

#### 8.1.1 Age

There is statistically significant difference between A-C & B-C and statistically non-significant difference between A-B.

#### 8.1.2. Gender

There is a statistically significant difference between B-C and a statistically non-significant difference between A-B & A-C.

#### 8.1.3. Number of comorbidities

There is a statistically significant difference between B-C and a statistically non-significant difference between A-B & A-C.

#### 8.1.4. Method of diagnosis

There is a statistically non-significant difference between the three groups

#### 8.1.5. Severity of COVID-19

There is a statistically significant difference between A-B & A-C and a statistically non-significant difference between C-B. There are statistically significant less severe cases in group A than groups B & C. Frequency of COVID severity in included COVID patients is presented in figure 9.

#### 8.1.6. WHO clinical progression scale

There is a statistically significant difference between A-B & A-C and a statistically non-significant difference between C-B. WHO scale is statistically significantly lower in group A than groups B & C.

#### 8.1.7. Number of symptoms

There is a statistically significant difference between A-B & A-C and a statistically non-significant difference between C-B.

#### 8.1.8. Antibiotics use

In general, there is statistically a non-significant difference between the three groups in antibiotics use. There is only a statistically significant difference between A-C in macrolides (azithromycin & clarithromycin) uses.

#### 8.1.9. Anticoagulants use (enoxaparin, heparin, and rivaroxaban)

There is a statistically non-significant difference between the three groups in anticoagulant use whether in prophylactic or therapeutic dose.

#### 8.1.10. Antiplatelet use (aspirin, clopidogrel)

There is a statistically significant difference between A-C and a statistically non-significant difference between A-B & C-B.

#### 8.1.11. Steroids use (dexamethasone, prednisolone and methylprednisolone)

There is a statistically significant difference between A-B and a statistically non-significant difference between A-C & C-B.

#### 8.1.12. Additive therapy uses (paracetamol, vitamin C, zinc, acetyl cysteine, lactoferrin)

In general; there is statistically non-significant difference between the three groups in additive therapy use. There is only a statistically significant difference between A-C in paracetamol and zinc use & A-B in zinc use.

#### 8.1.13. Oxygen therapy use

In general, there is a statistically significant difference between A-B & A-C and statistically non-significant difference between C-B. There is a statistically non-significant difference between the three groups in nasal prongs and HFNC use.

There is statistically significant difference between A-B & A-C in simple face mask (SFM), Continuous Positive Airway Pressure (CPAP) or Non-Invasive Ventilation (NIV) and IMV use.

There is a statistically significant difference between B-C in mask reservoir (MR) use.

#### 8.1.14. Vasopressor use

There is a statistically significant difference between A-B & A-C in vasopressor use and a statistically non-significant difference between C-B. Use of vasopressors in group A is statistically significantly less than groups B & C.

#### 8.1.15. Prone positioning

There is a statistically nonsignificant difference between the three groups.

#### 8.1.16. Vital signs

There is a statistically non-significant difference between the three groups in heart rate, respiratory rate and temperature.

#### 8.1.17. Oxygen saturation

There is a statistically non-significant difference between the three groups in O2 saturation on O2 therapy. But a statistically significant difference exists between A-B & A-C in O2 saturation on Room air (RA).

#### 8.1.18. Liver function tests

There is a statistically non-significant difference between the three groups in liver enzymes (AST & ALT), bilirubin, and albumin levels

#### 8.1.19. Coagulation profile

There is a statistically significant difference between A-B & A-C in PT and INR.

#### 8.1.20. Kidney function test

There is a statistically significant difference between A-C & B-C in serum creatinine.

#### 8.1.21. Inflammatory markers

There is a statistically non-significant difference between the three groups in CK, LDH, Ferritin and CRP levels

There is a statistically significant difference between the three groups in D-dimer.

#### 8.1.22. Blood picture

There is a statistically significant difference between B-C in TLC and A-B & A-C in hematocrit.

There is a statistically non-significant difference between the three groups in lymphocytic counts, hemoglobin and platelets.

#### 8.1.23. Electrolytes

There is a statistically non-significant difference between the three groups in sodium level. There is a statistically significant difference between B-C in Potassium level.

#### 8.1.24. Blood gases

There is a statistically significant difference between A-B & A-C in PaO2 and between A-B in PaO2/FiO2. There is a statistically non-significant difference between the three groups in PaCO2.

#### 8.1.25. Consciousness level

There is a statistically significant difference between A-C & B-C in GCS and statistically non-significant difference between A-B.

#### 8.1.26. Multi-Organ Functions Assessment

There is a statistically significant difference between the three groups in SOFA score.

#### 8.1.27. Regression analysis

Regression analysis is performed to explore the effect of baseline characteristics (that show a statistically significant difference between the three groups) on the outcomes of the study and the possibility of existence of confounding variables as shown in table 4.

**Table 4.**
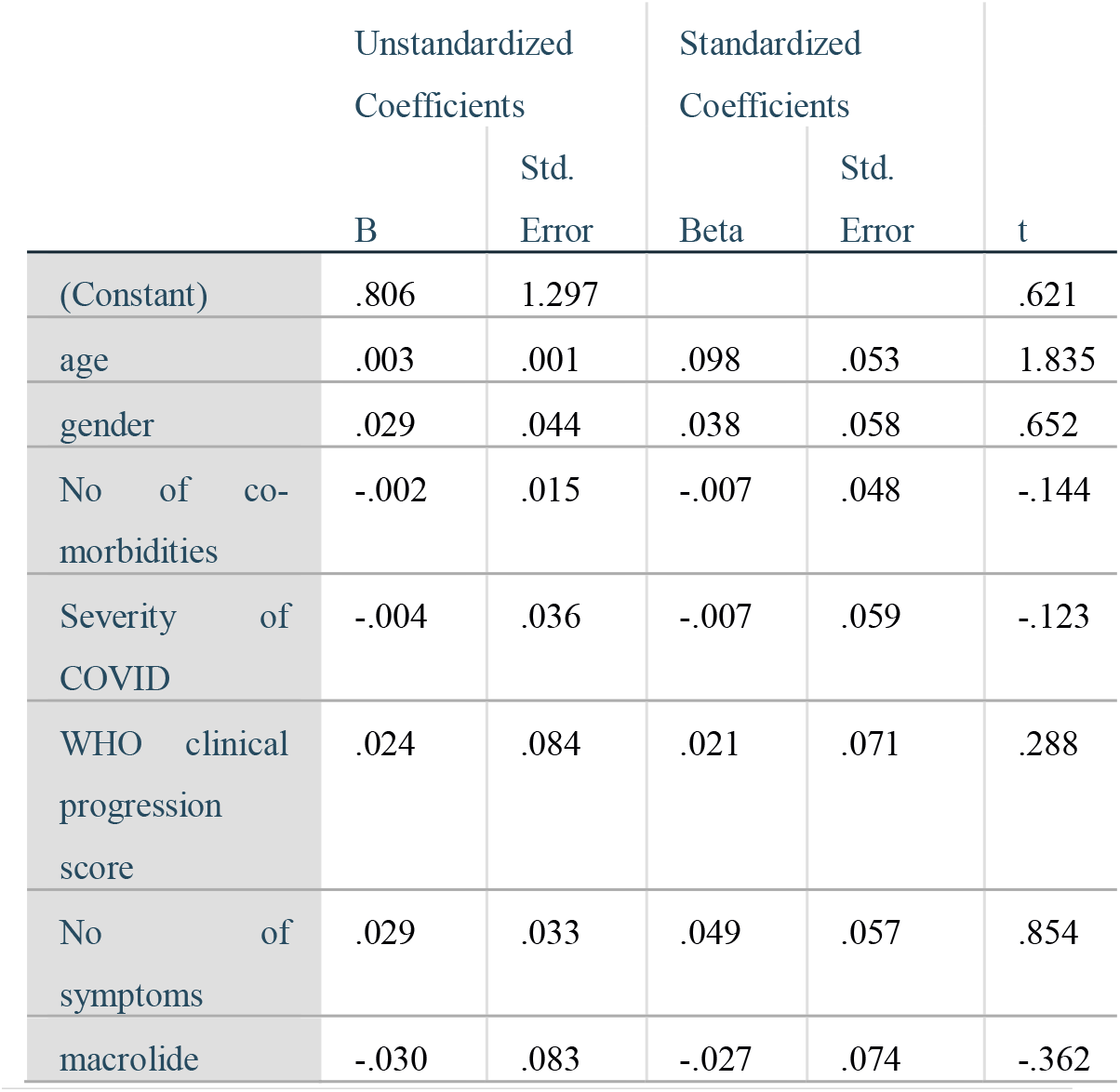
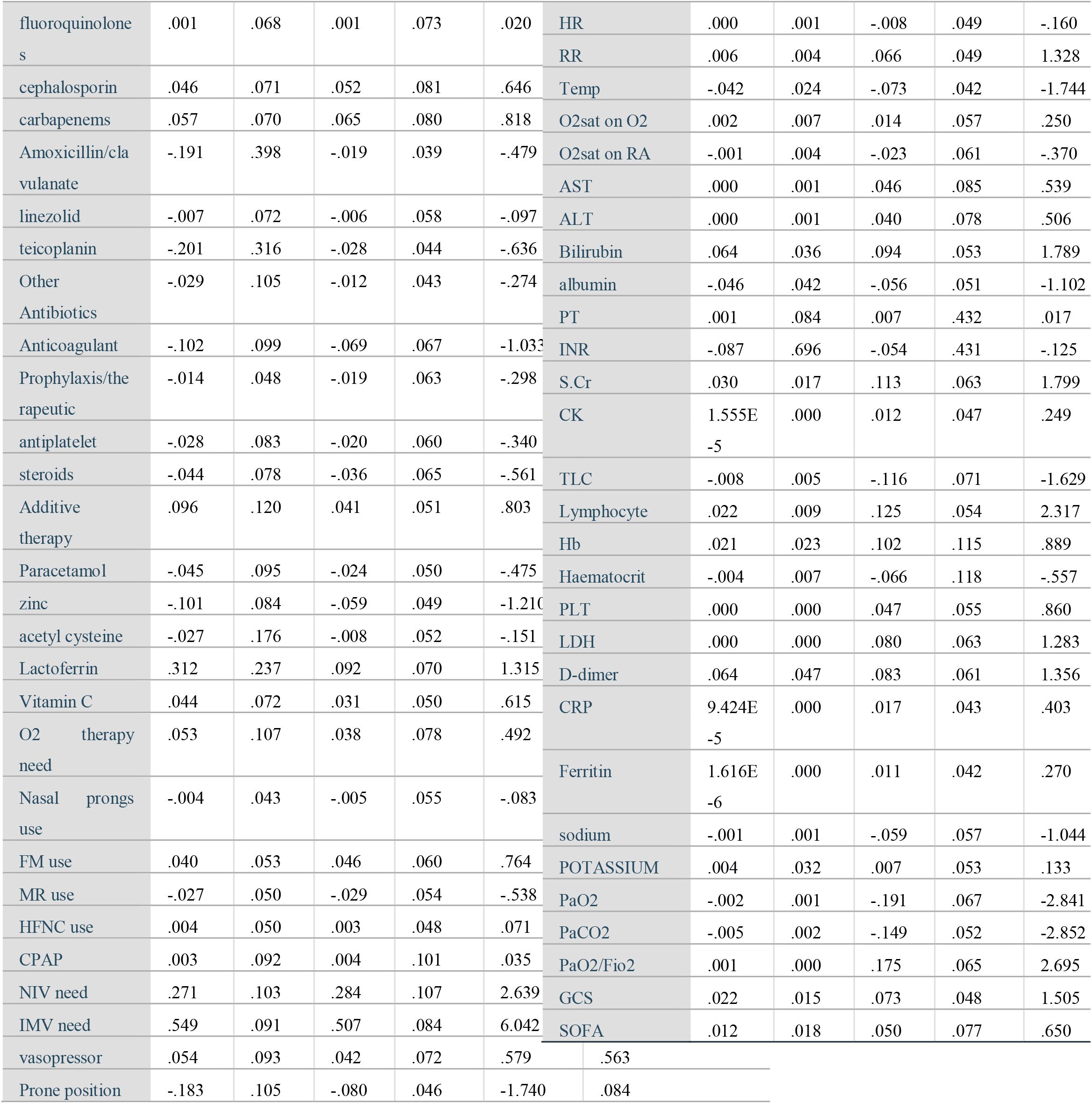
the best regression model for studying the effects of confounding variables on 28-day mortality

#### 8.2. Regarding outcomes of the study after intervention in the three groups

Table 5 shows the significance of difference between the three groups and also includes a pairwise comparison between every two groups in clinical outcomes if they show statistically significant difference between the three groups. Figures (24-78) show the distributions and frequencies of these outcomes across the three groups.

**Table 5.**
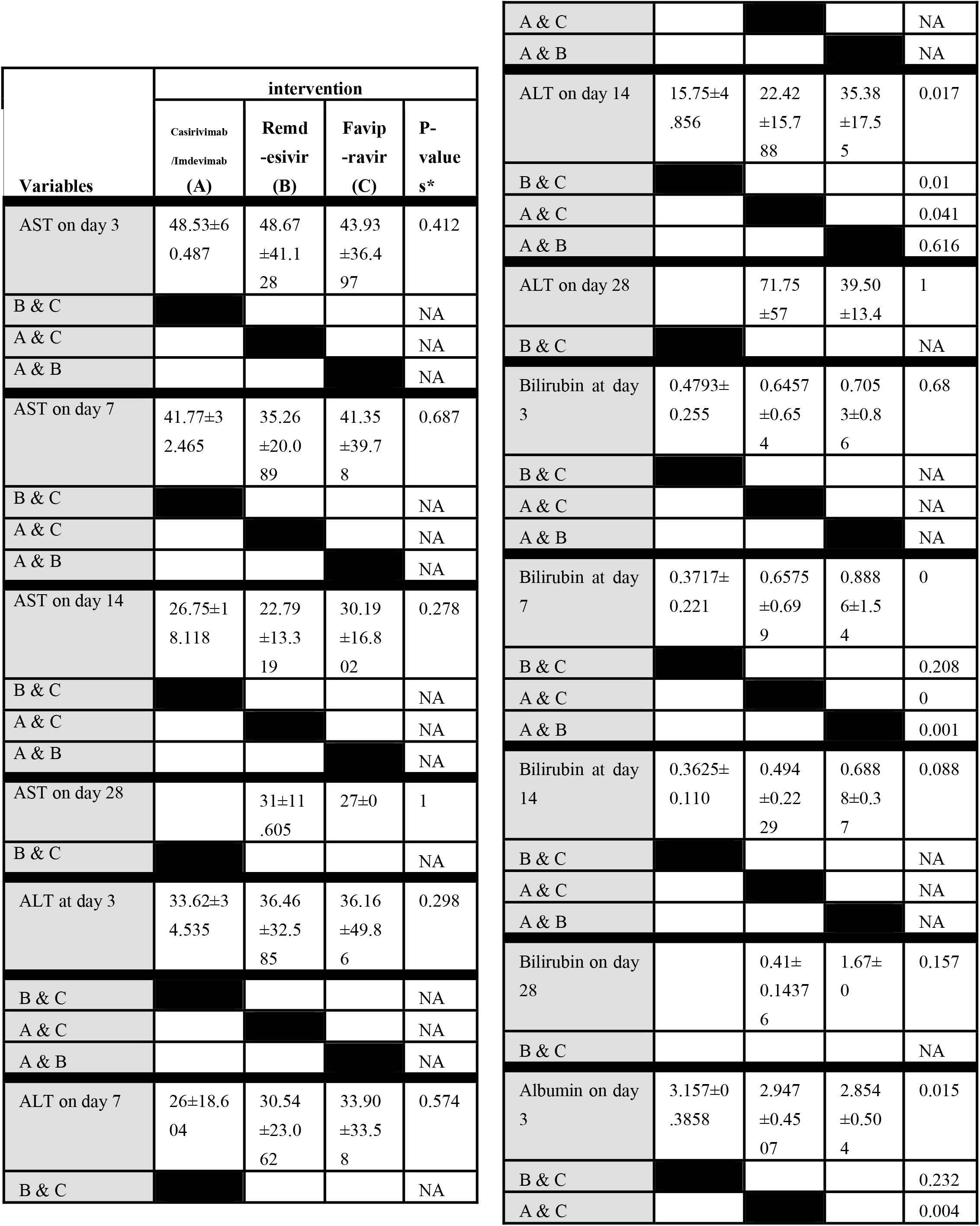
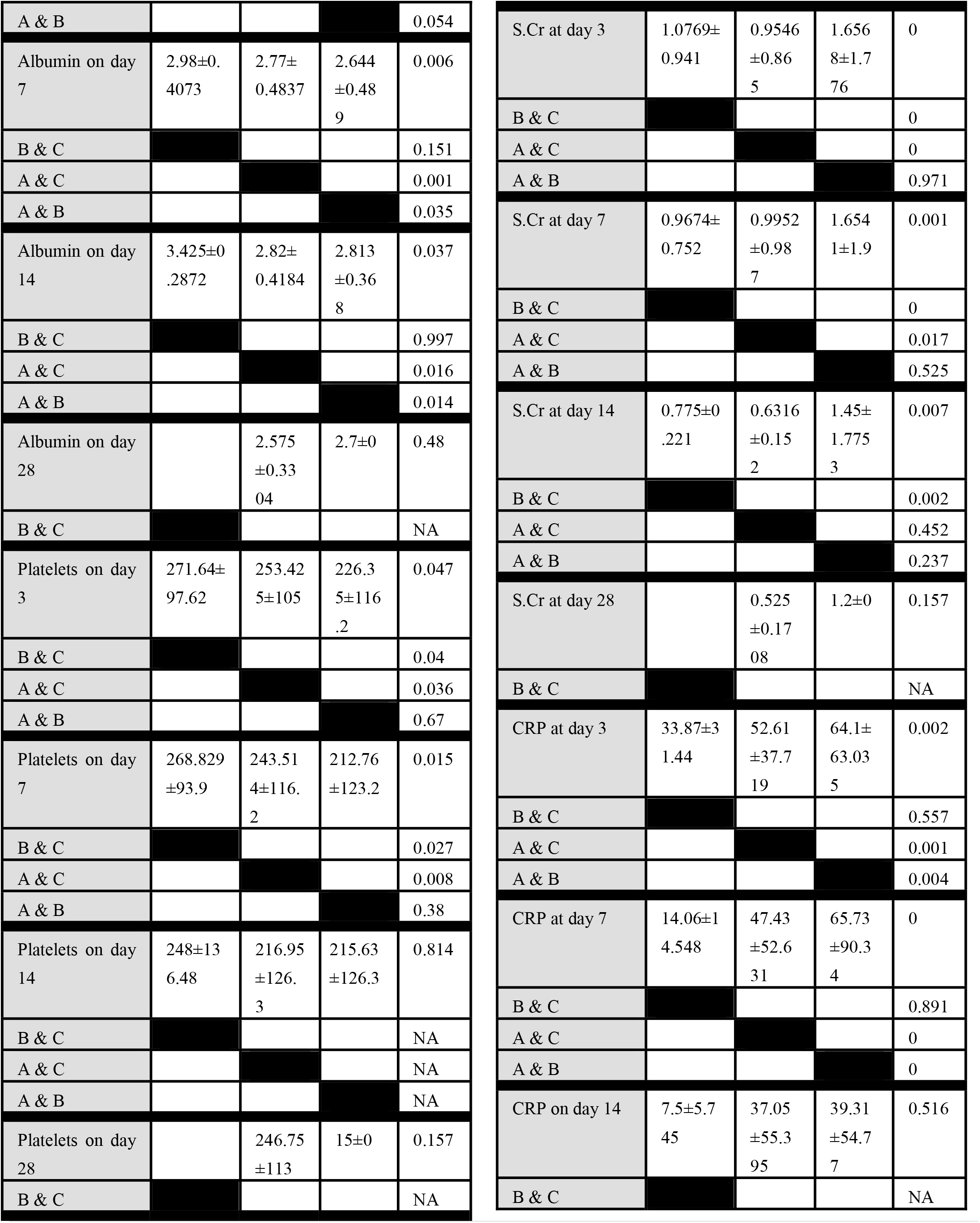
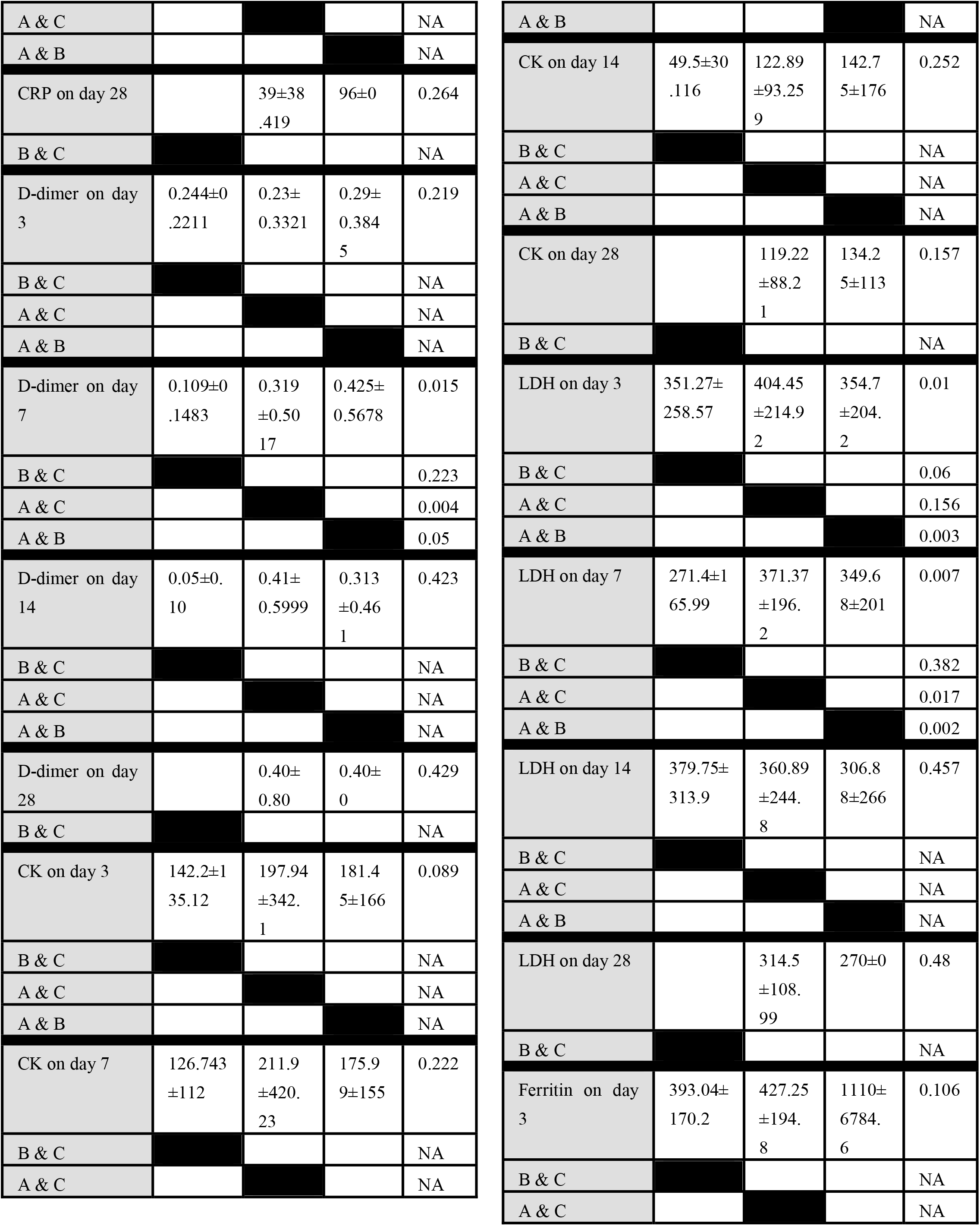
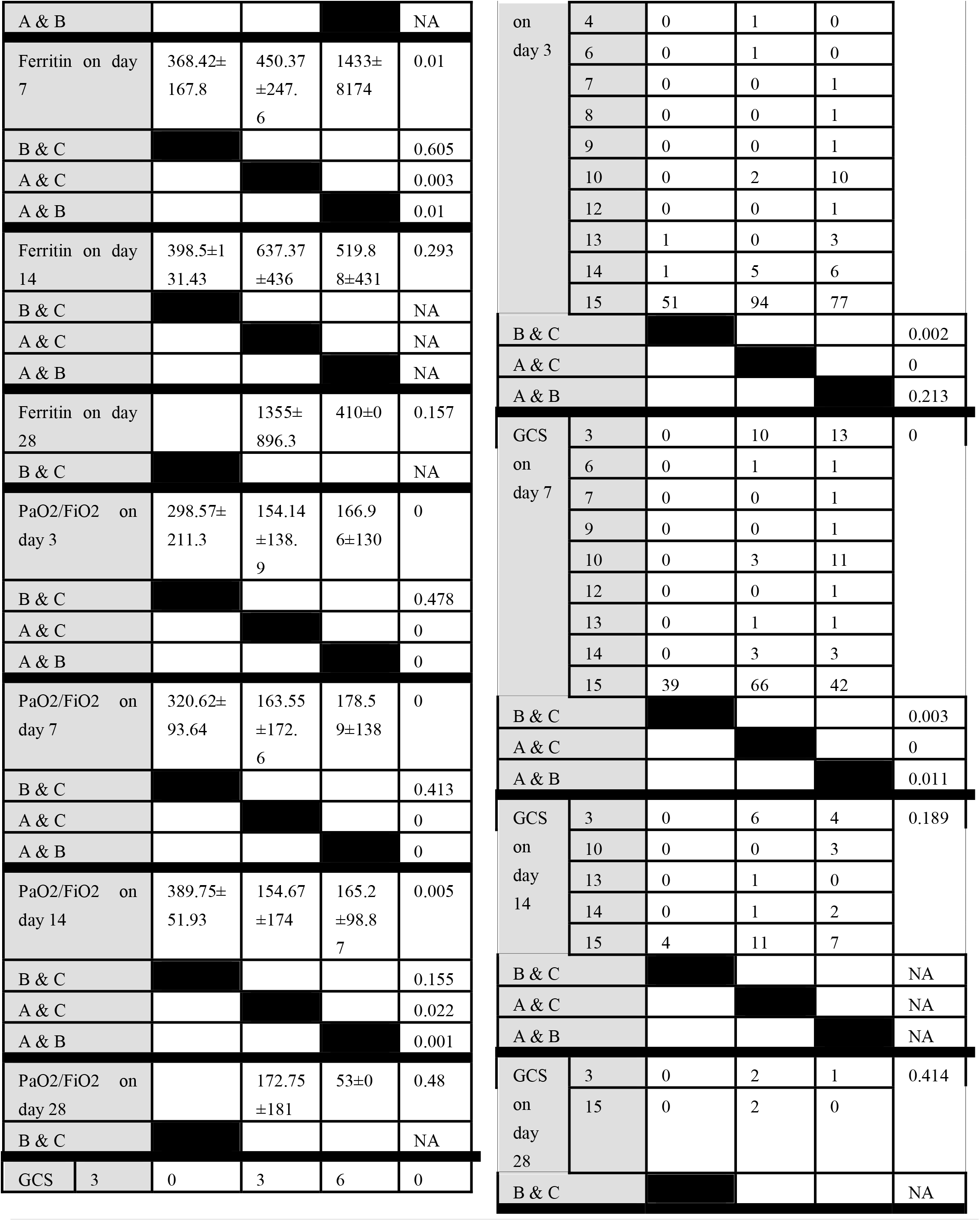
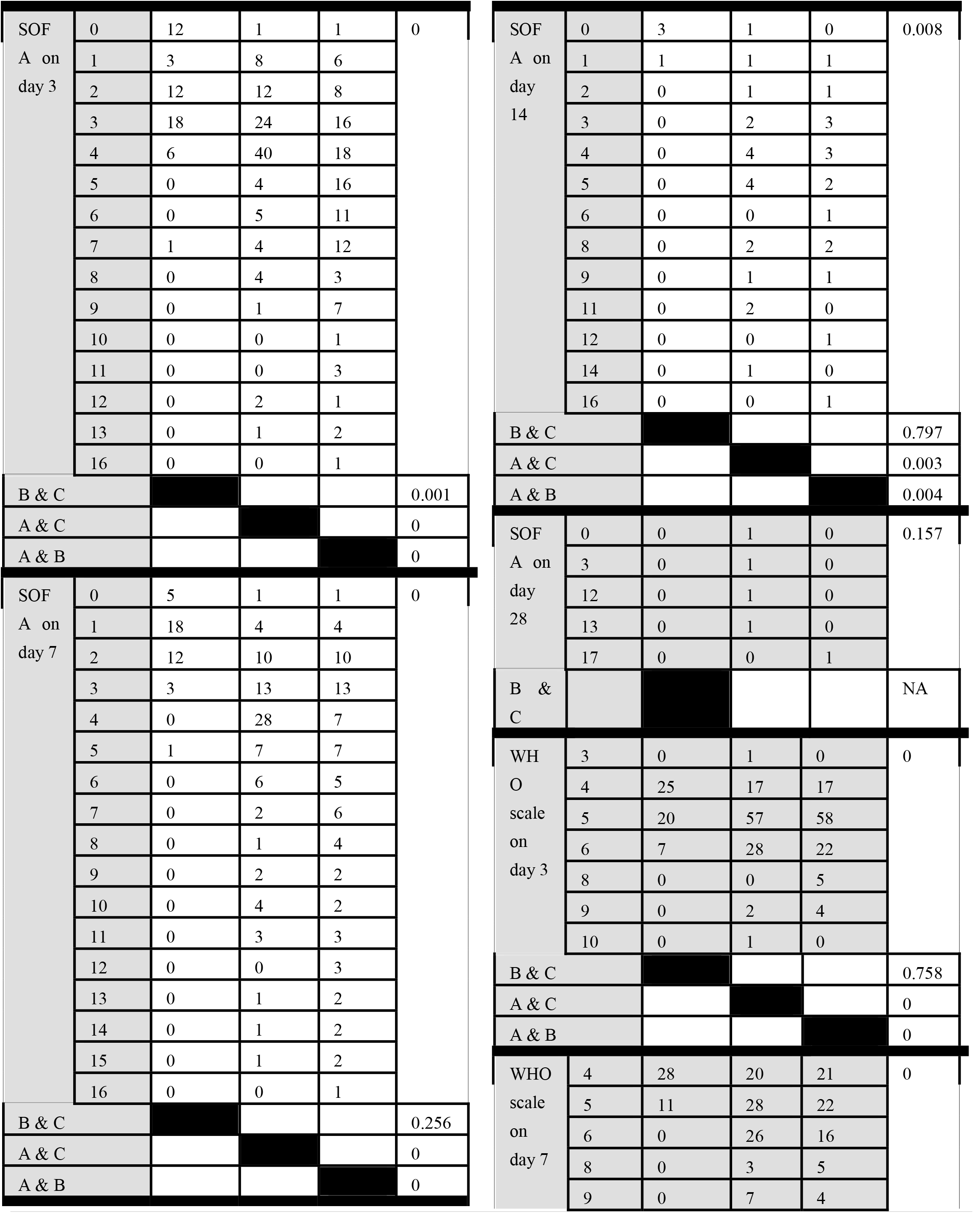
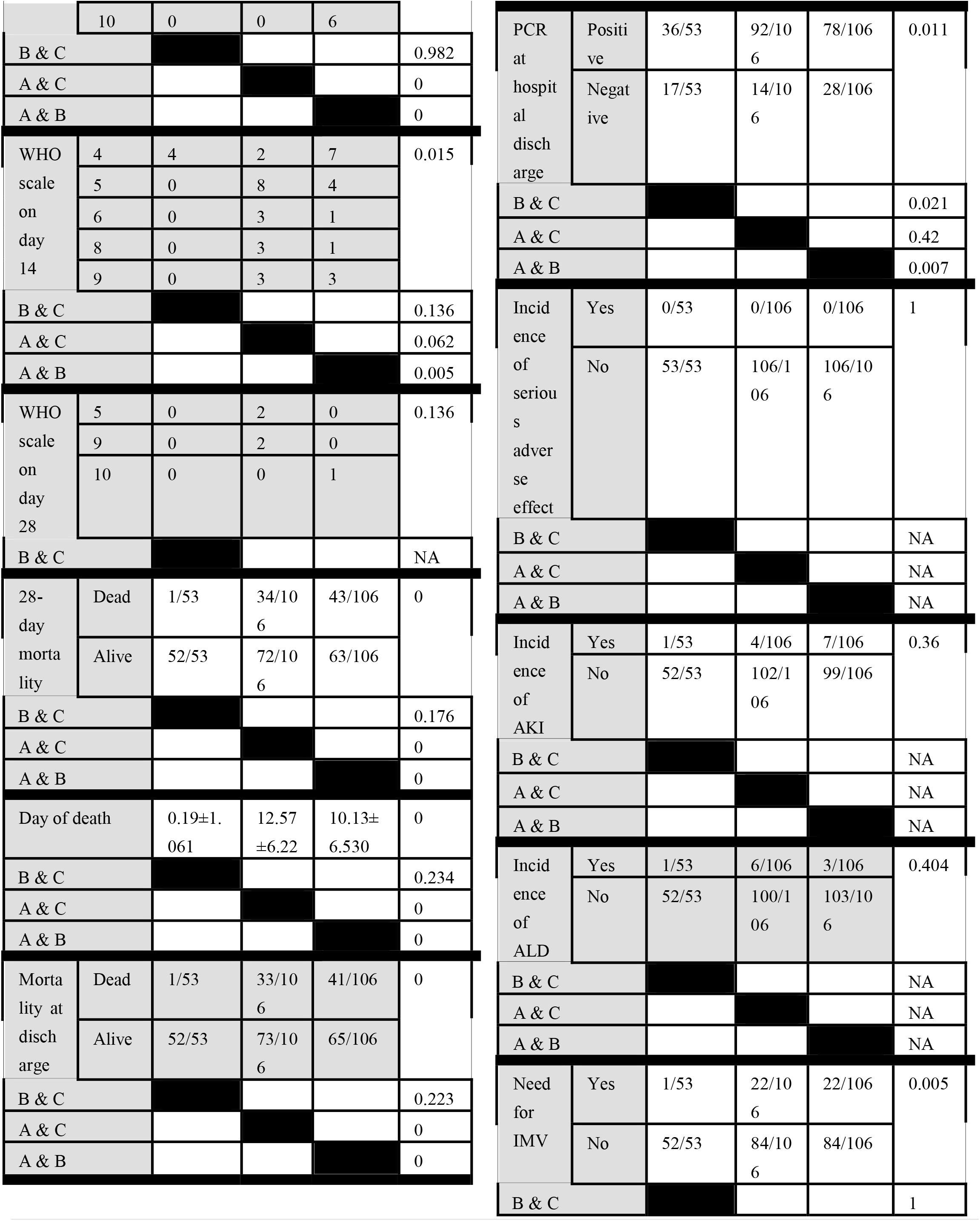
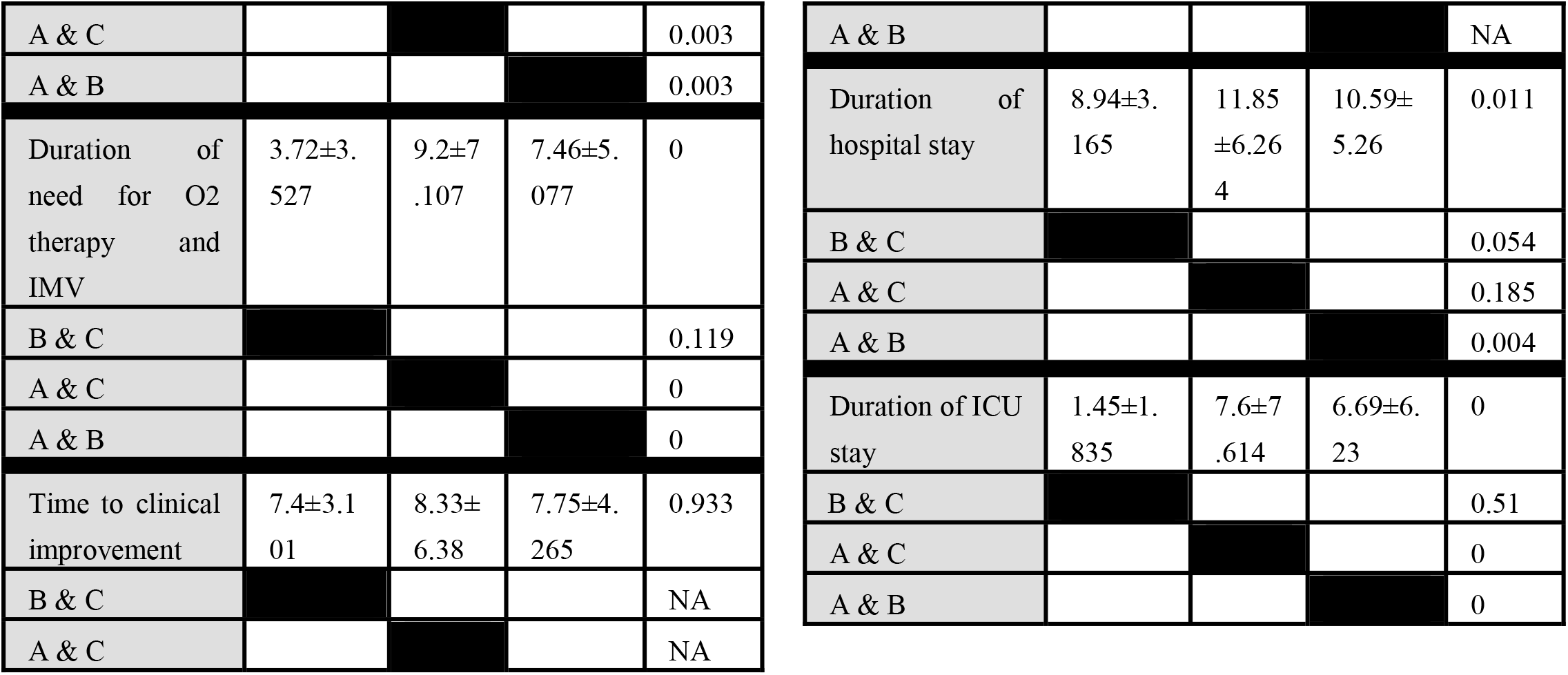
the Significance of differences in baseline characteristics between the three groups

**Figure 24.**
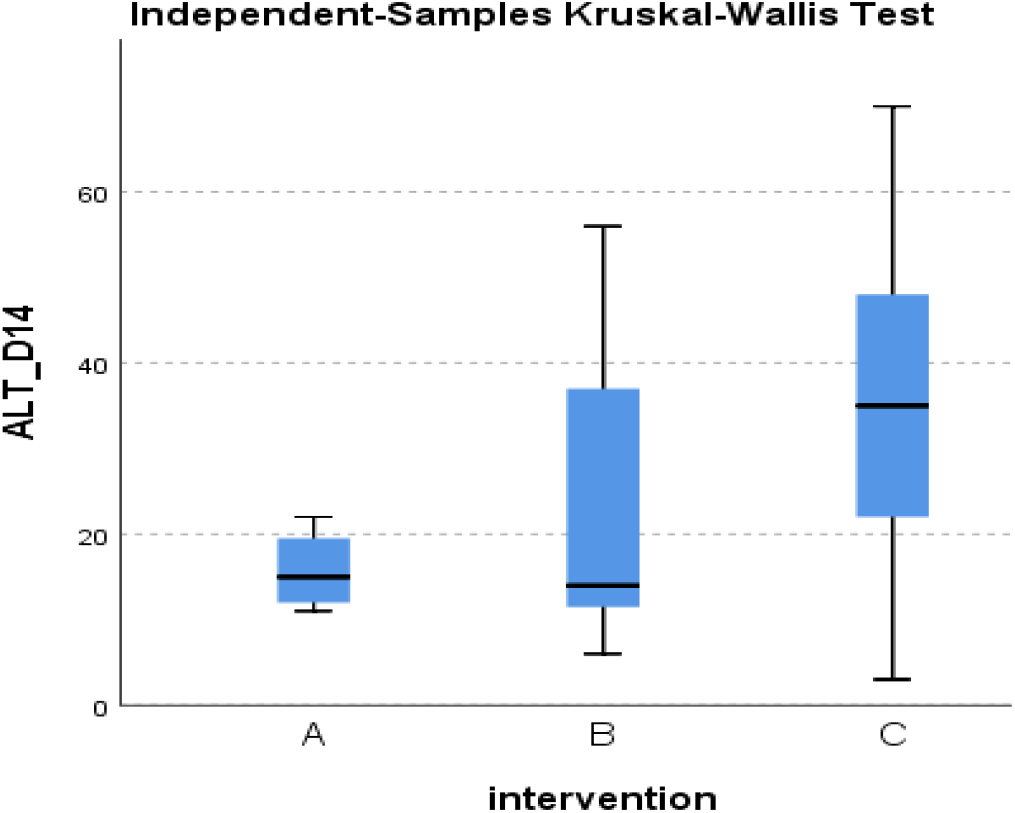
Distribution of ALT at day 14 across the three groups.

**Figure 25.**
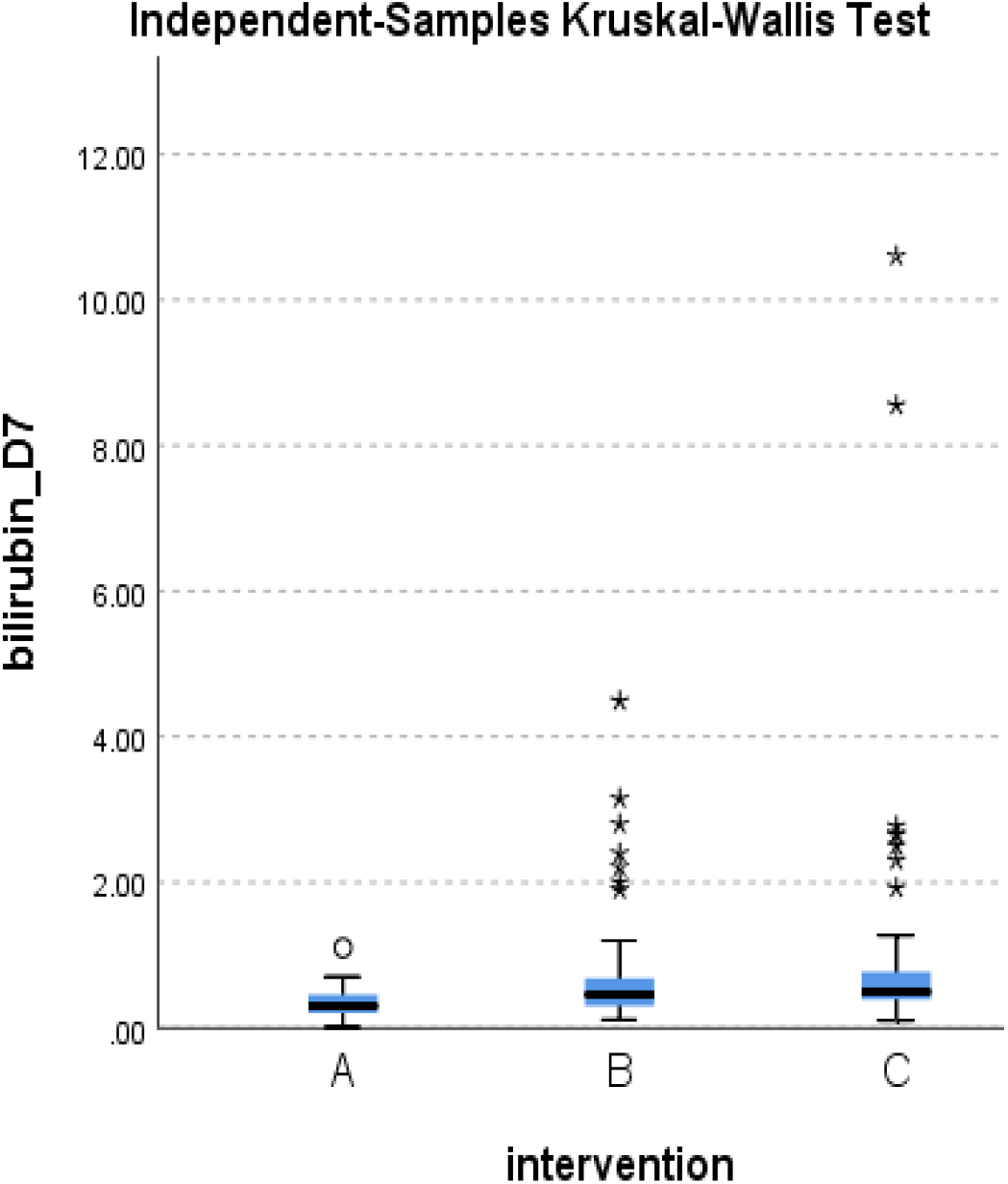
Distribution of Bilirubin at day 7 across the three groups.

**Figure 26.**
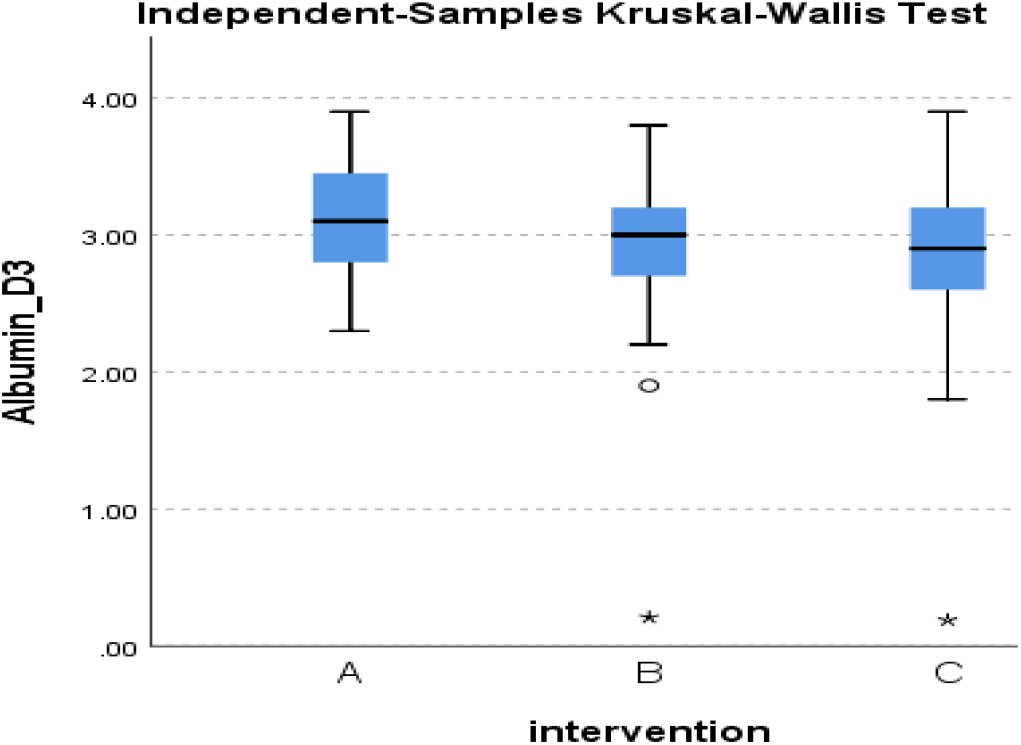
Distribution of Albumin at day 3 across the three groups.

**Figure 27.**
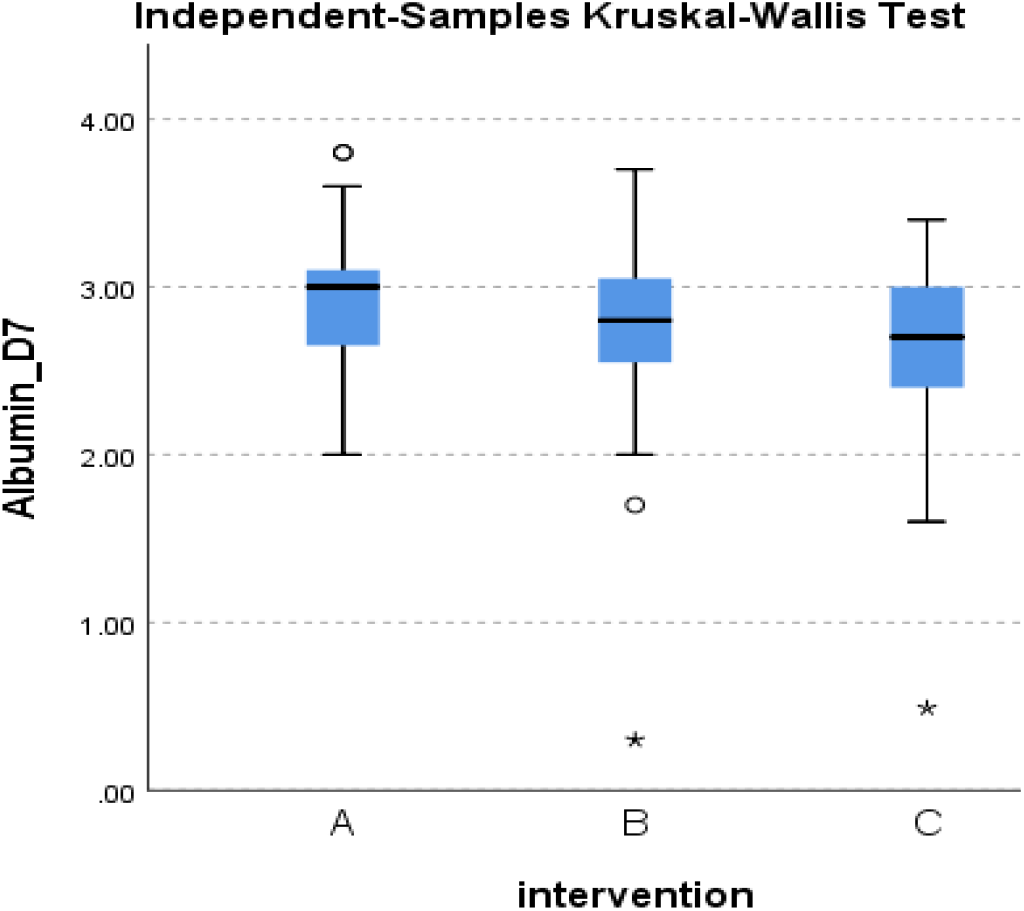
Distribution of Albumin at day 7 across the three groups.

**Figure 28.**
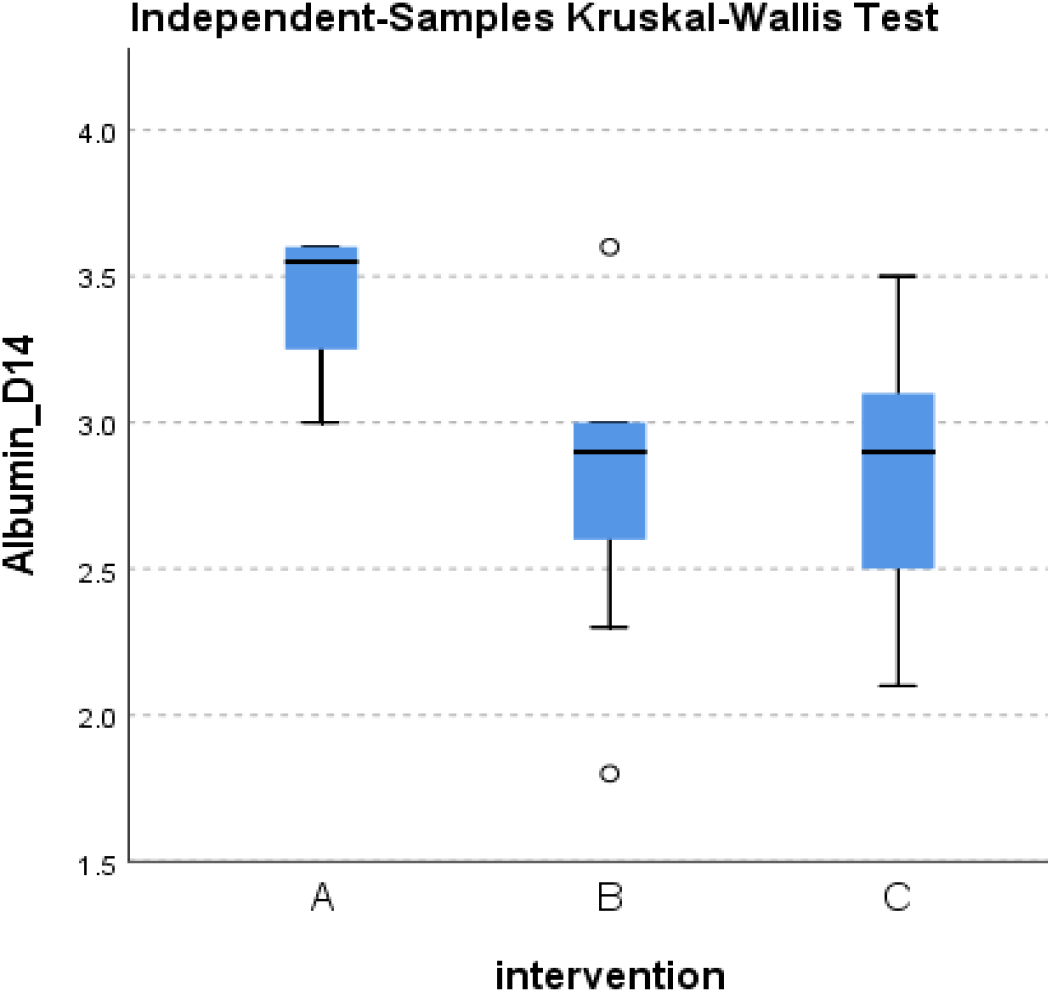
Distribution of Albumin at day 14 across the three groups.

**Figure 29.**
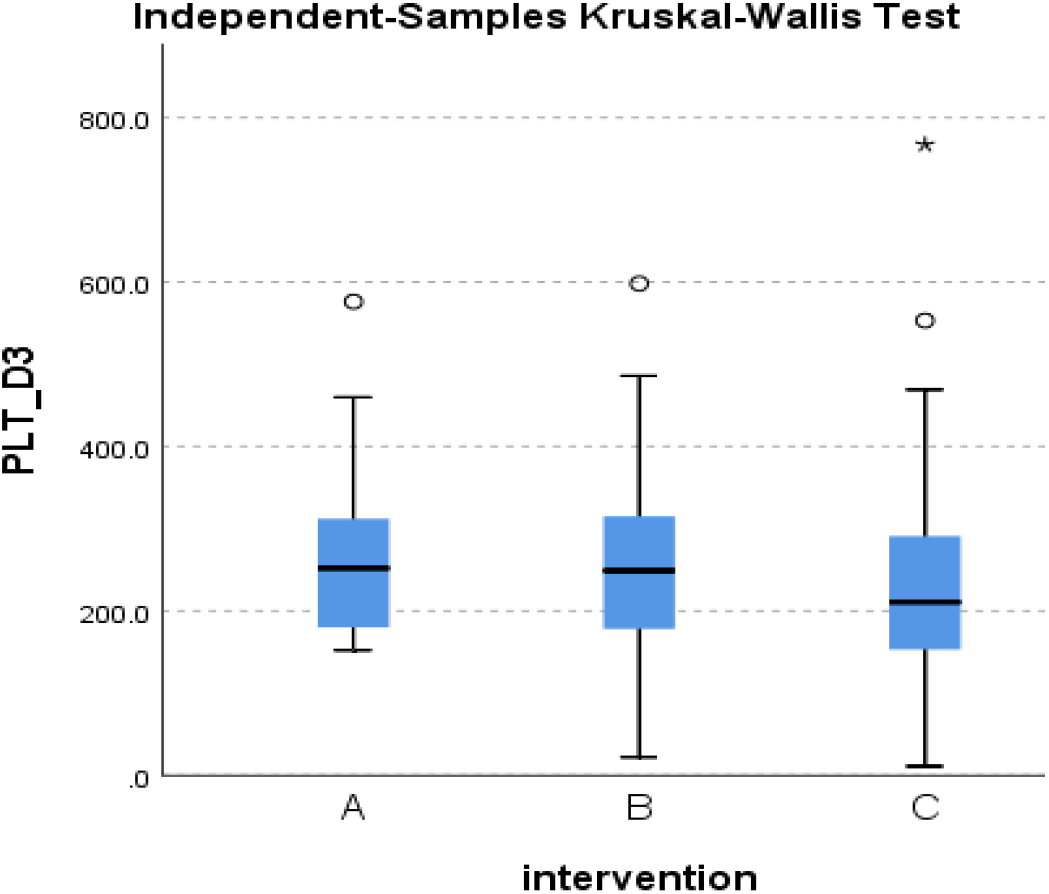
Distribution of platelets at day 3 across the three groups.

**Figure 30.**
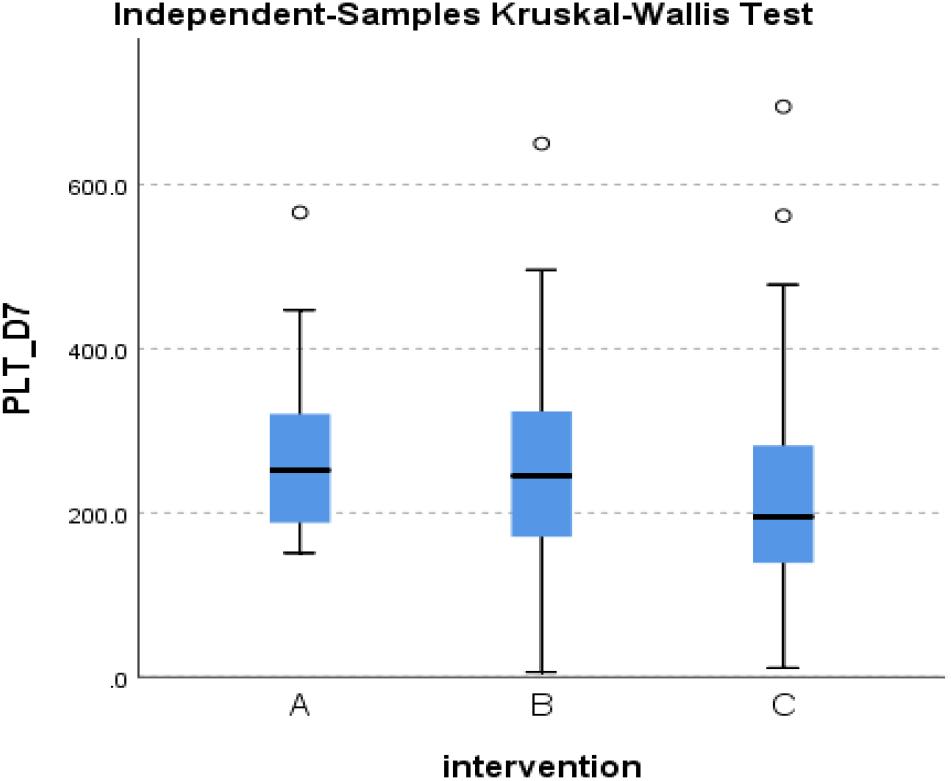
Distribution of platelets at day 7 across the three groups.

**Figure 31.**
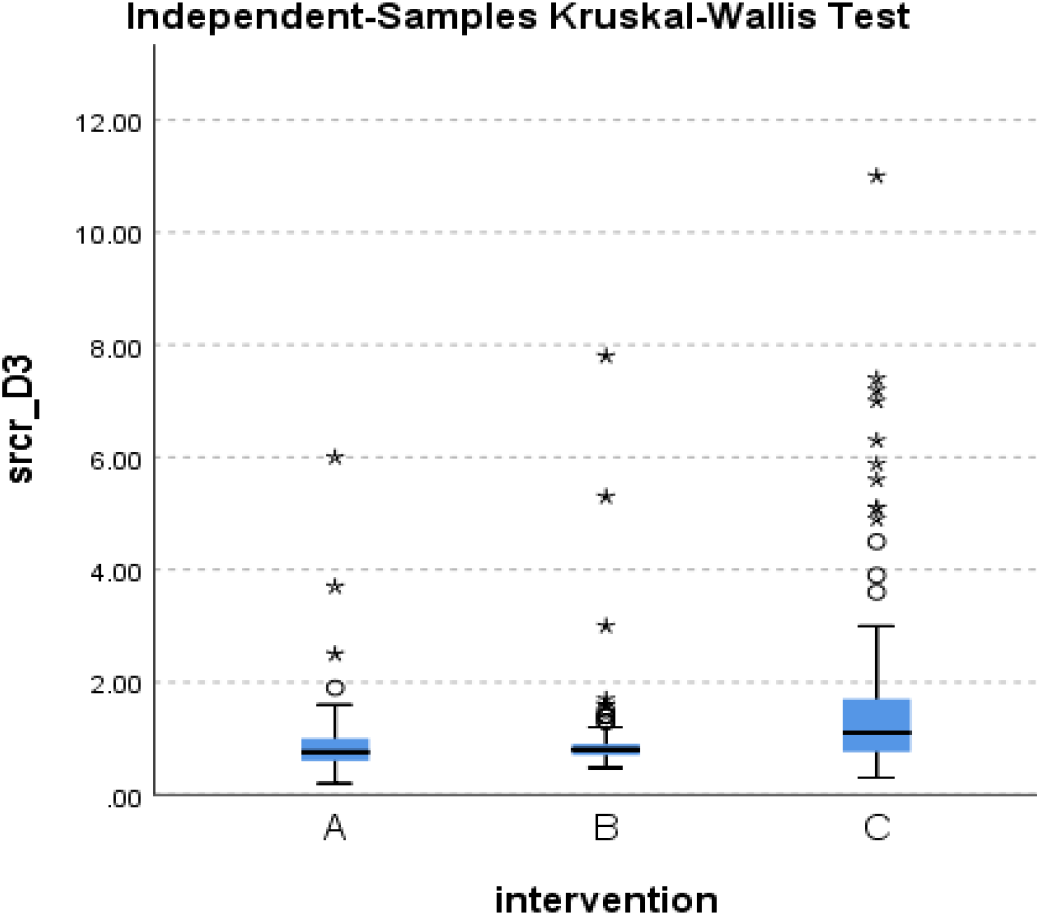
Distribution of S.Cr at day 3 across the three groups.

**Figure 32.**
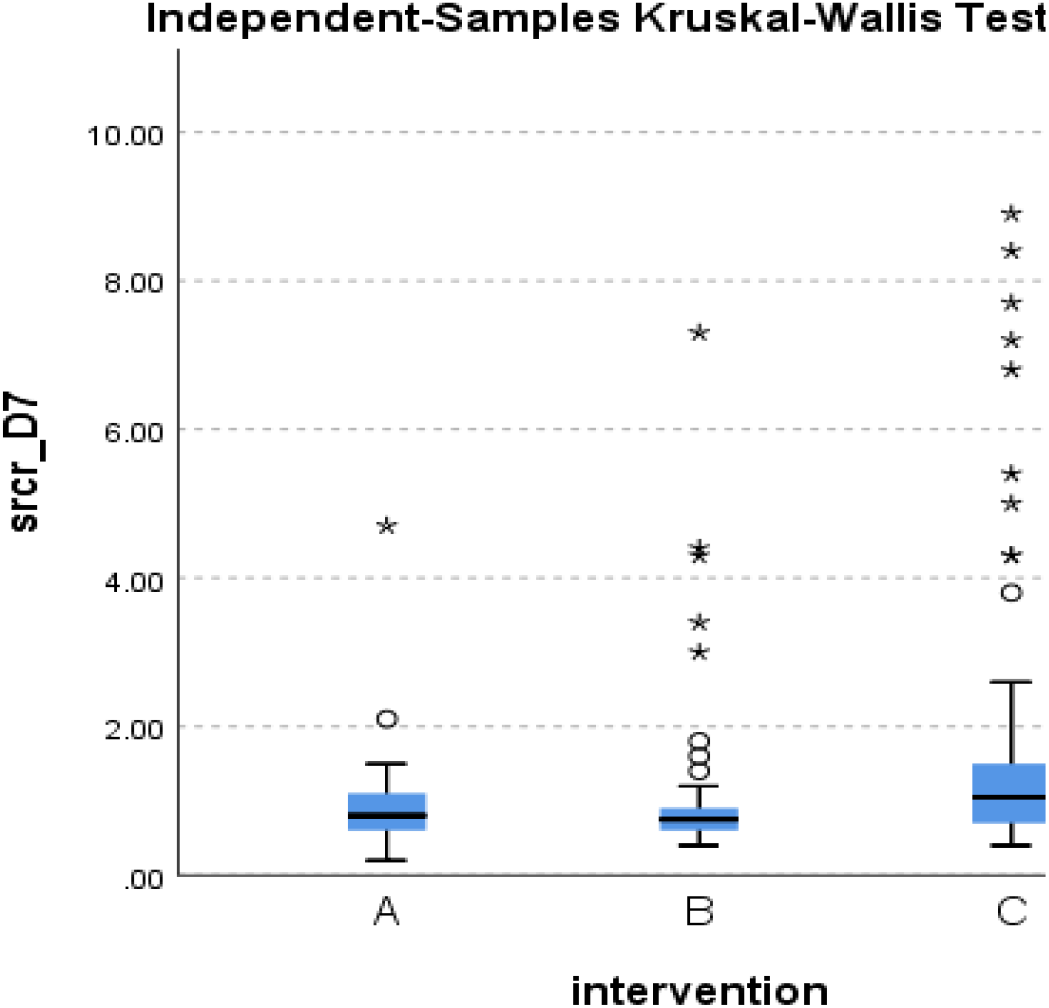
Distribution of S.Cr at day 7 across the three groups.

**Figure 33.**
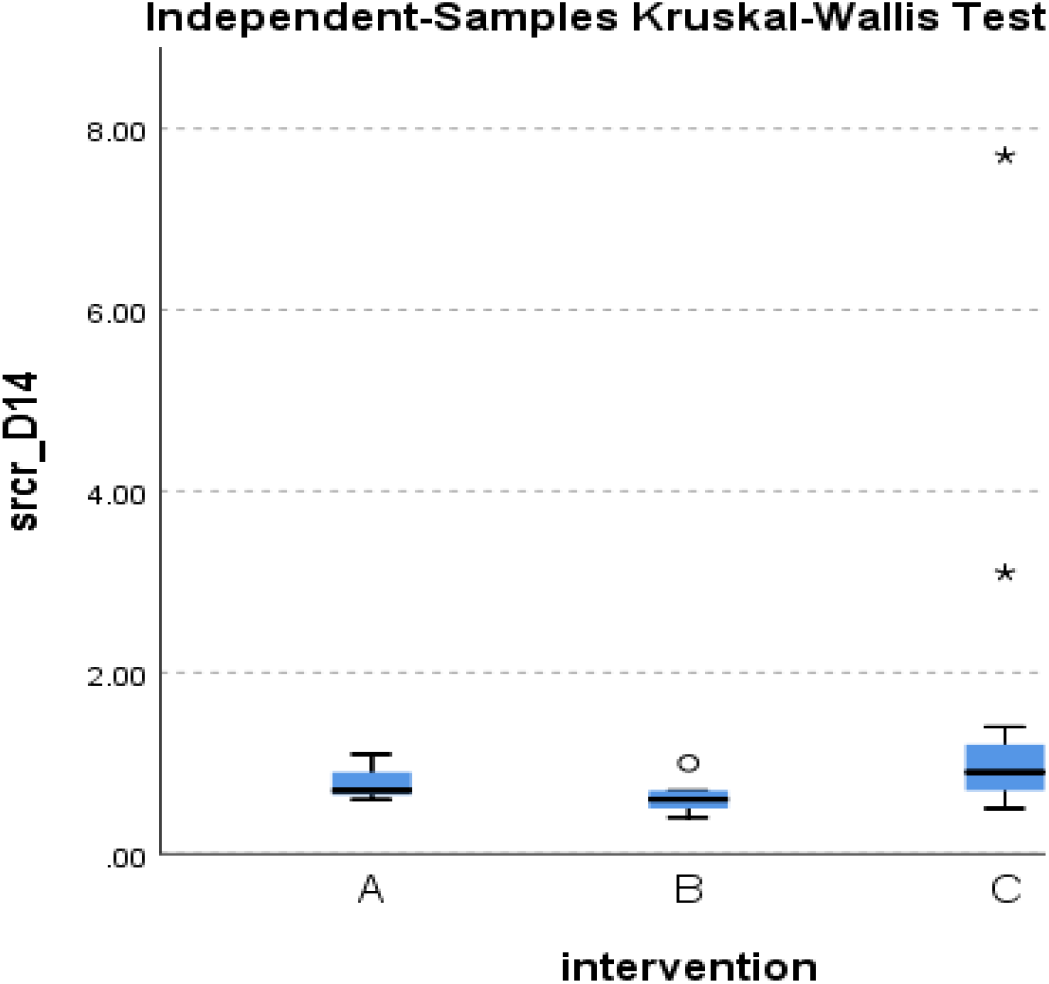
Distribution of S.Cr at day 14 across the three groups.

**Figure 34.**
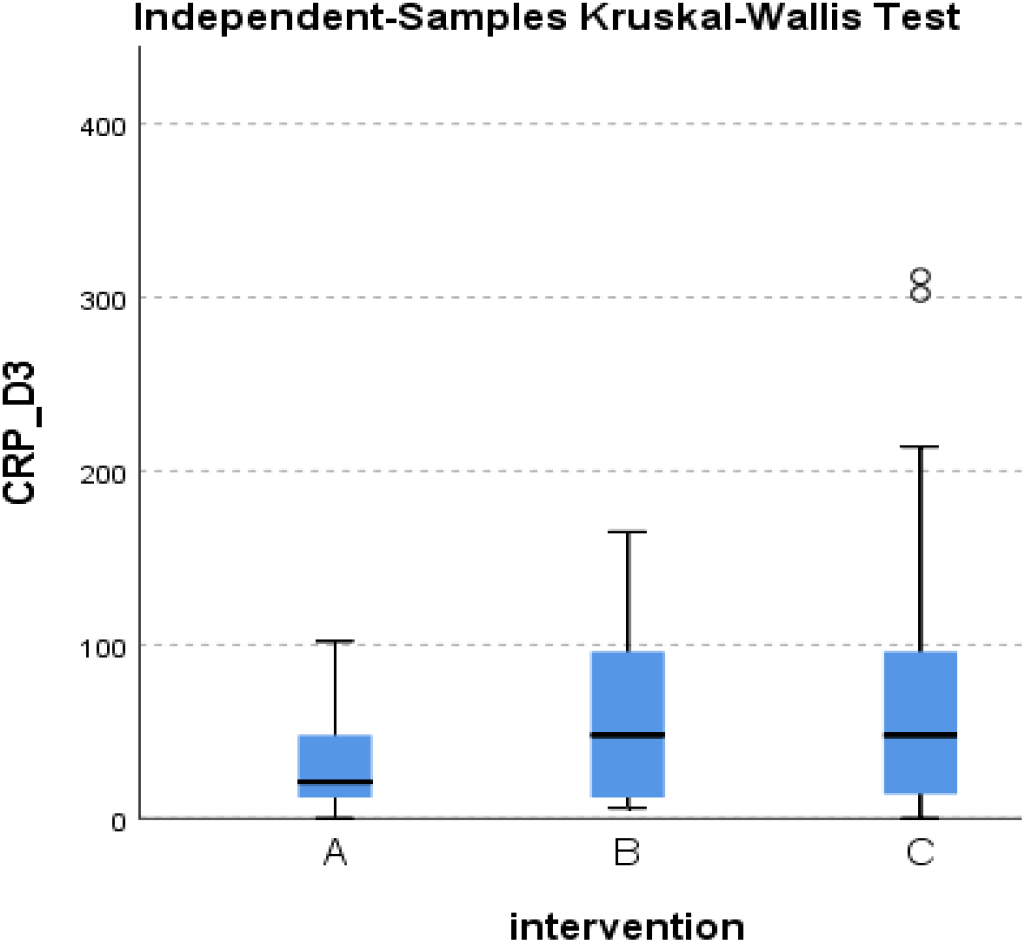
Distribution of CRP at day 3 across the three groups.

**Figure 35.**
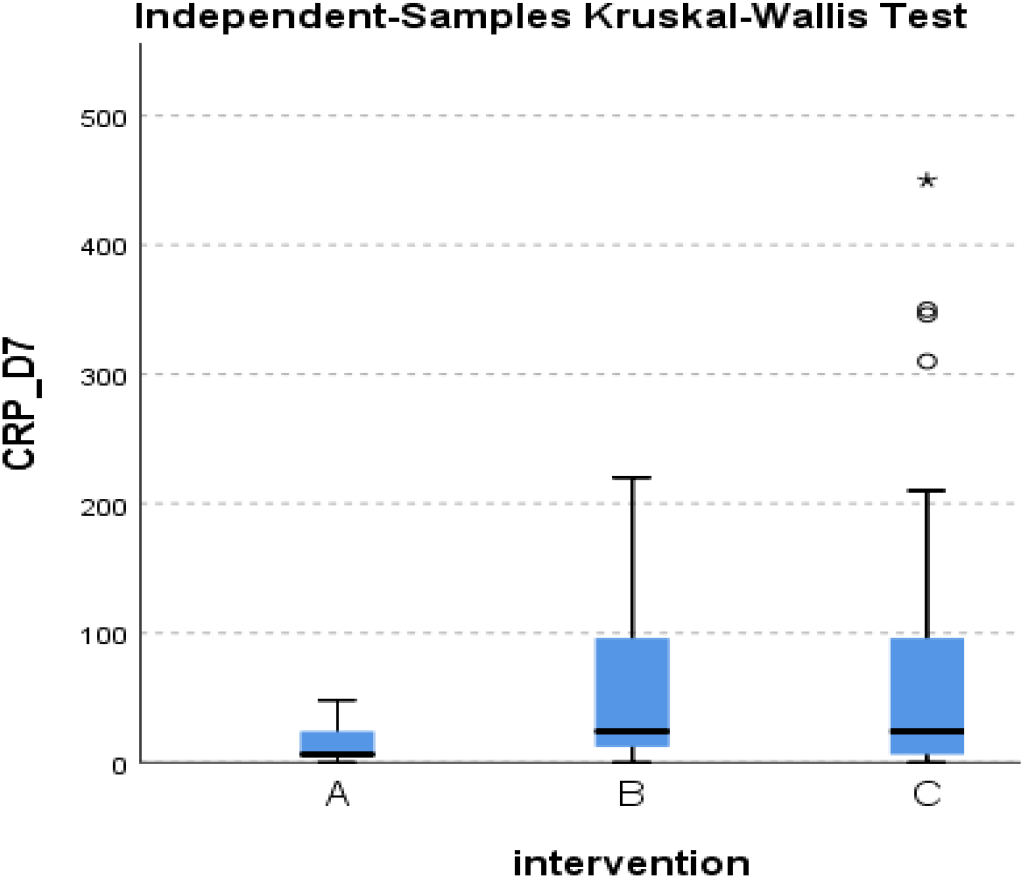
Distribution of CRP at day 7 across the three groups.

**Figure 36.**
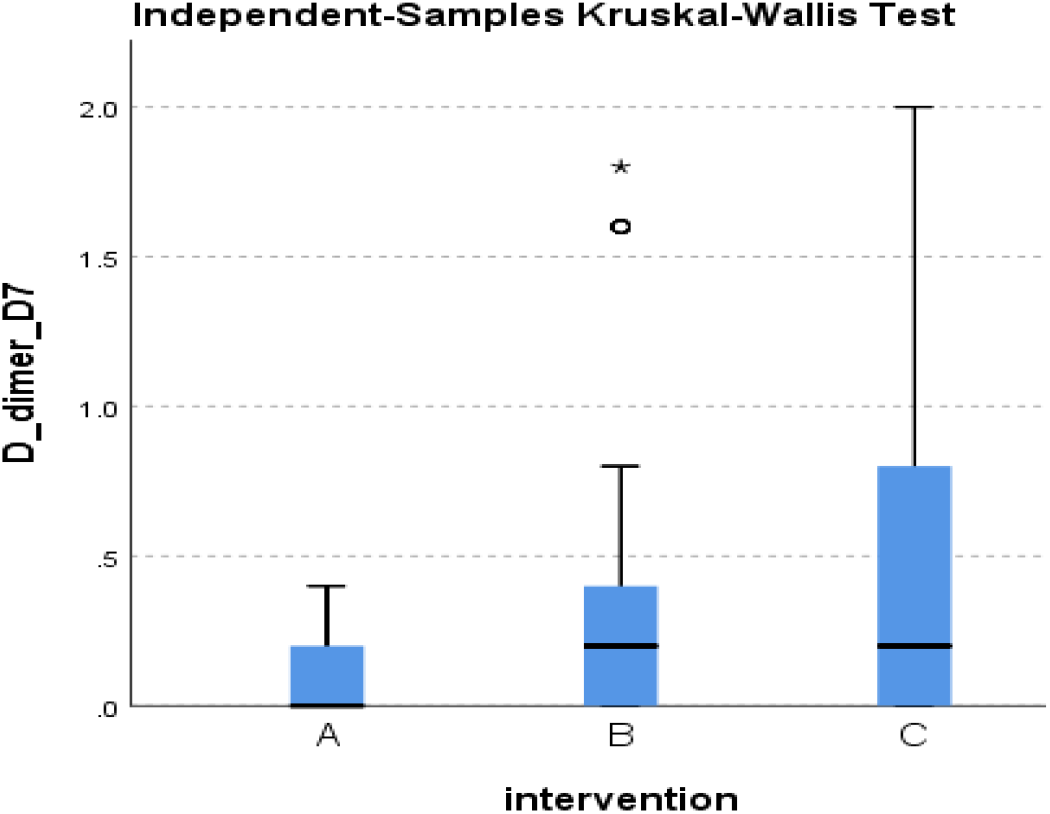
Distribution of D-dimer at day 7 across the three groups.

**Figure 37.**
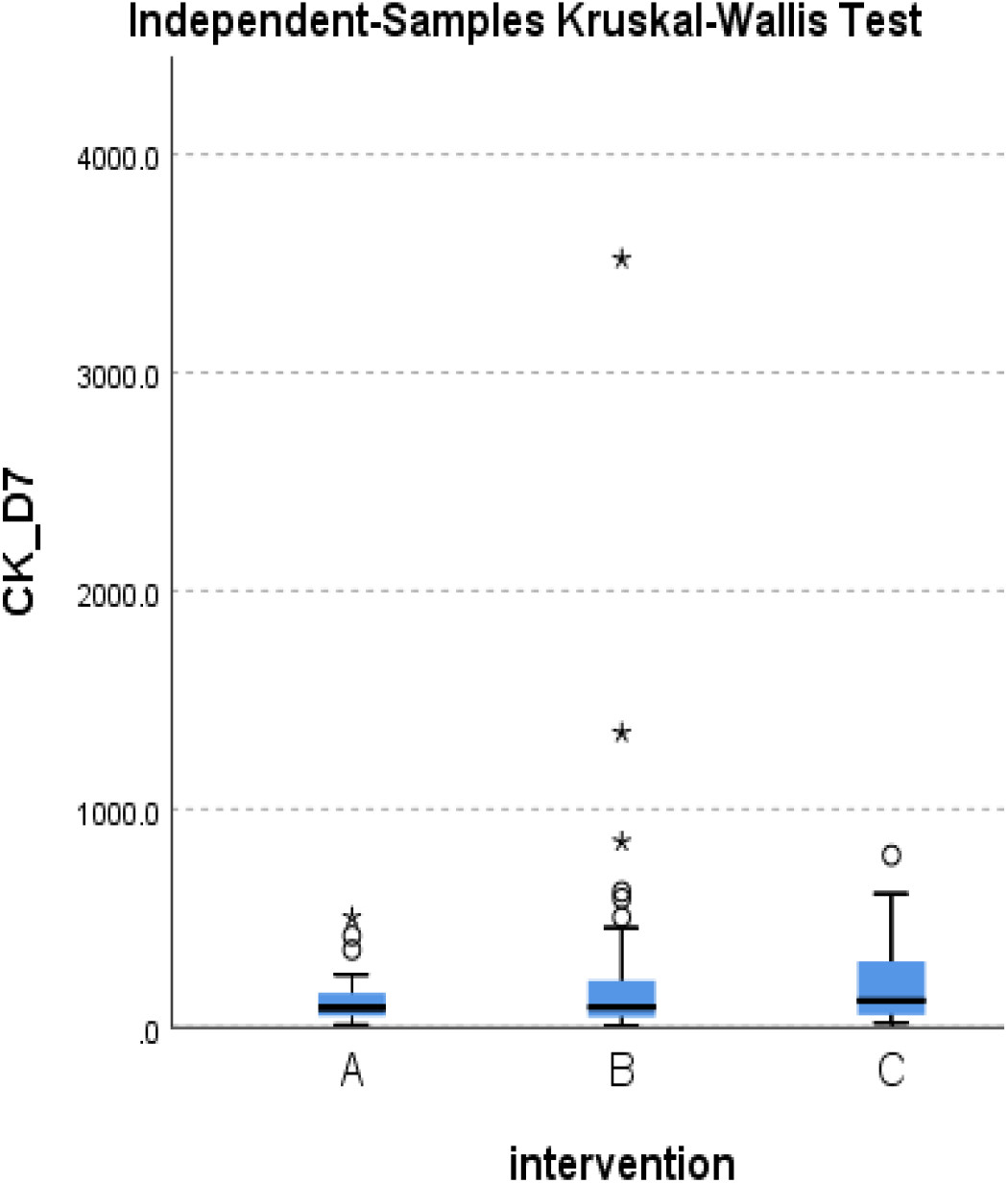
Distribution of CK at day 7 across the three groups.

**Figure 38.**
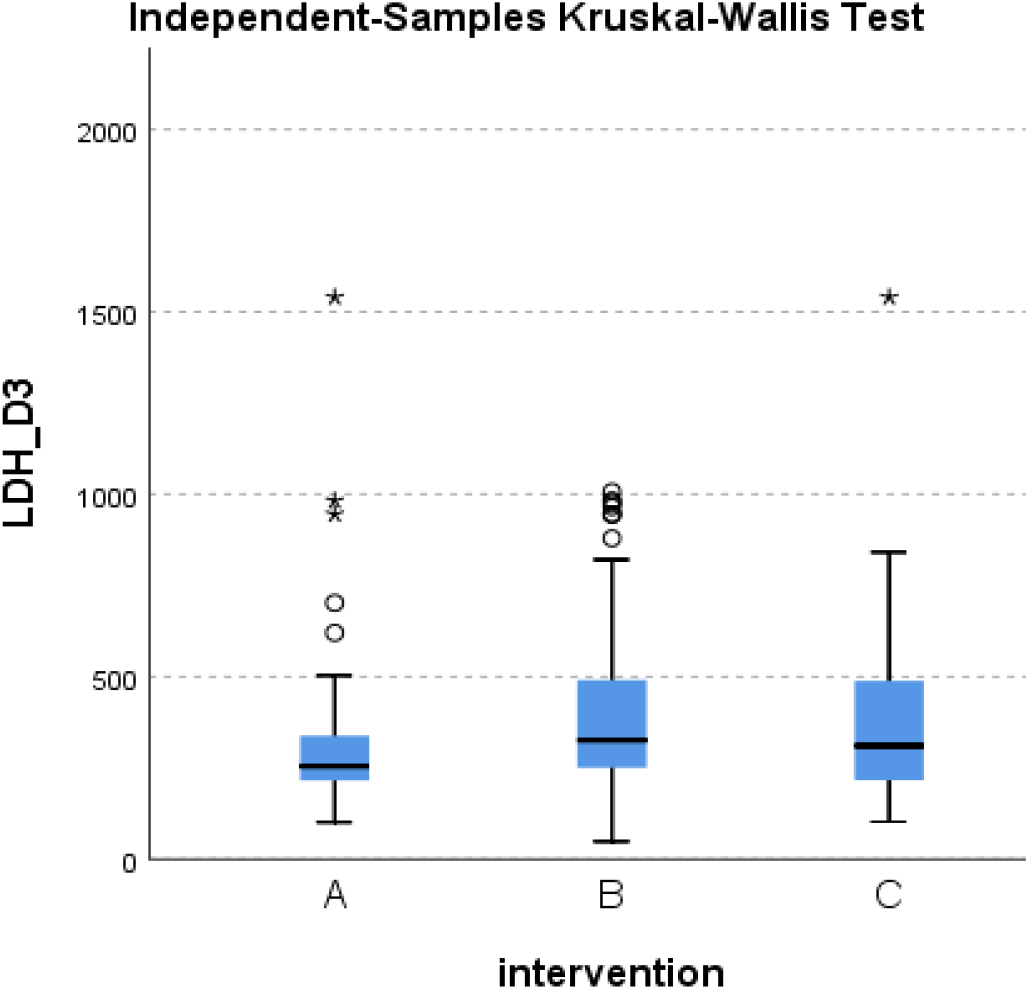
Distribution of LDH at day 3 across the three groups.

**Figure 39.**
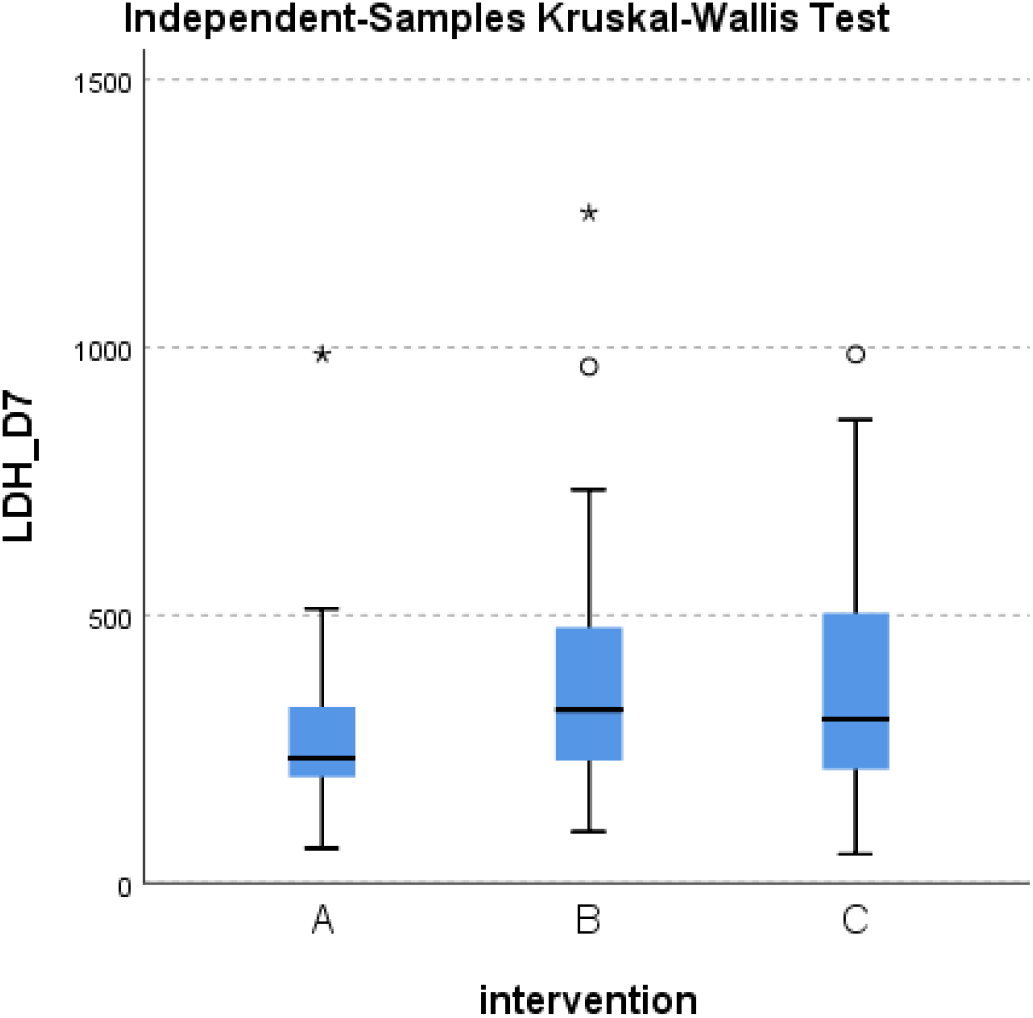
Distribution of LDH at day 7 across the three groups.

**Figure 40.**
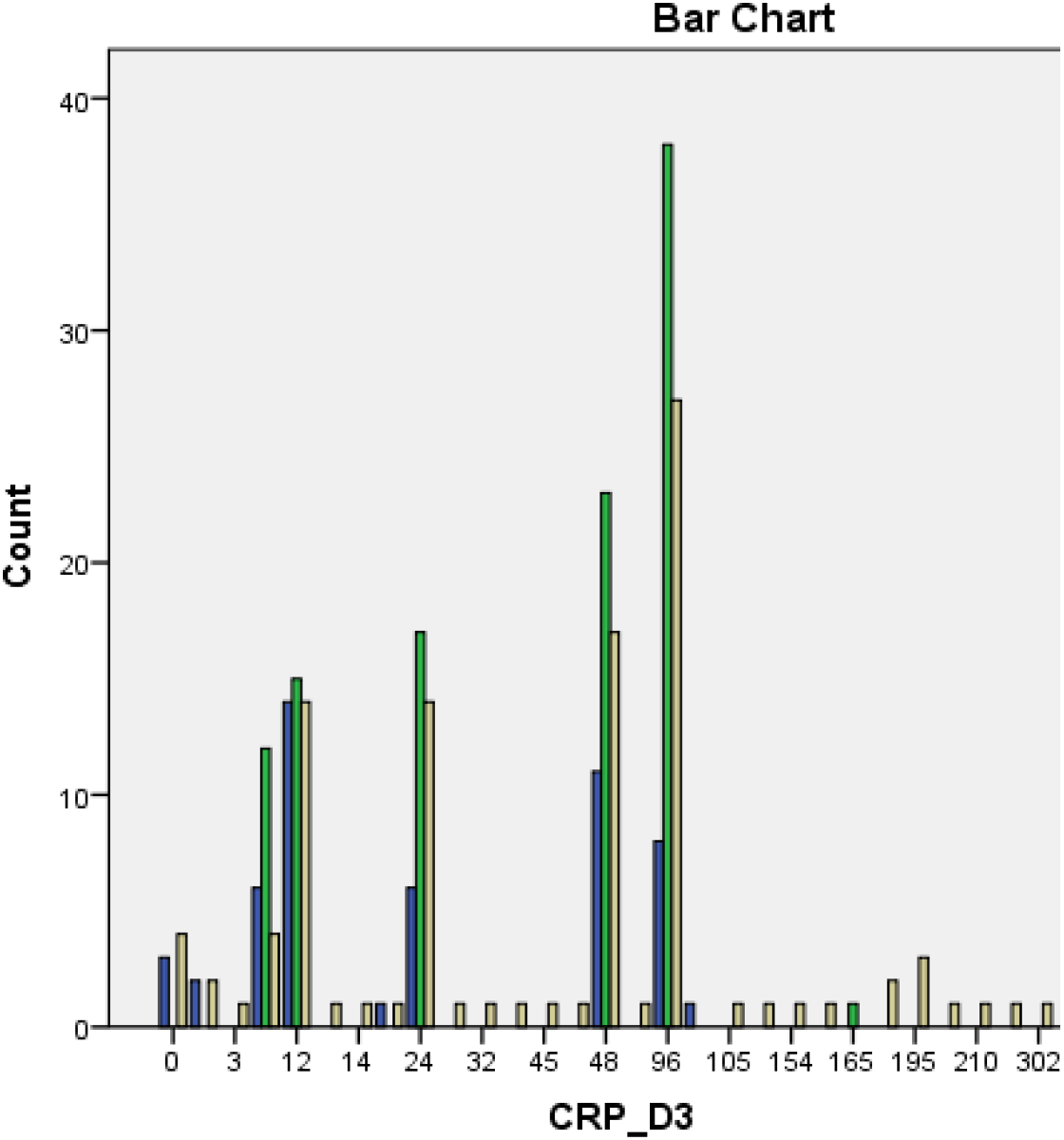
Frequency of CRP at day 3 across the three groups.

**Figure 41.**
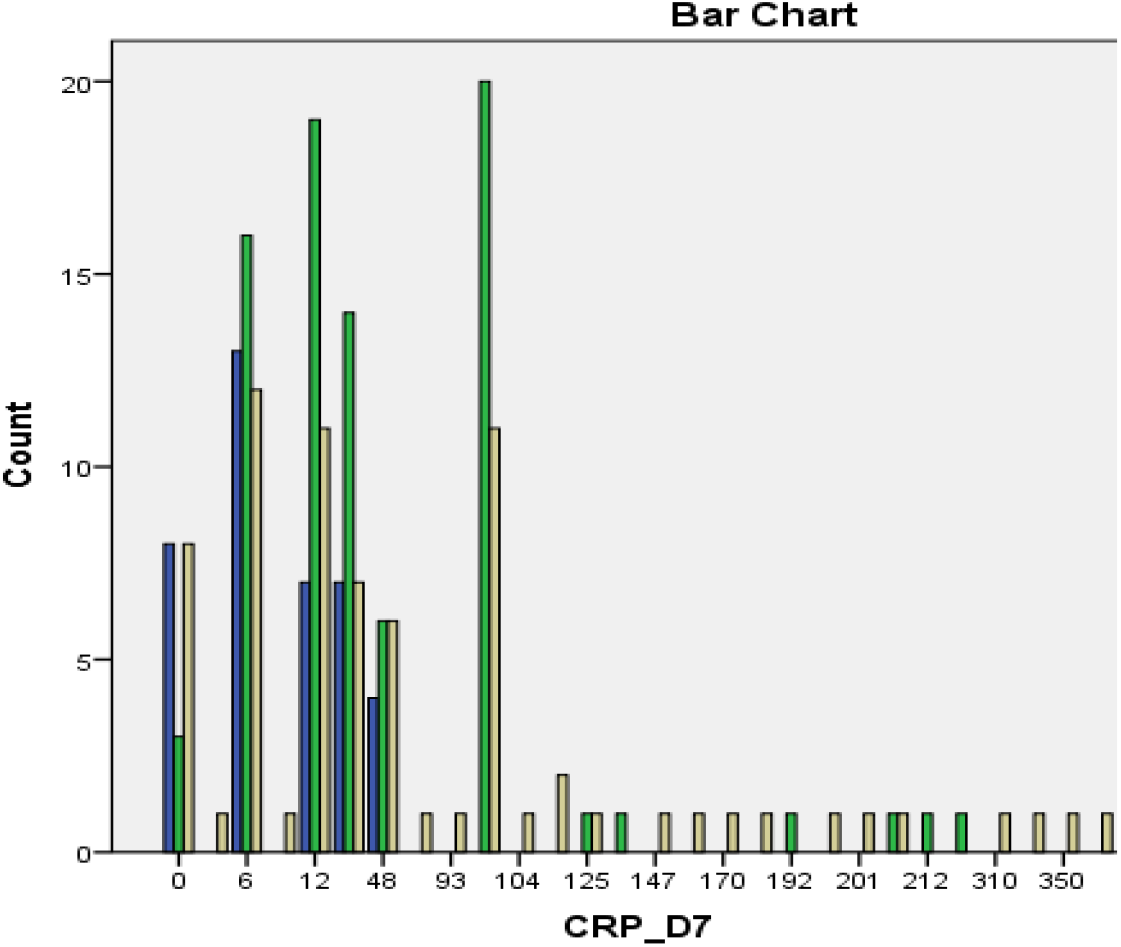
Frequency of CRP at day 7 across the three groups.

**Figure 42.**
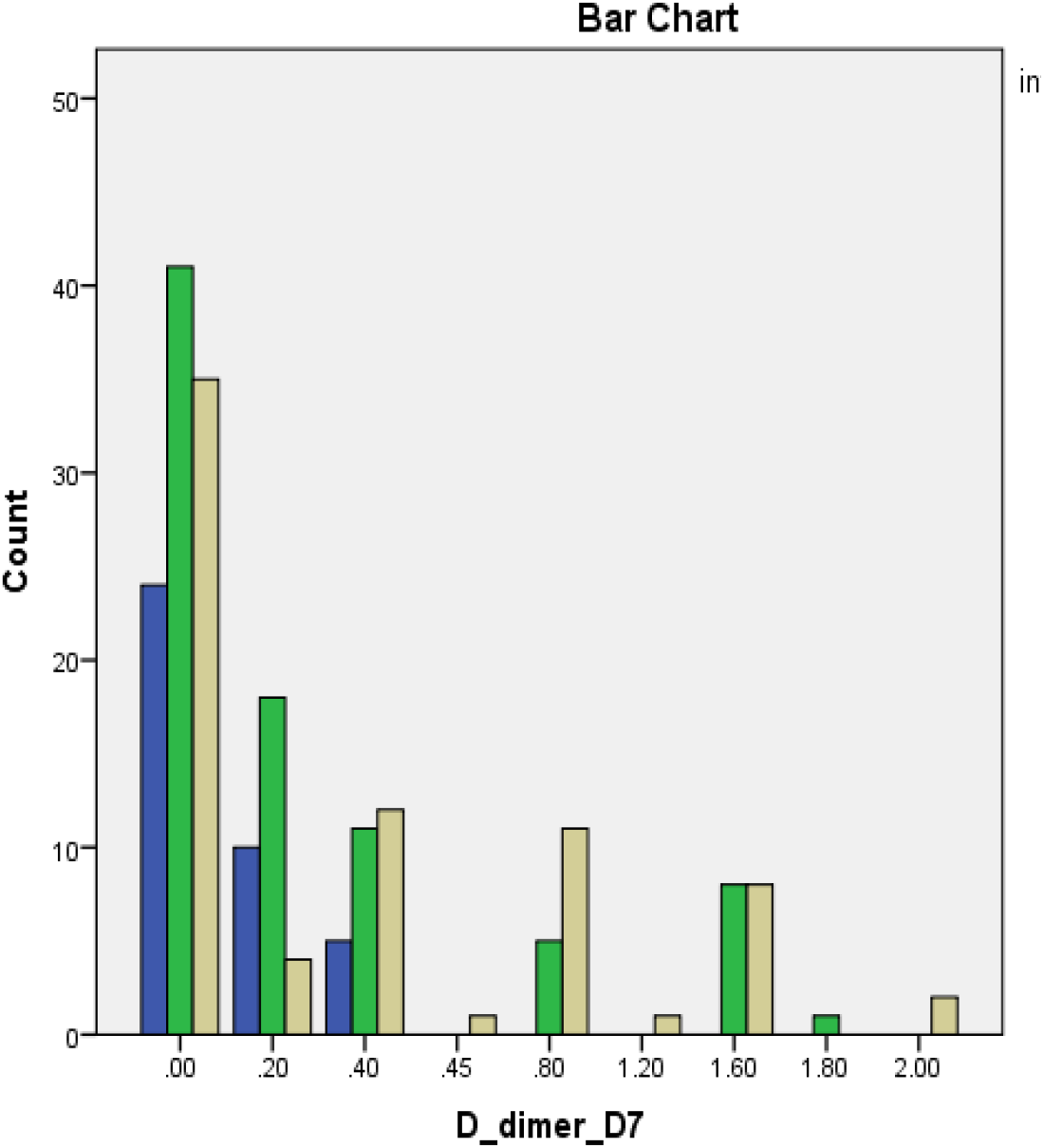
Frequency of D-dimer at day 7 across the three groups.

**Figure 43.**
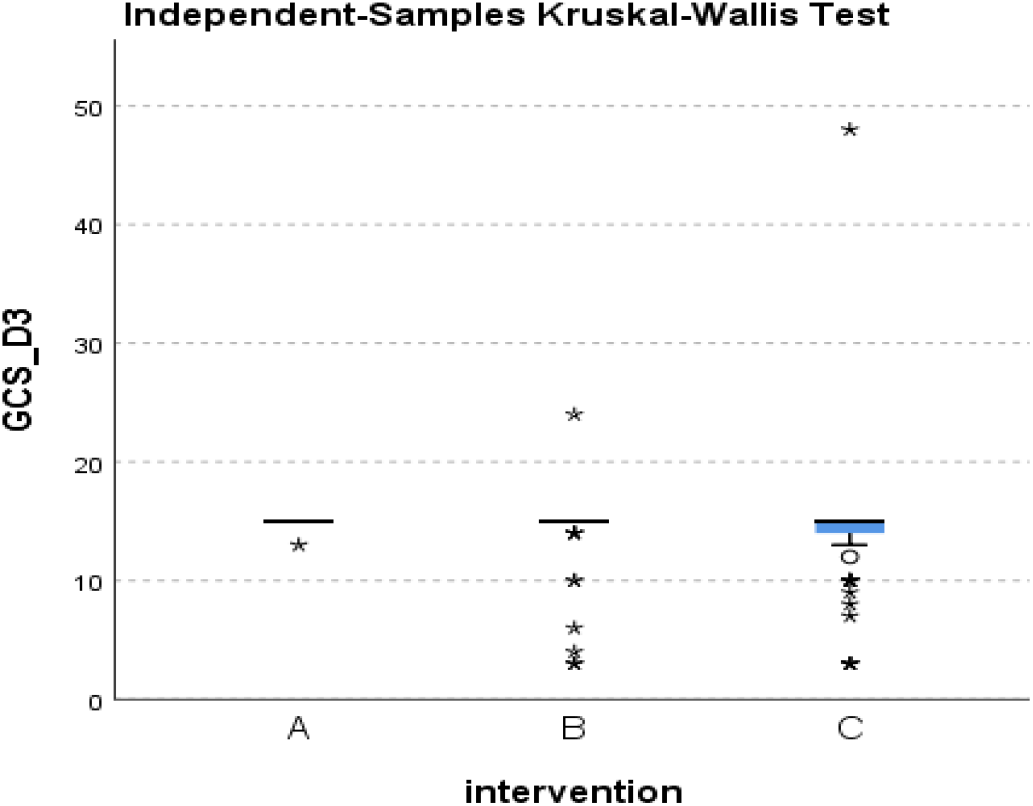
Distribution of GCS at day 3 across the three groups.

**Figure 44.**
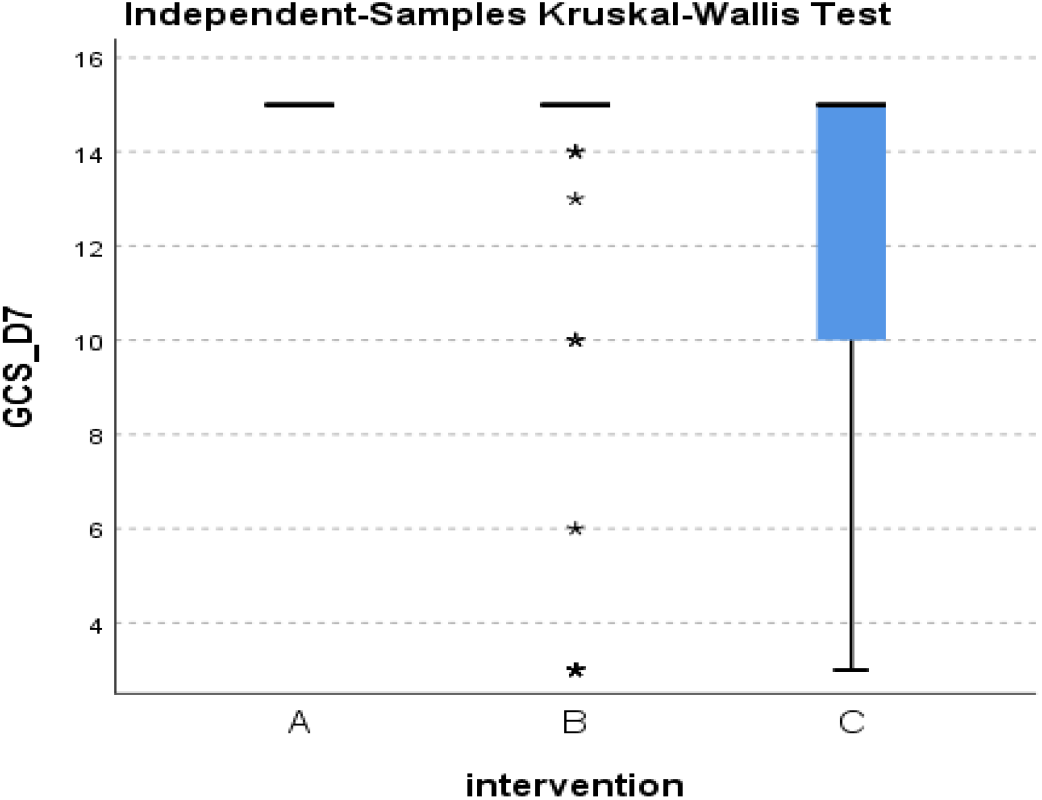
Distribution of GCS at day 7 across the three groups.

**Figure 45.**
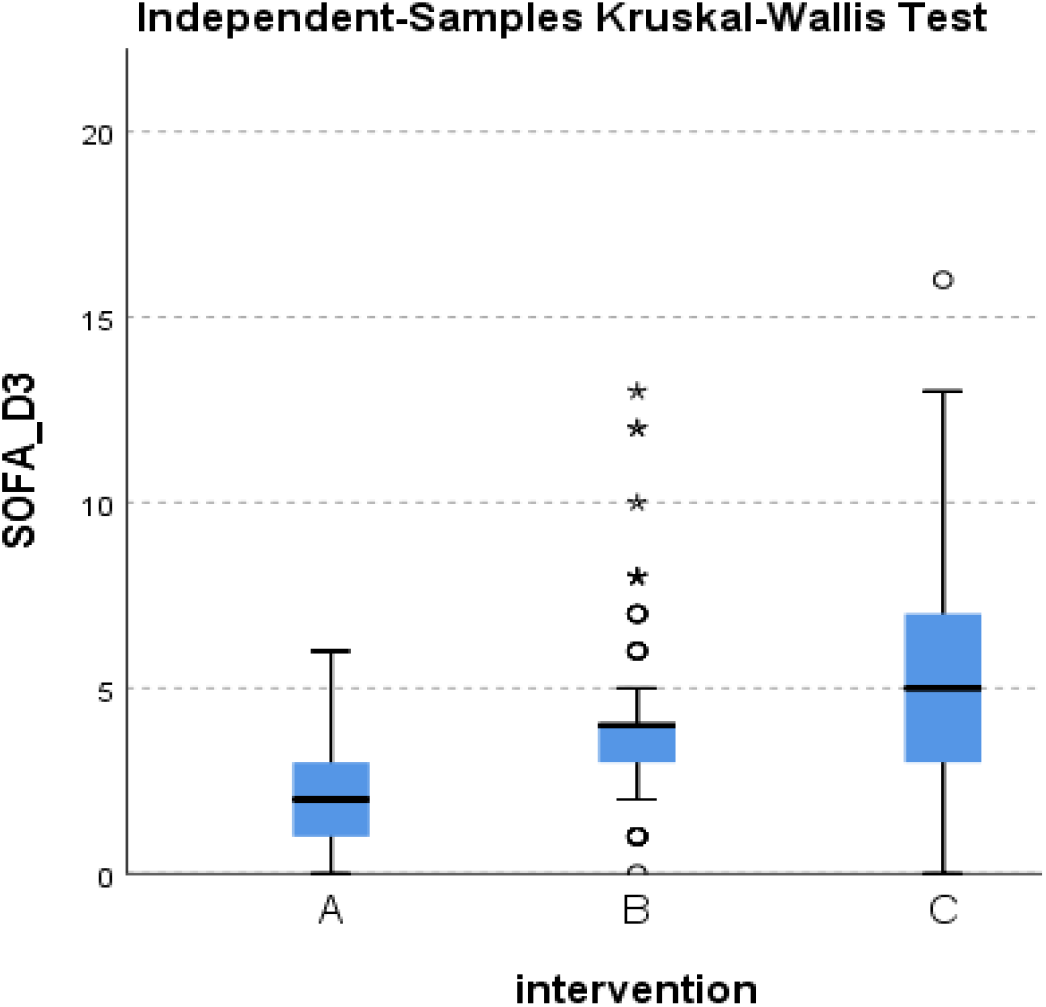
Distribution of SOFA at day 3 across the three groups.

**Figure 46.**
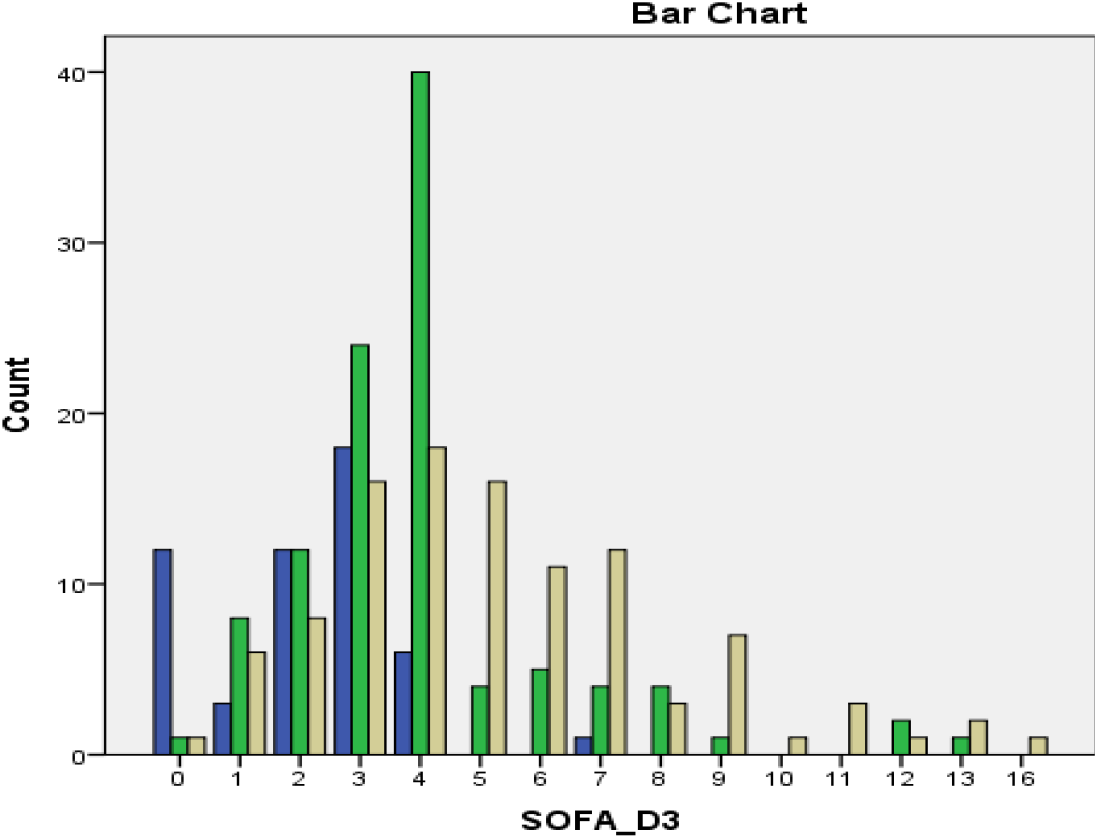
Frequency of SOFA score at day 3 across the three groups.

**Figure 47.**
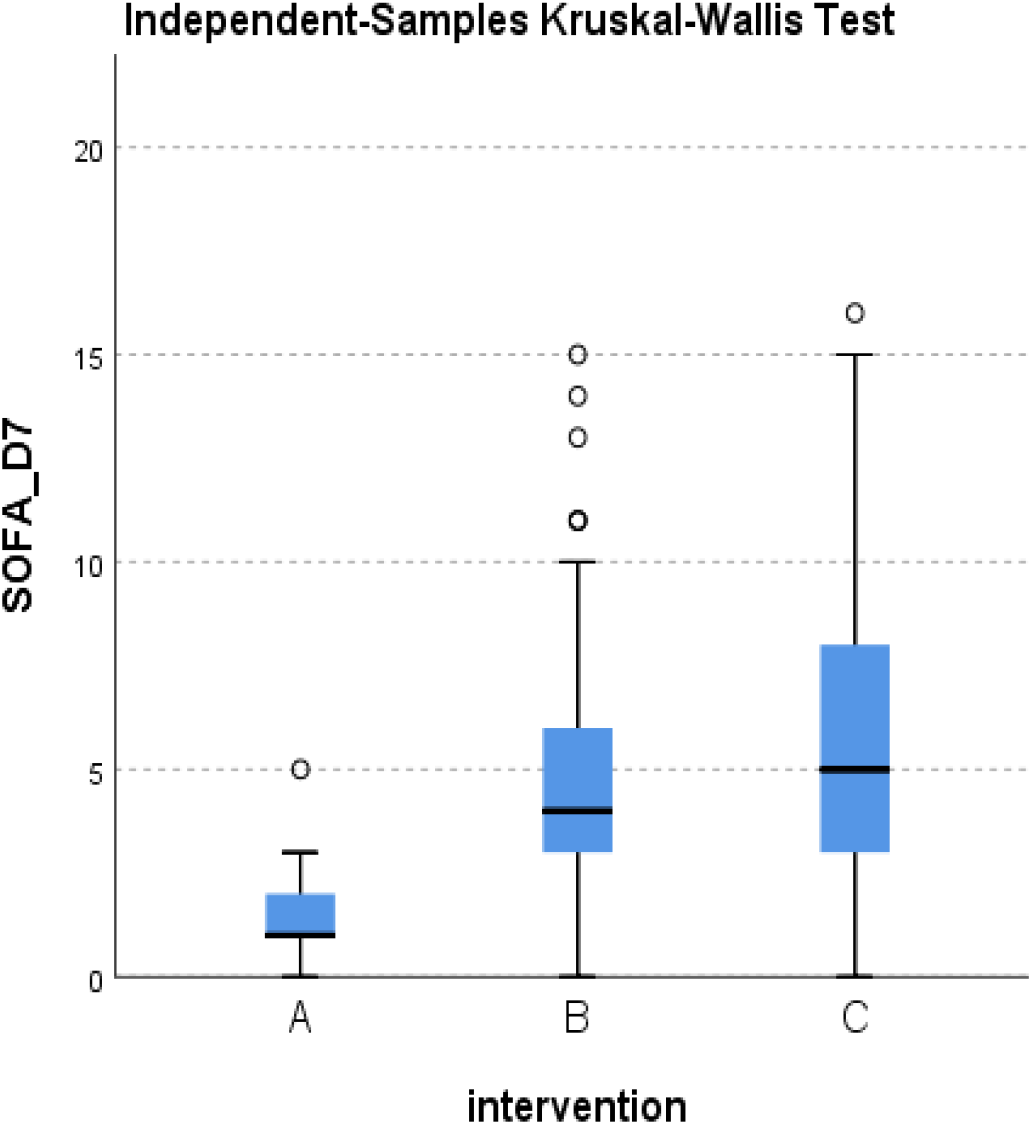
Distribution of SOFA at day 7 across the three groups.

**Figure 48.**
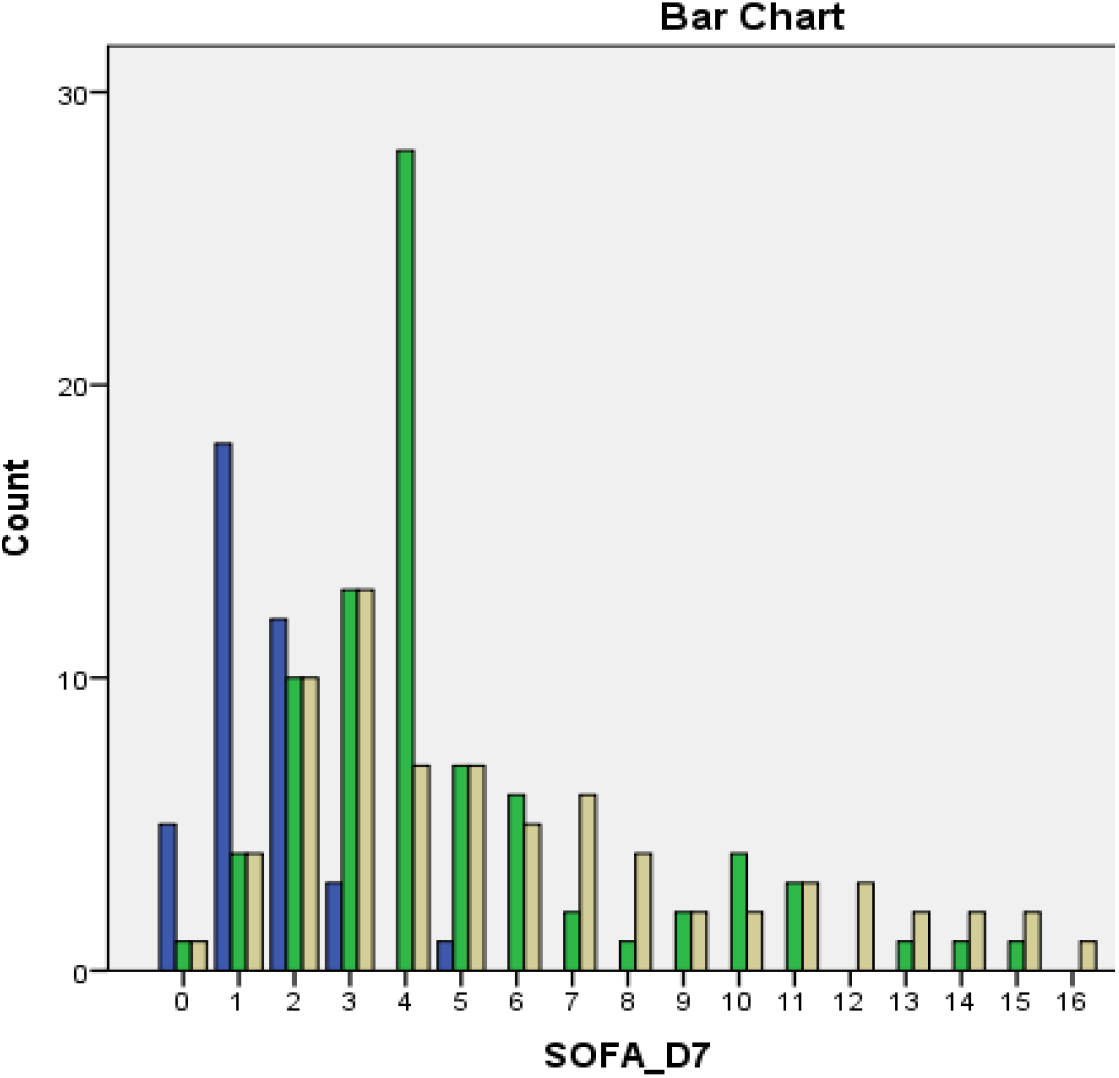
Frequency of SOFA score at day 7 across the three groups.

**Figure 49.**
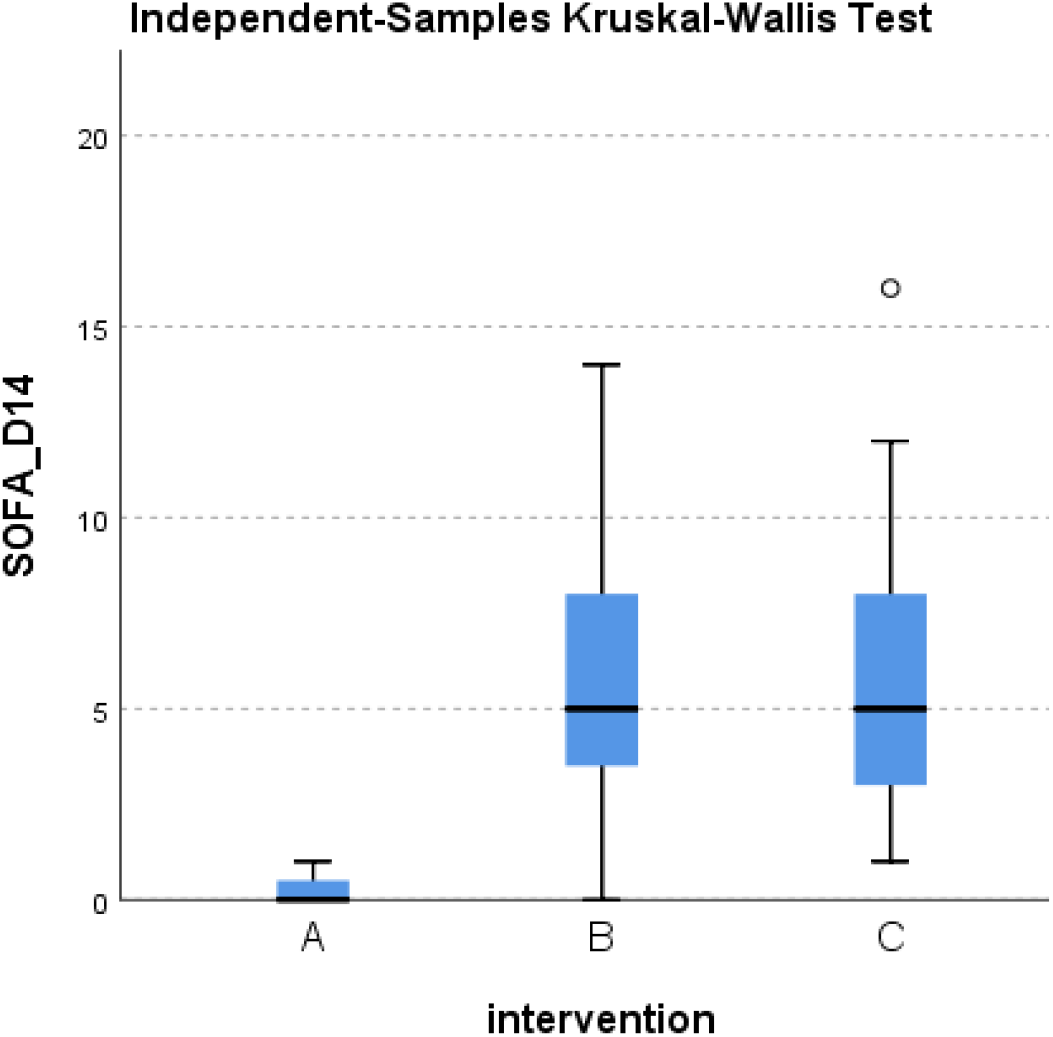
Distribution of SOFA at day 14 across the three groups.

**Figure 50.**
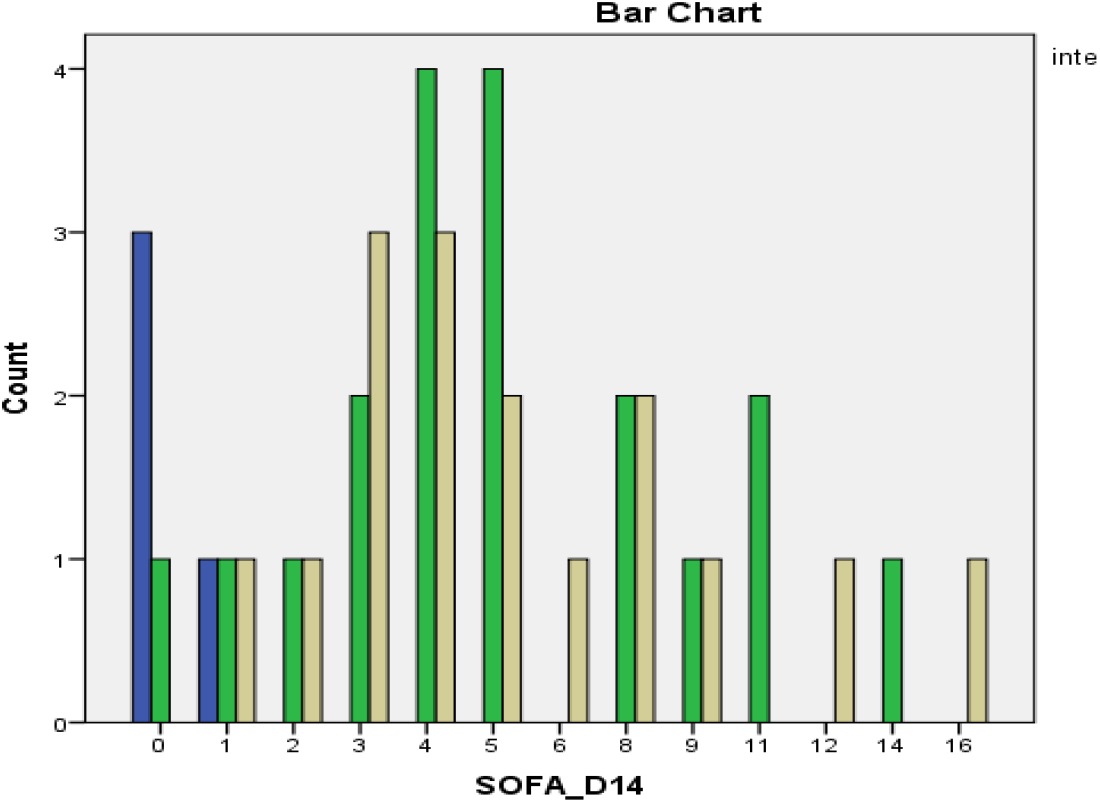
Frequency of SOFA score at day 14 across the three groups.

**Figure 51.**
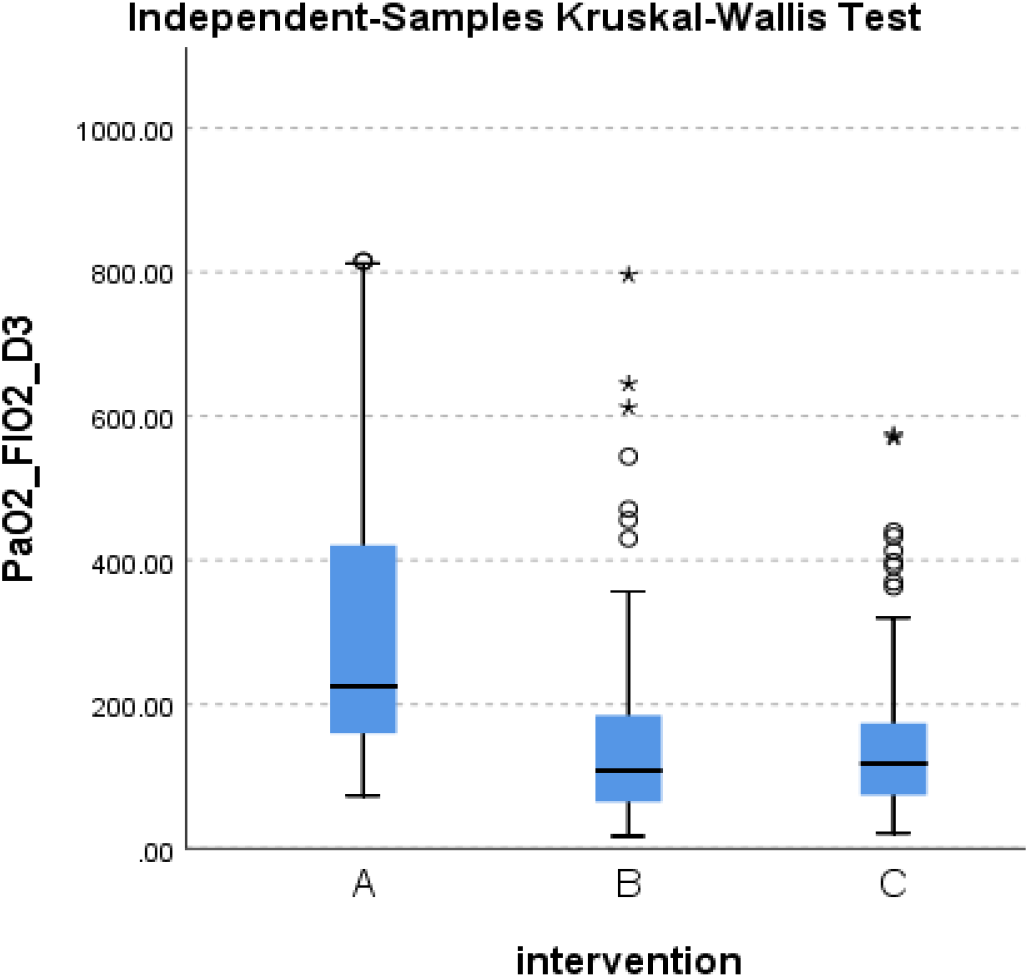
Distribution of PaO2/FiO2 at day 3 across the three groups.

**Figure 52.**
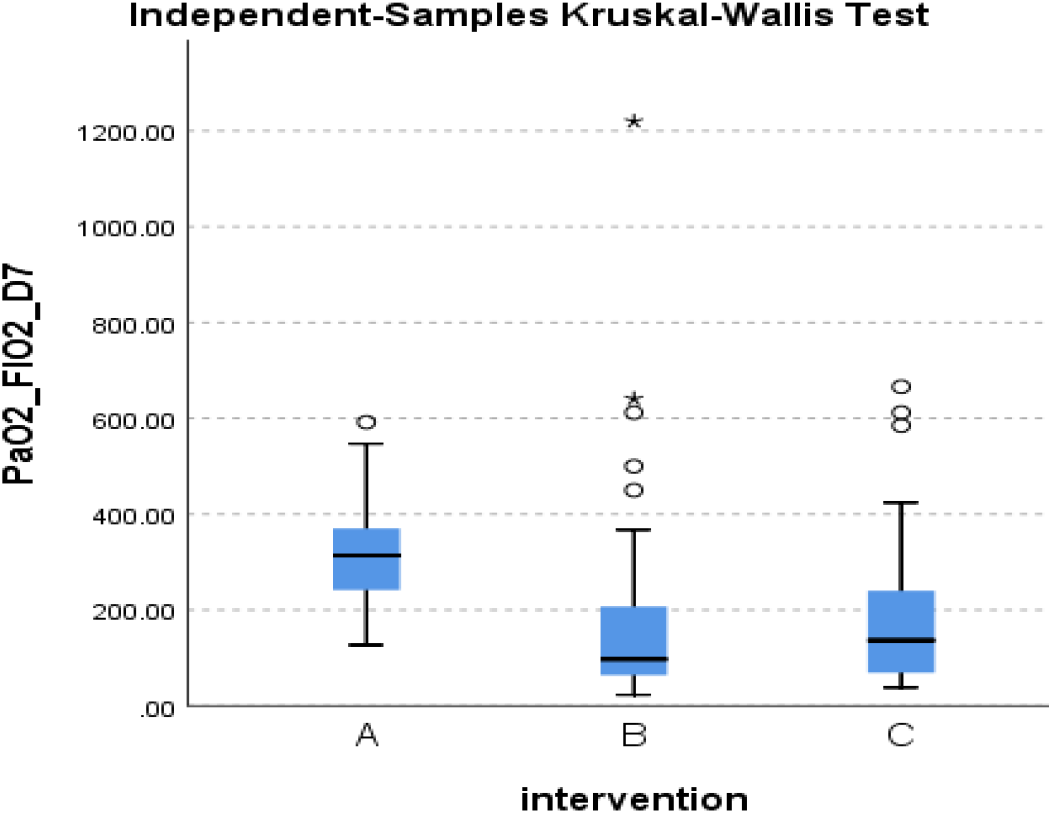
Distribution of PaO2/FiO2 at day 7 across the three groups.

**Figure 53.**
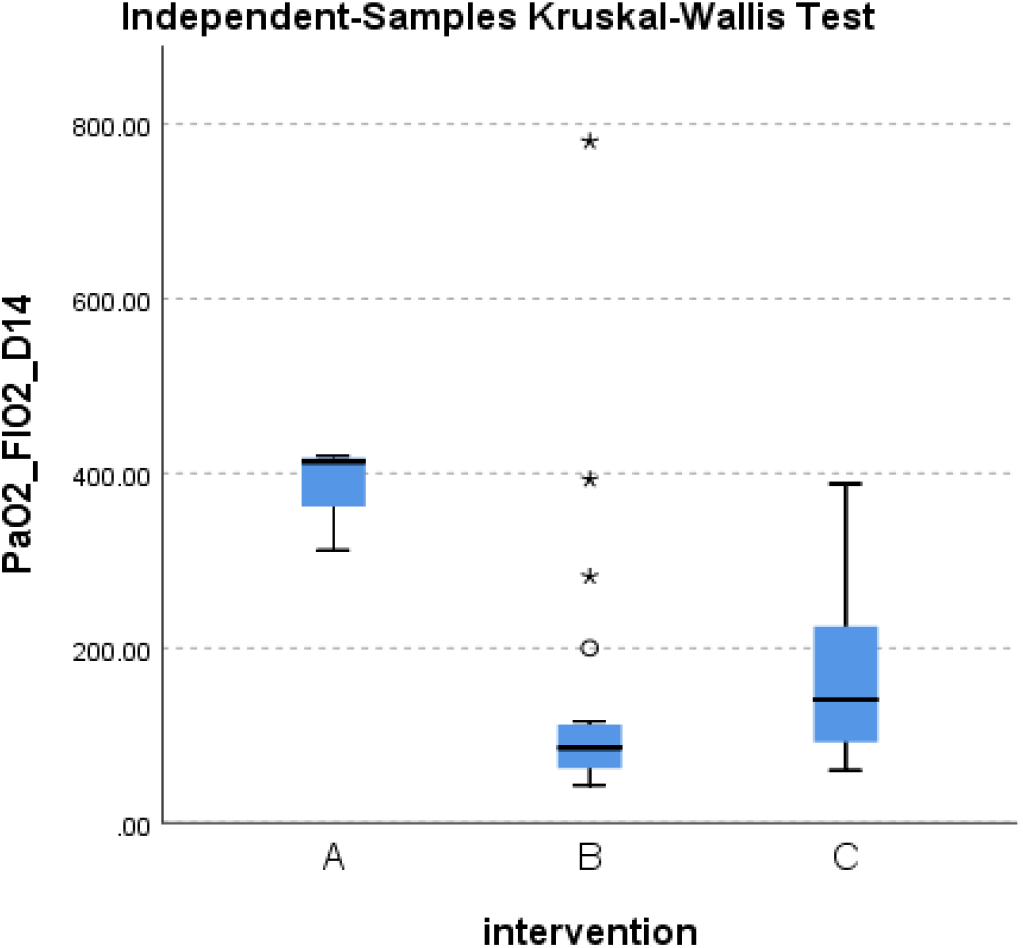
Distribution of PaO2/FiO2 at day 14 across the three groups.

**Figure 54.**
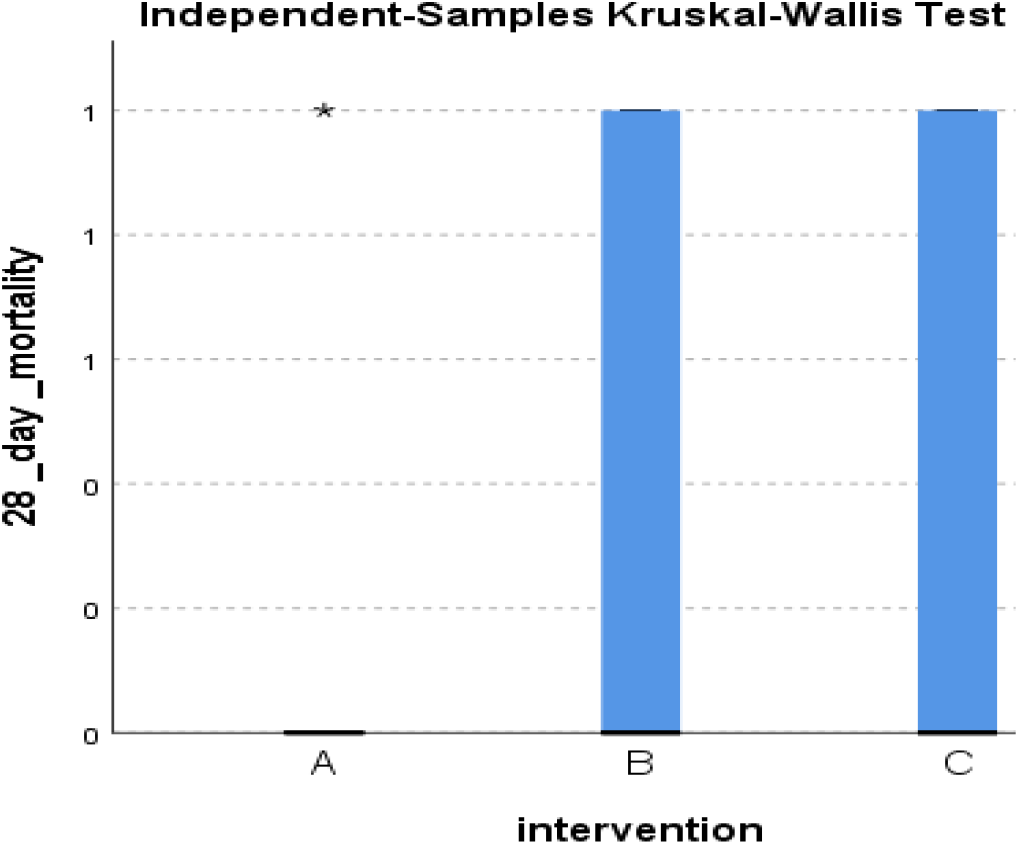
Distribution of 28-day mortality across the three groups.

**Figure 55.**
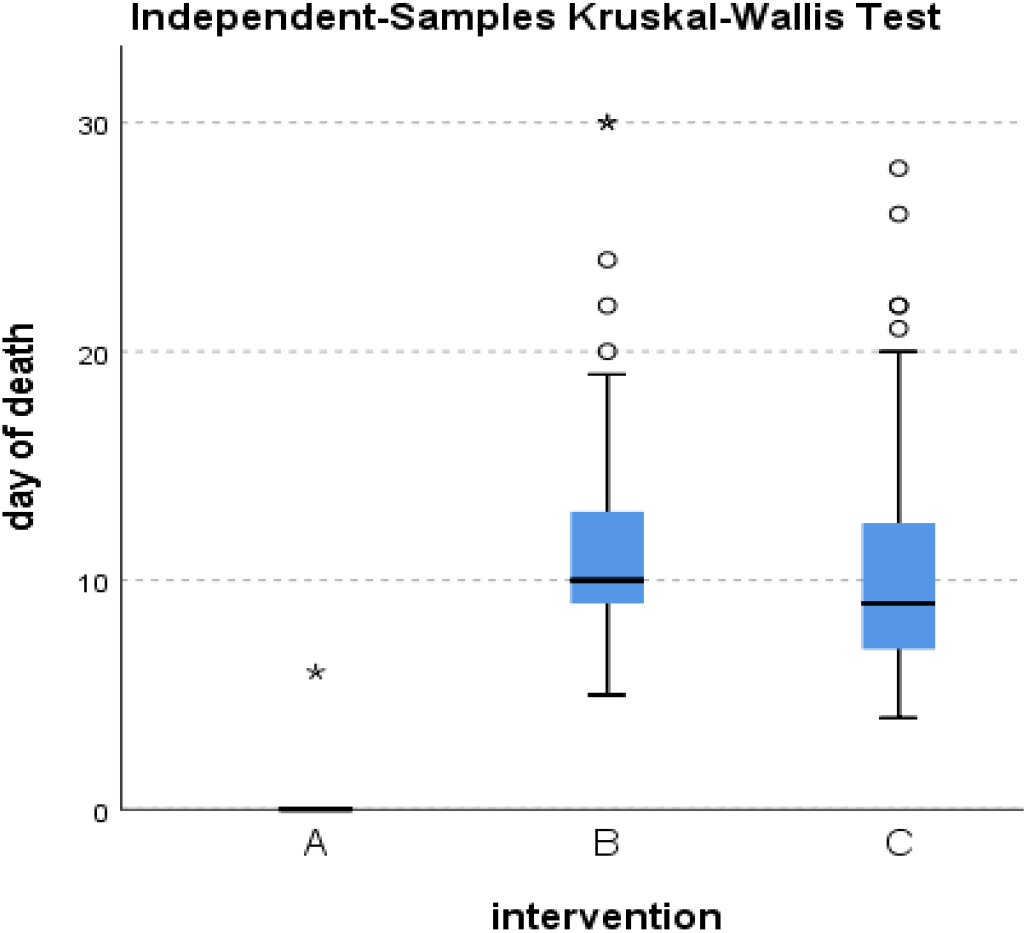
Distribution of day of death across the three groups.

**Figure 56.**
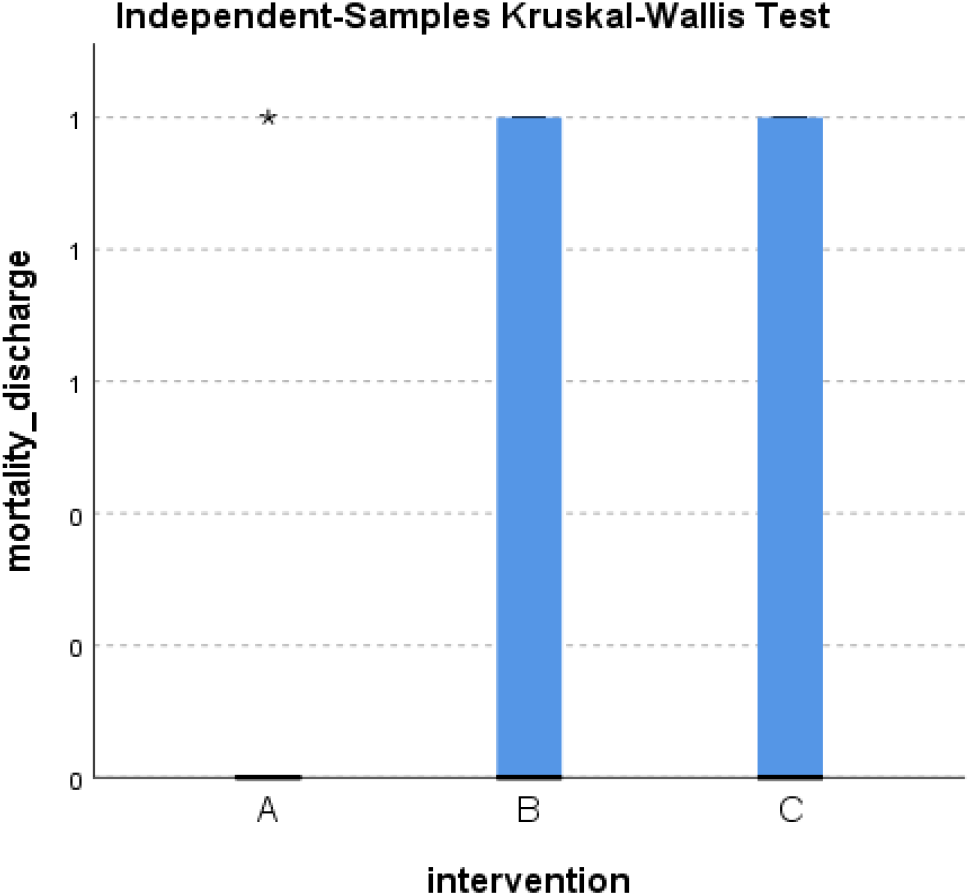
Distribution of mortality at discharge across the three groups.

**Figure 57.**
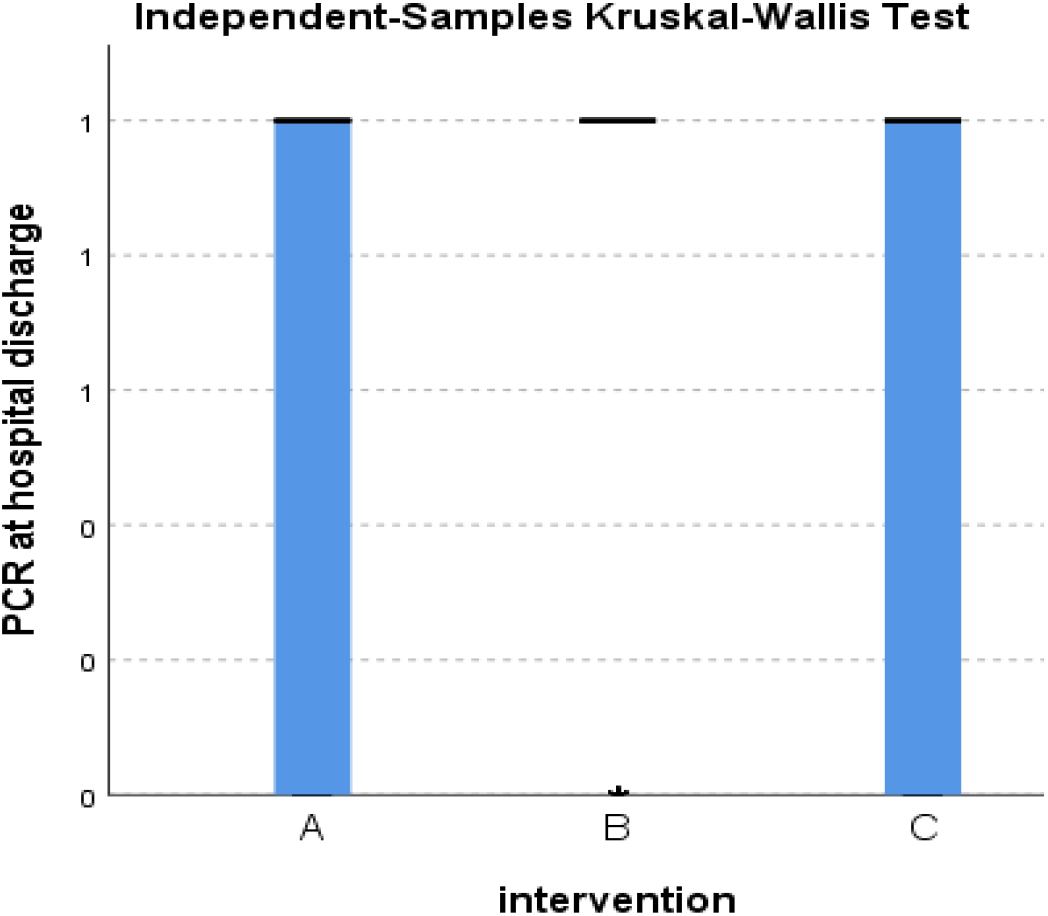
Distribution of PCR results at discharge across the three groups.

**Figure 58.**
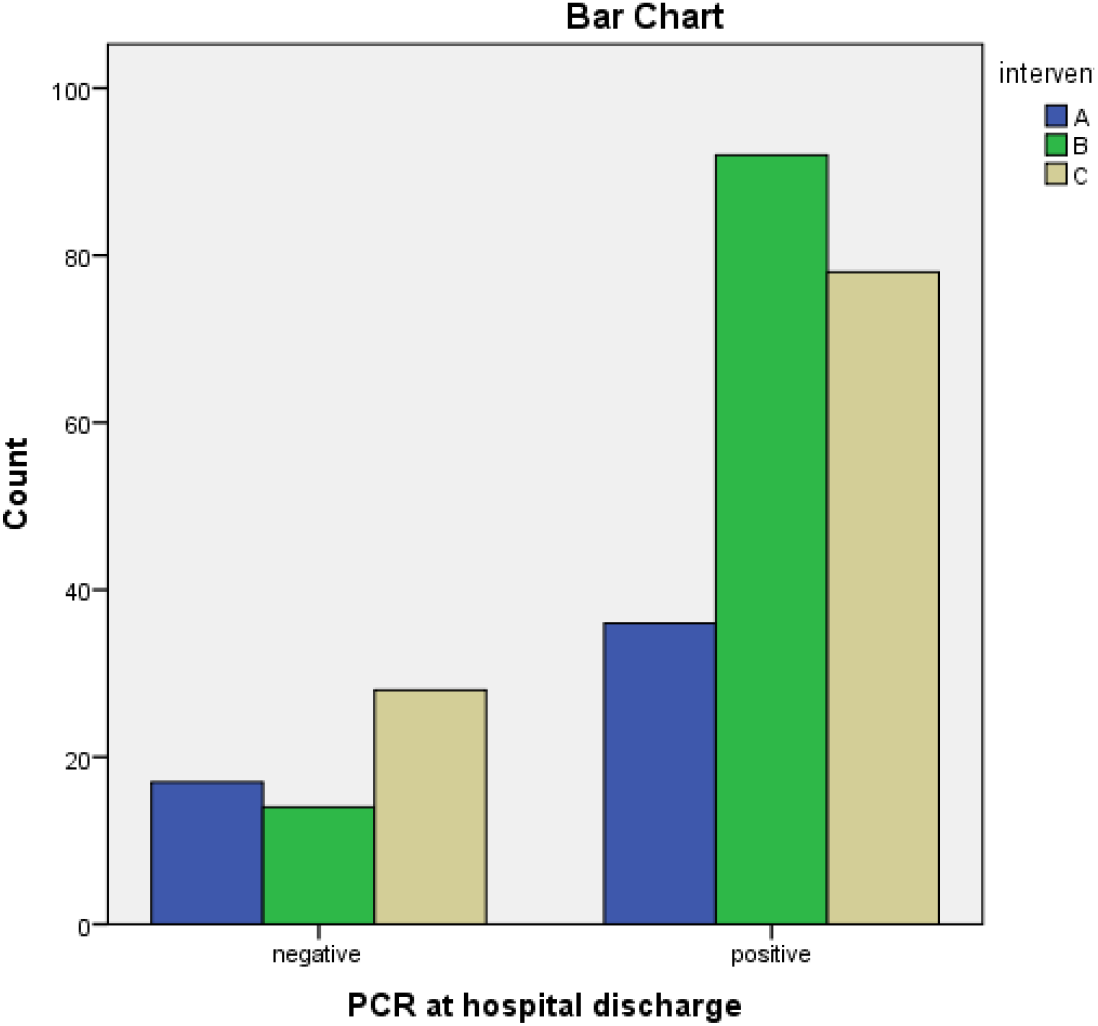
Frequency of PCR results at discharge across the three groups.

**Figure 59.**
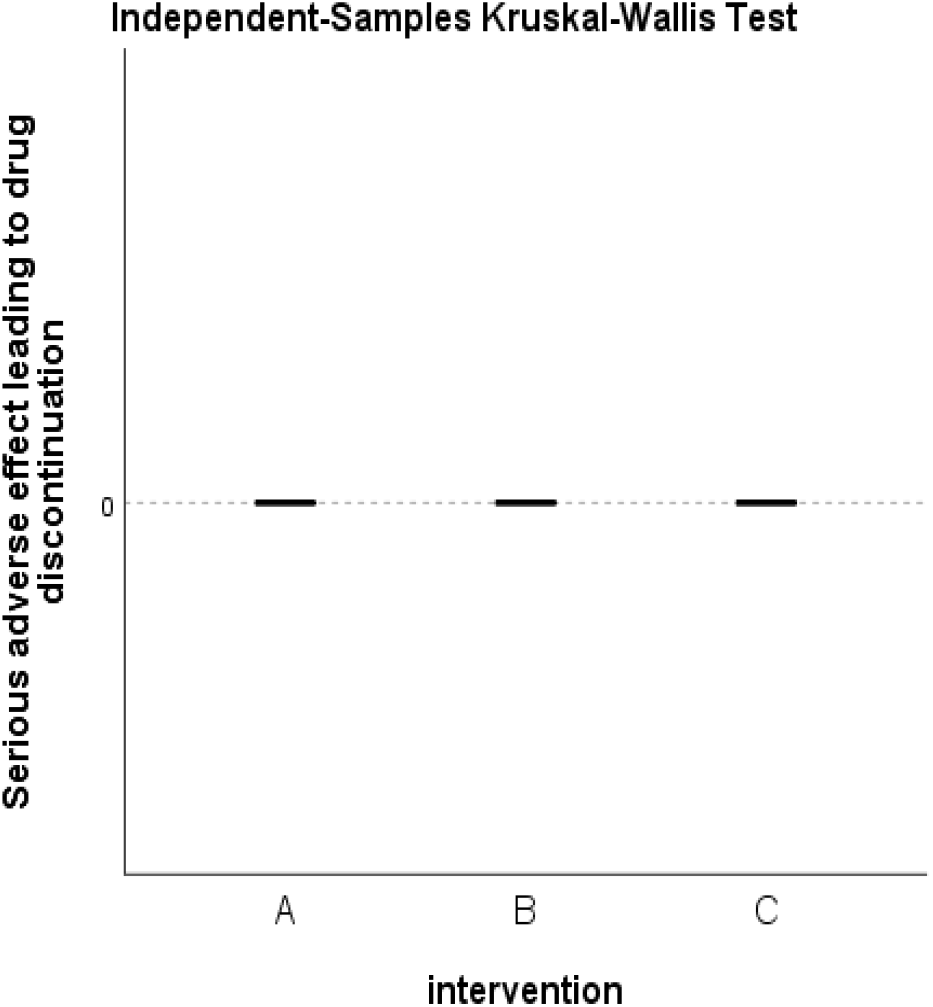
Distribution of serious adverse event incidence across the three groups.

**Figure 60.**
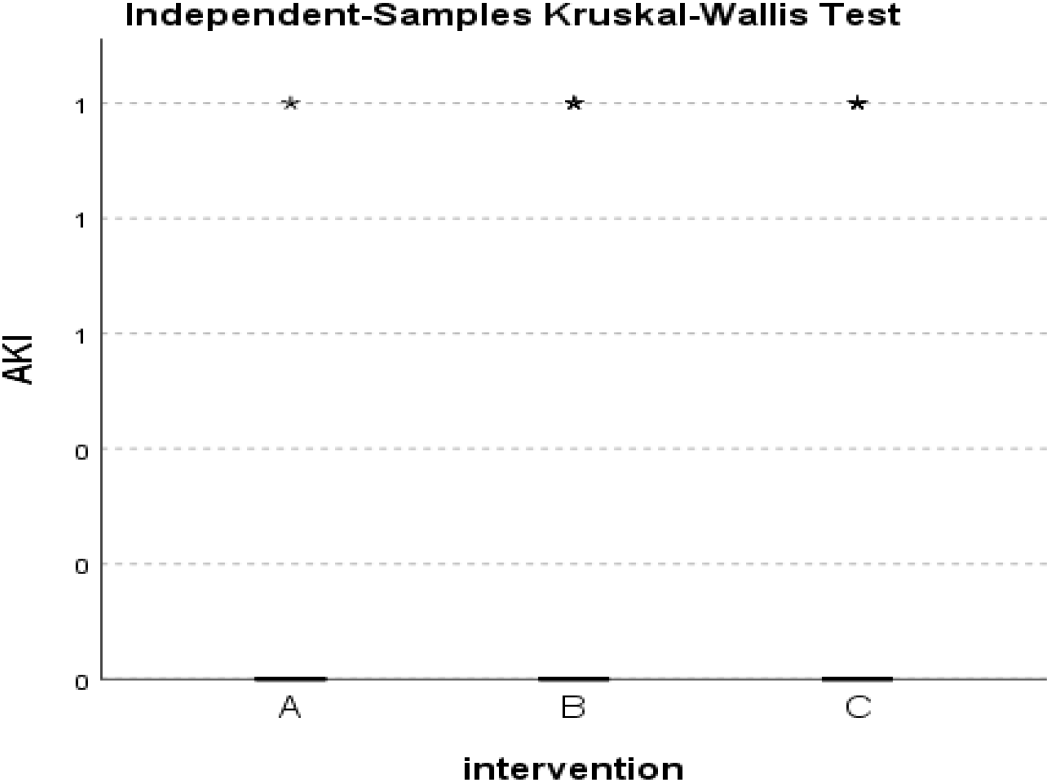
Distribution of AKI incidence across the three groups.

**Figure 61.**
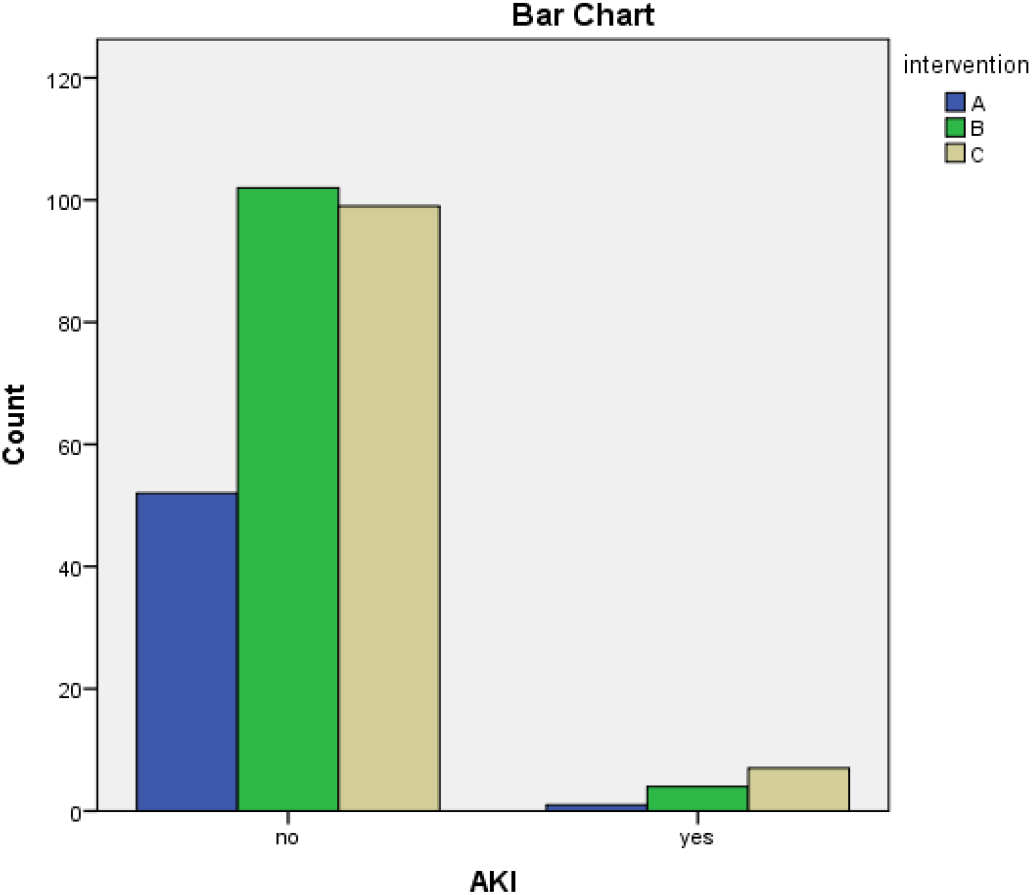
Frequency of AKI incidence across the three groups.

**Figure 62.**
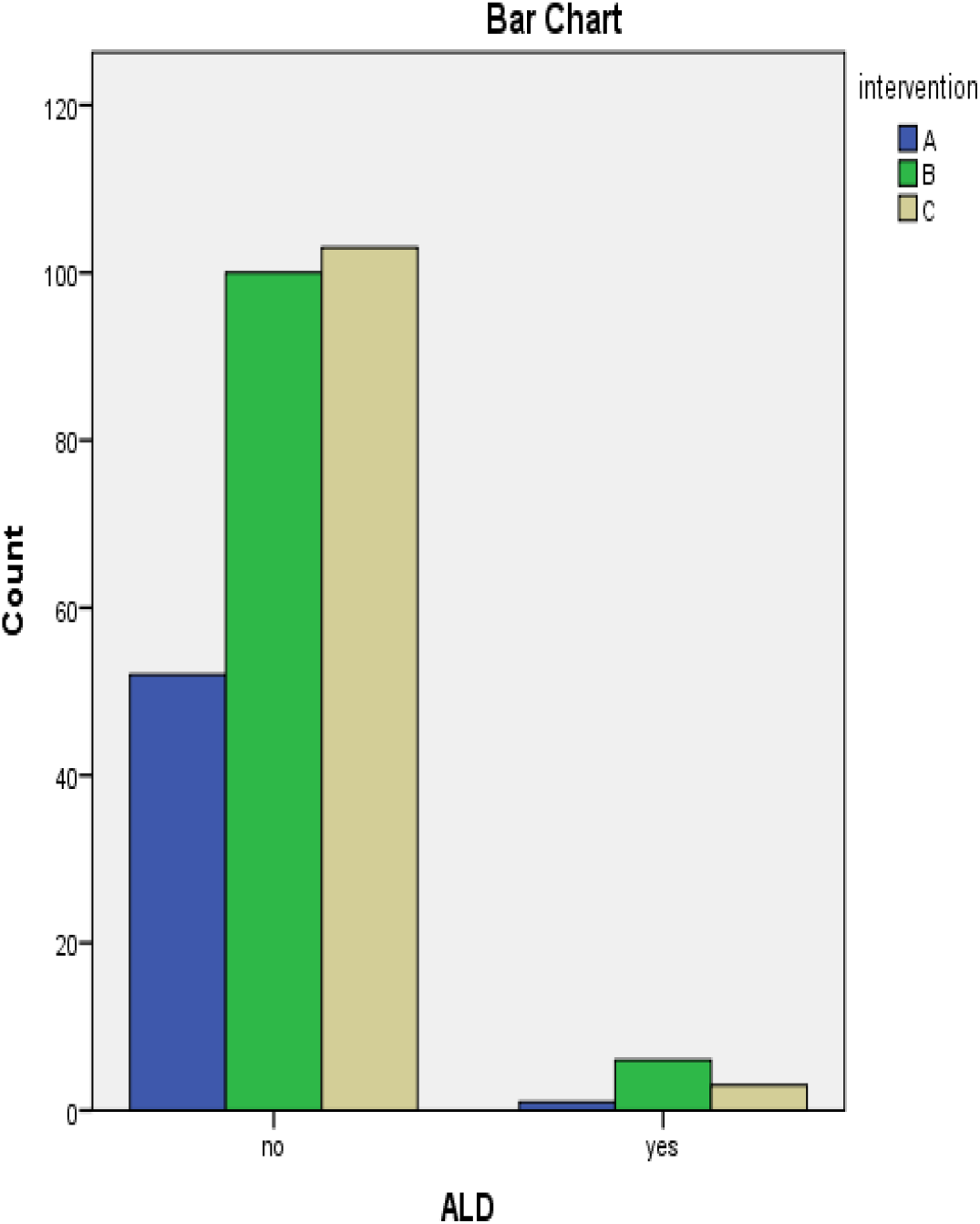
Frequency of ALD incidence across the three groups.

**Figure 63.**
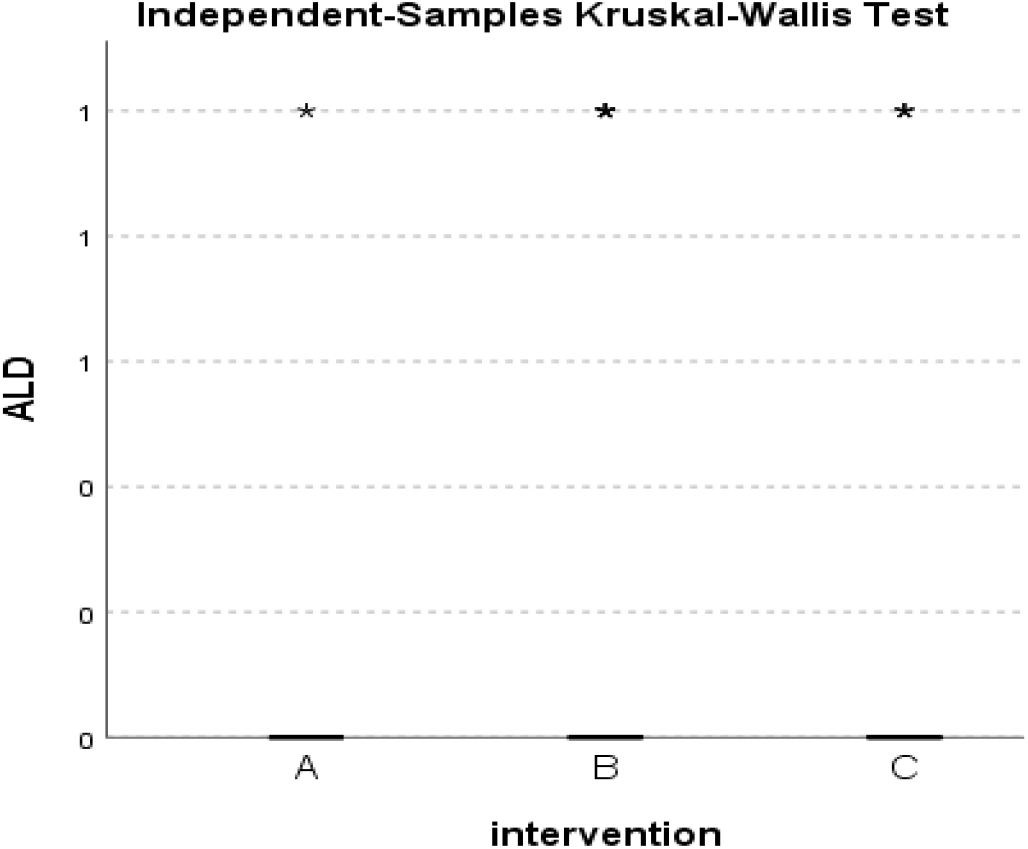
Distribution of ALD incidence across the three groups.

**Figure 64.**
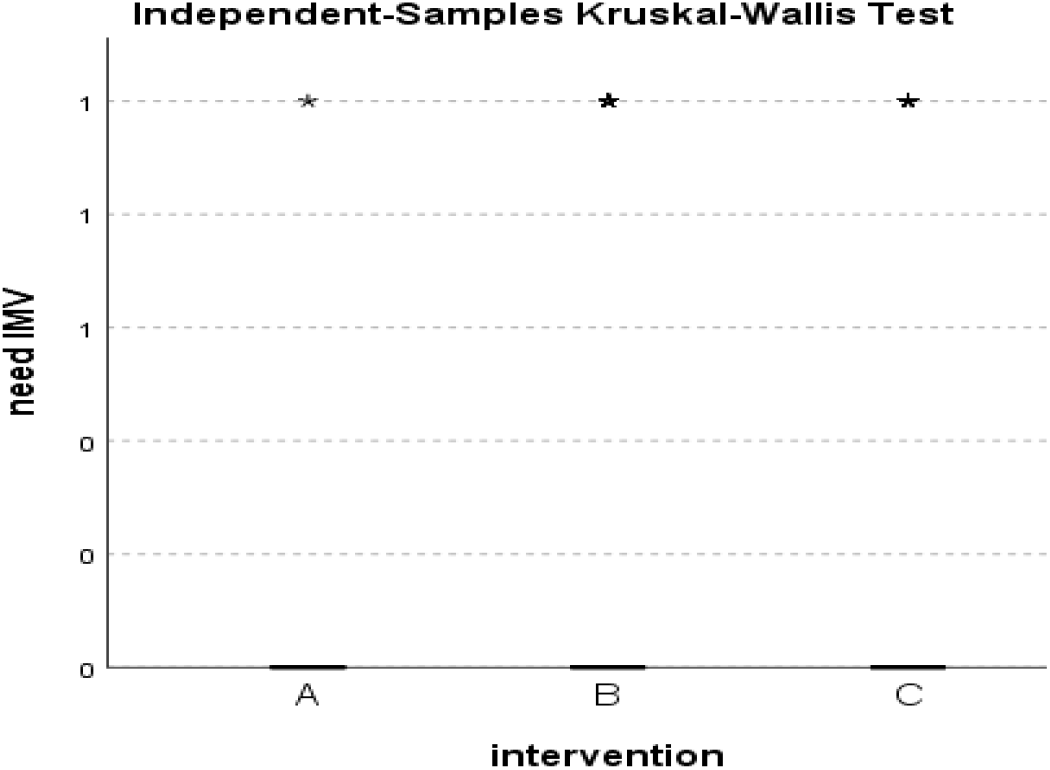
Distribution of IMV need across the three groups.

**Figure 65.**
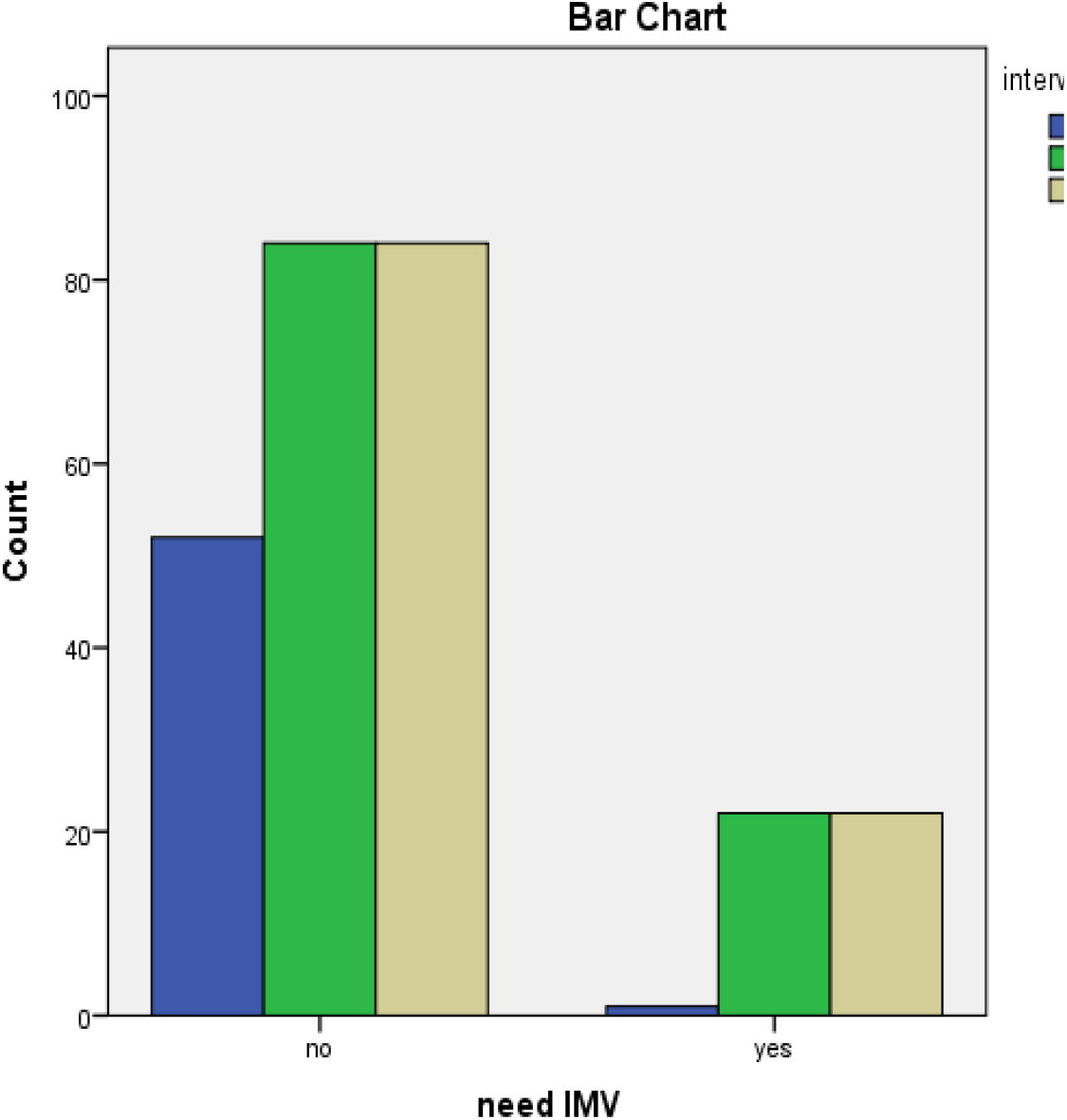
Frequency of IMV need across the three groups.

**Figure 66.**
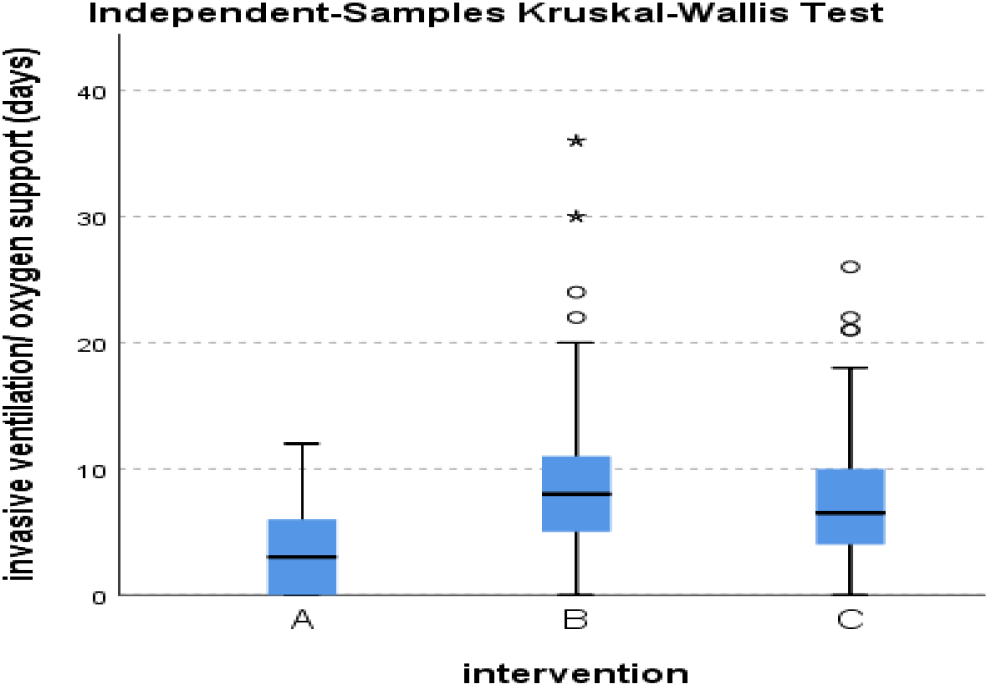
Distribution of O2 therapy and IMV need duration across the three groups.

**Figure 67.**
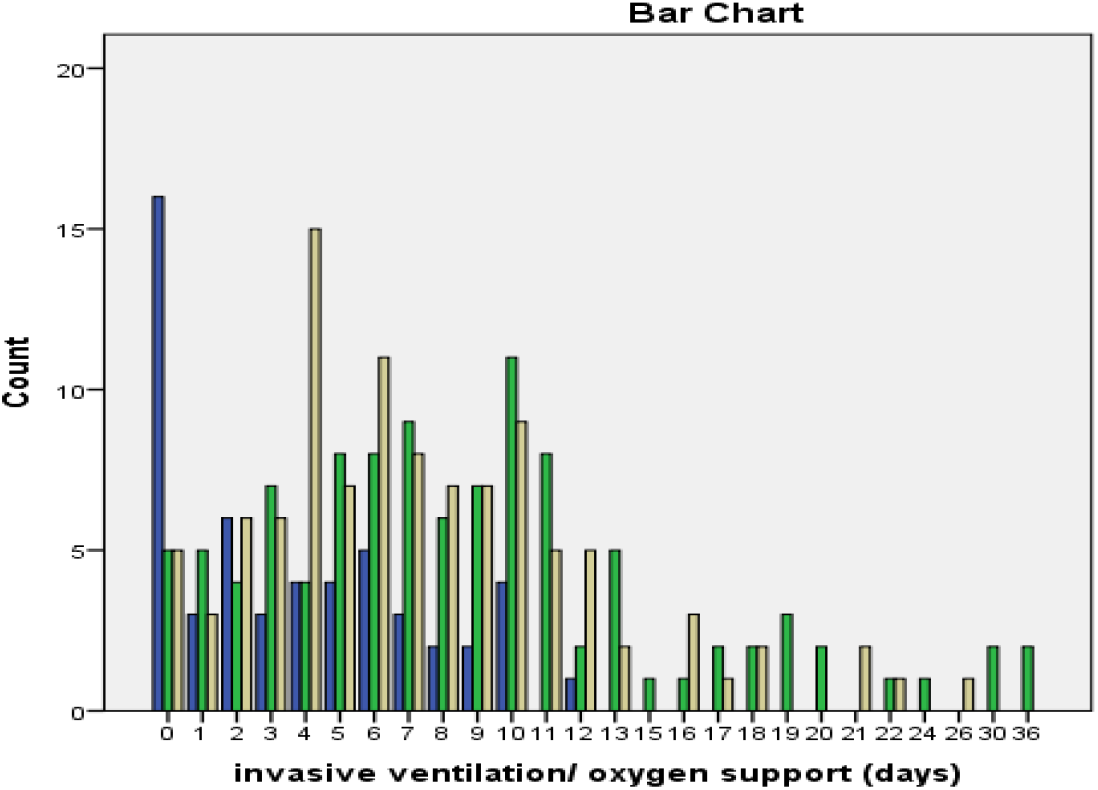
Frequency of O2 therapy and IMV need duration across the three groups.

**Figure 68.**
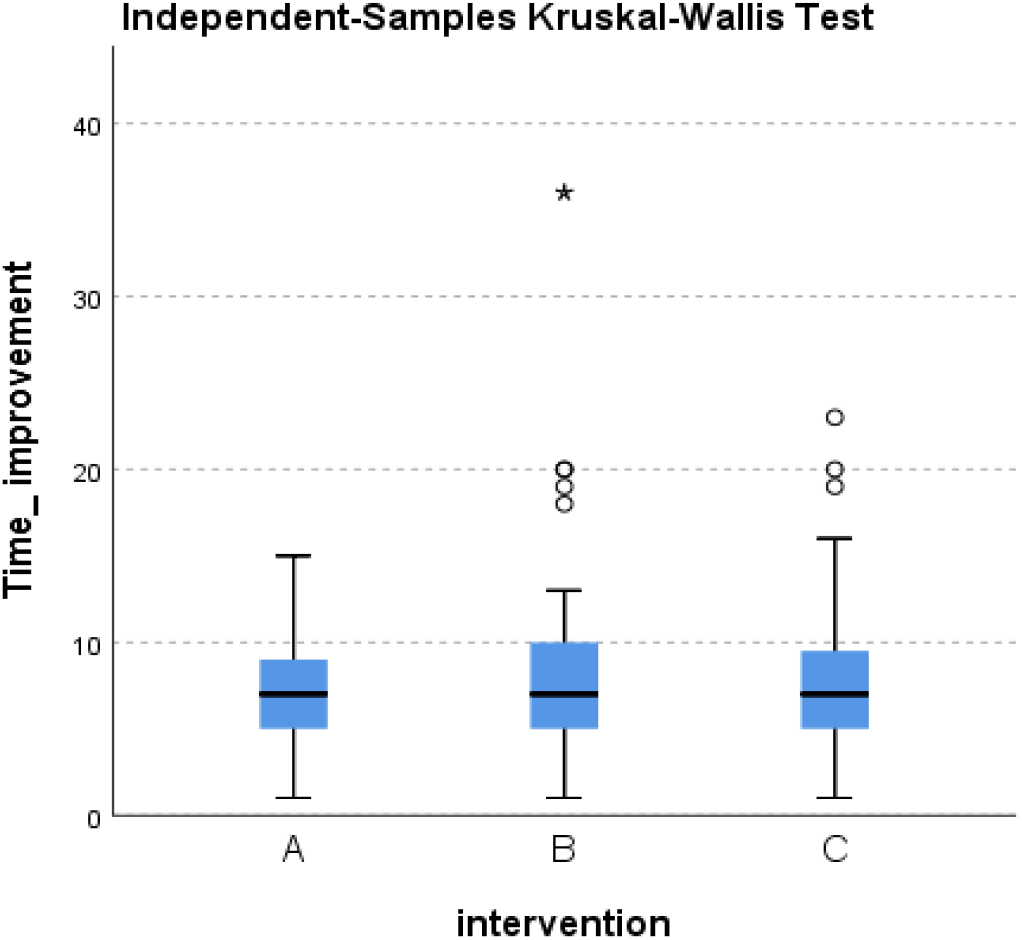
Distribution of Time to clinical improvement across the three groups.

**Figure 69.**
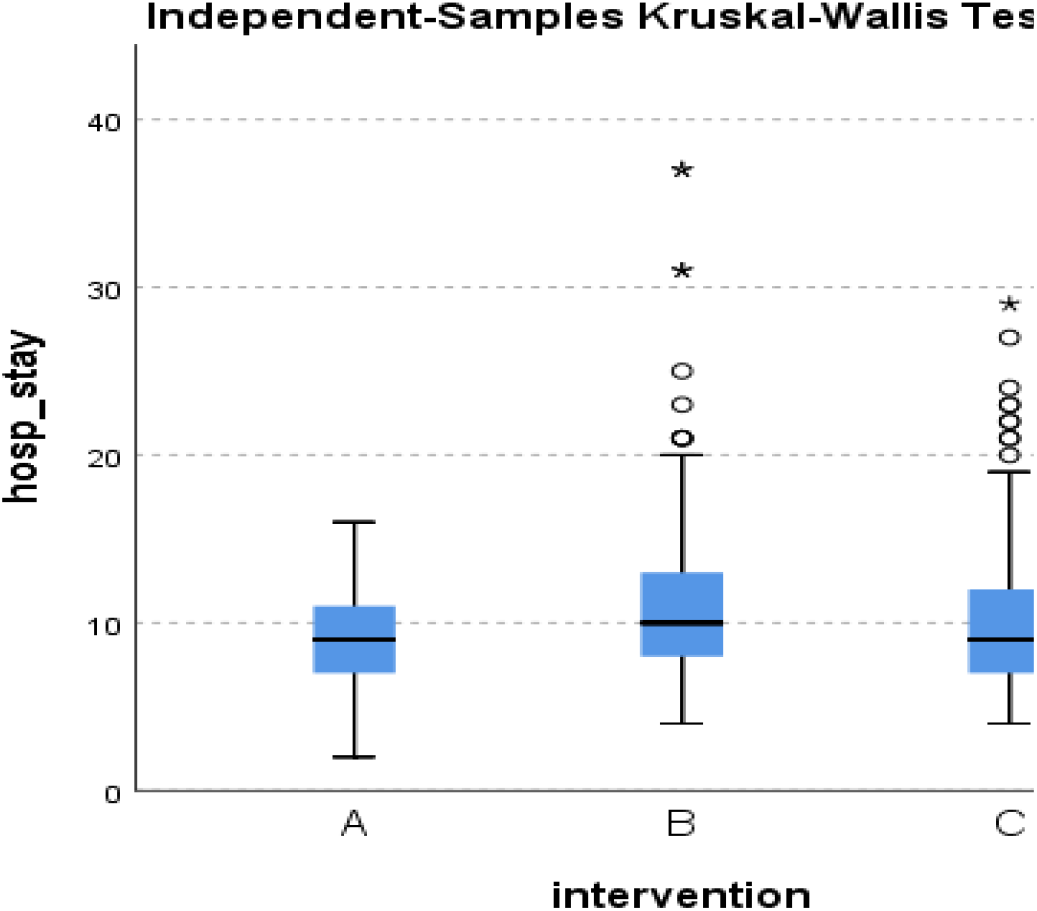
Distribution of hospital stay duration across the three groups.

**Figure 70.**
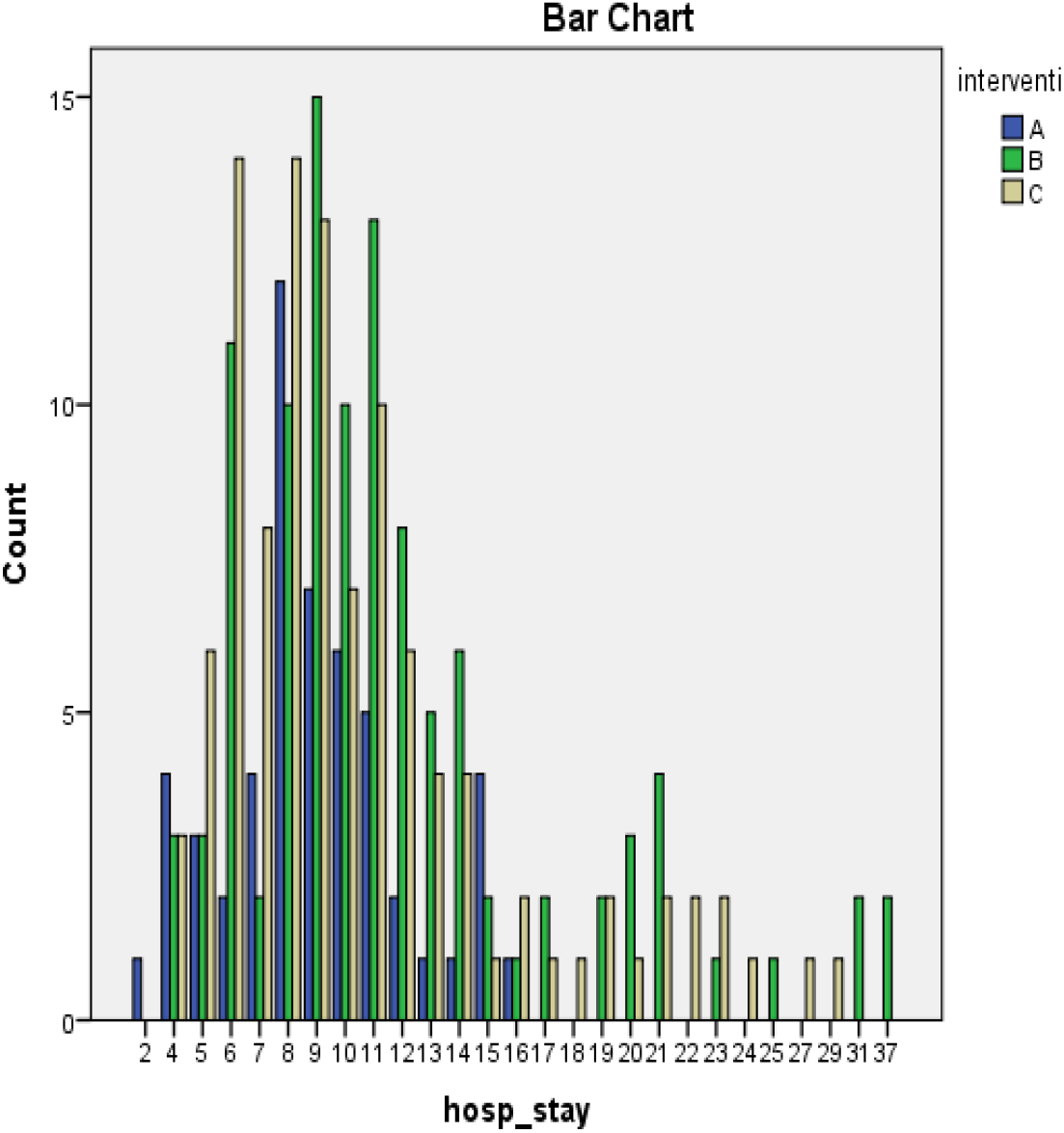
Frequency of hospital stay duration across the three groups.

**Figure 71.**
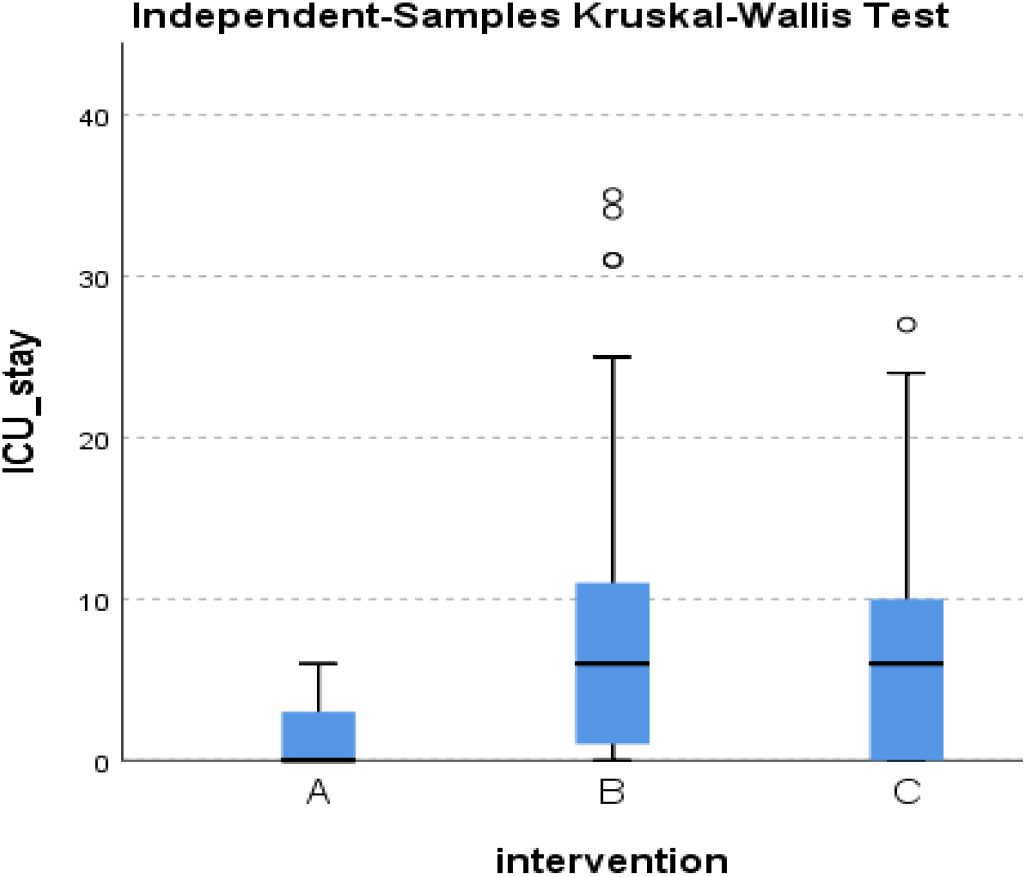
Distribution of ICU stay duration across the three groups.

**Figure 72.**
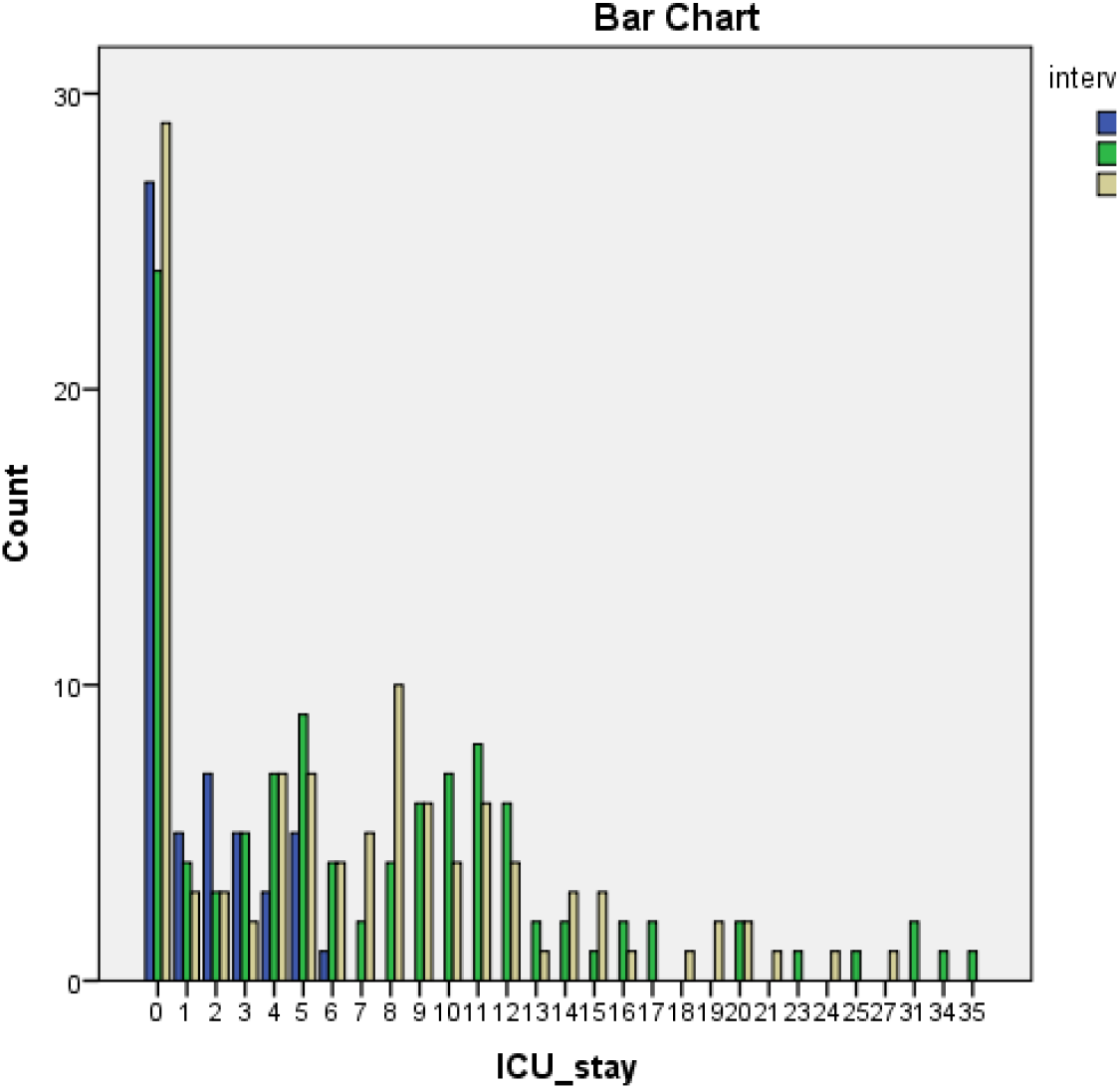
Frequency of ICU stay duration across the three groups.

**Figure 73.**
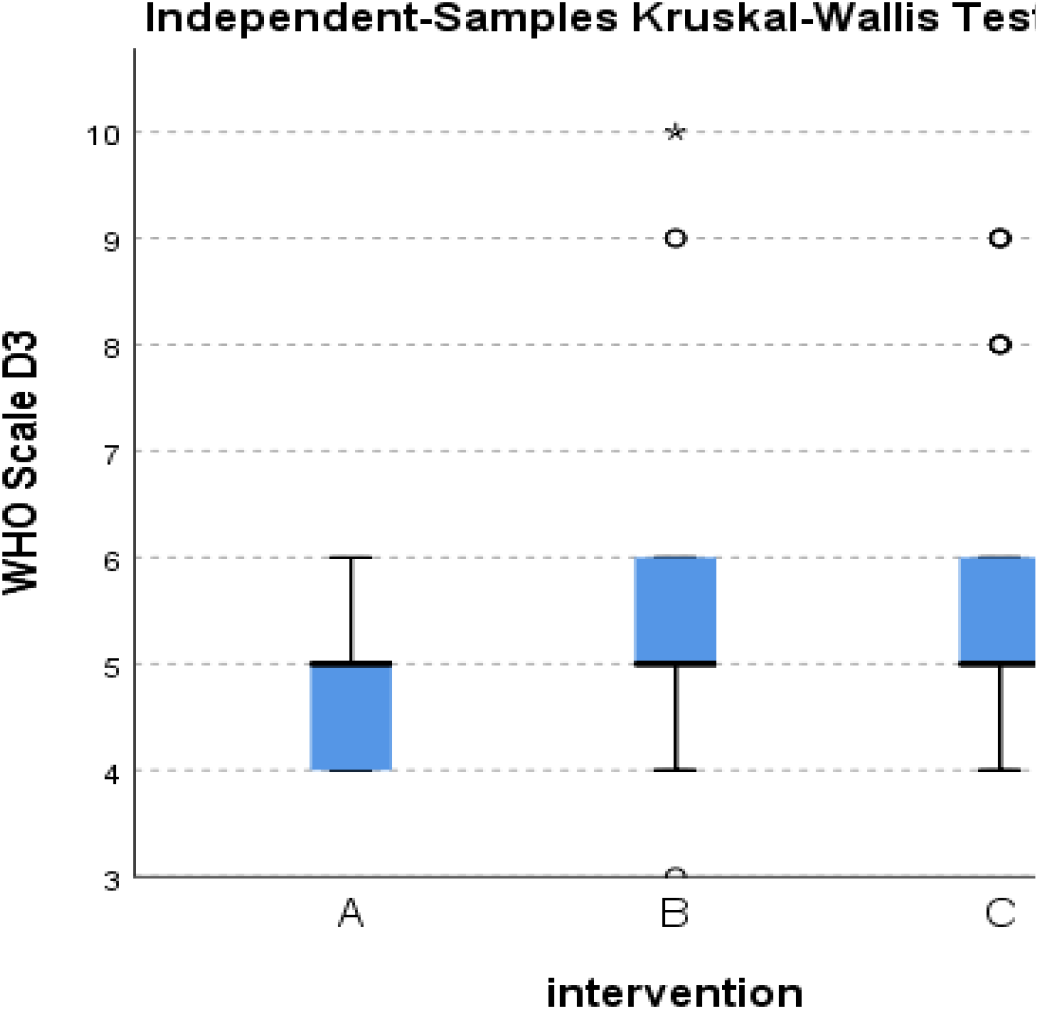
Distribution of WHO score at day 3 across the three groups.

**Figure 74.**
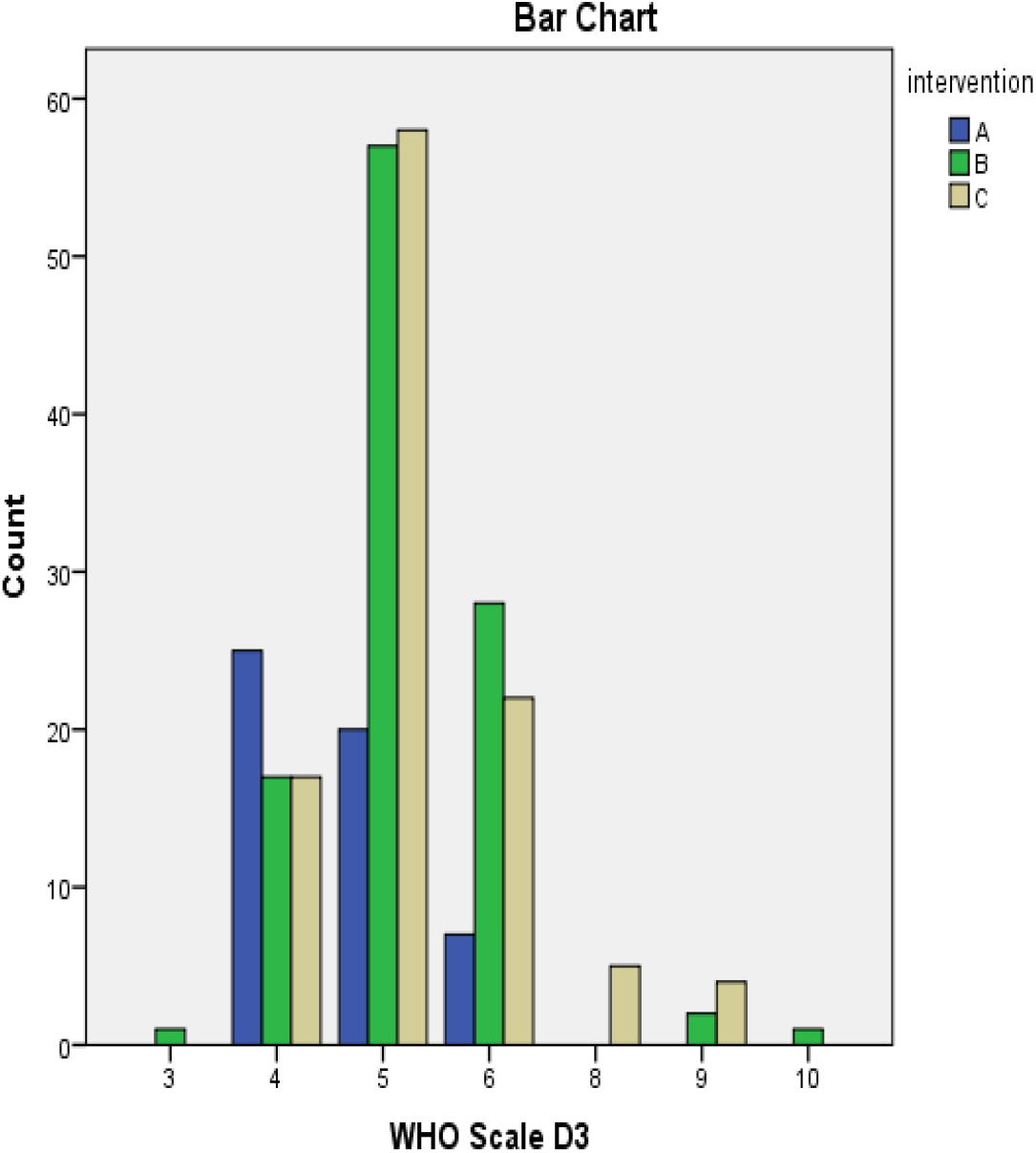
Frequency of WHO score at day 3 across the three groups.

**Figure 75.**
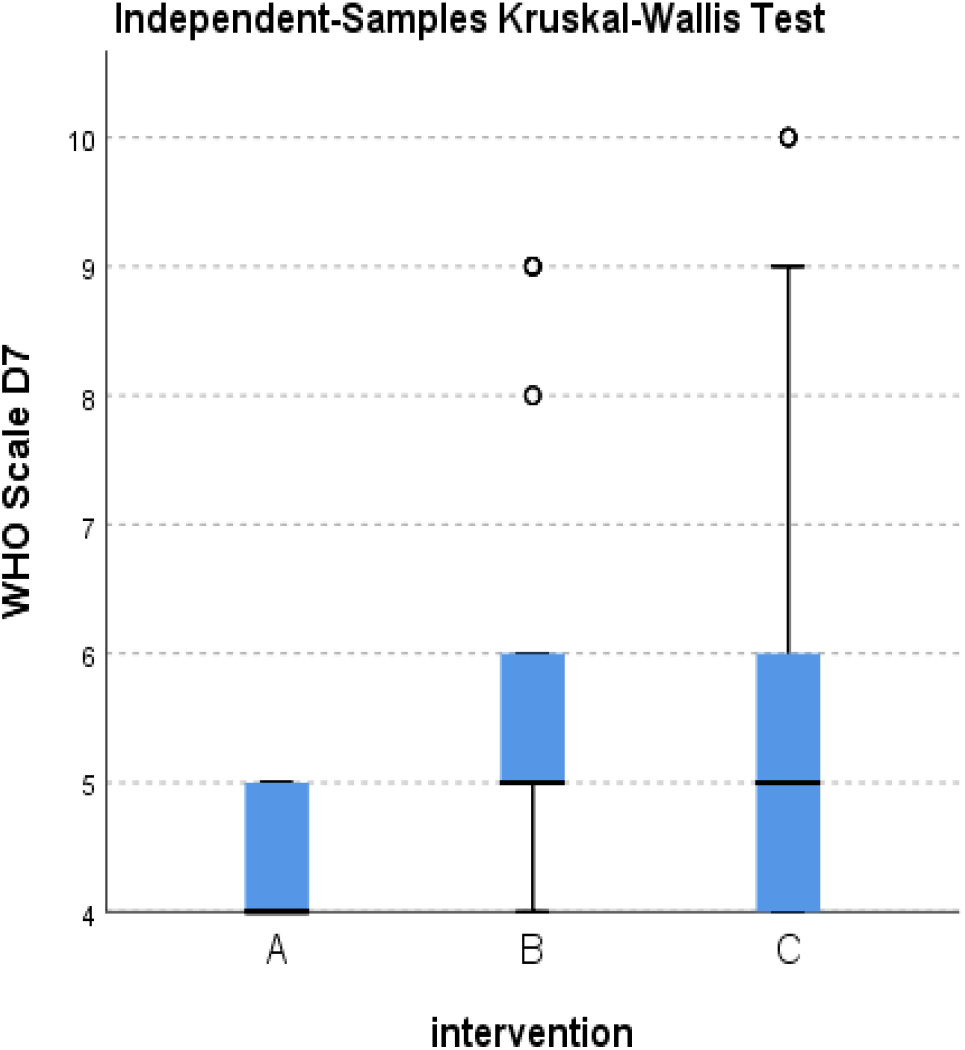
Distribution of WHO score at day 7 across the three groups.

**Figure 76.**
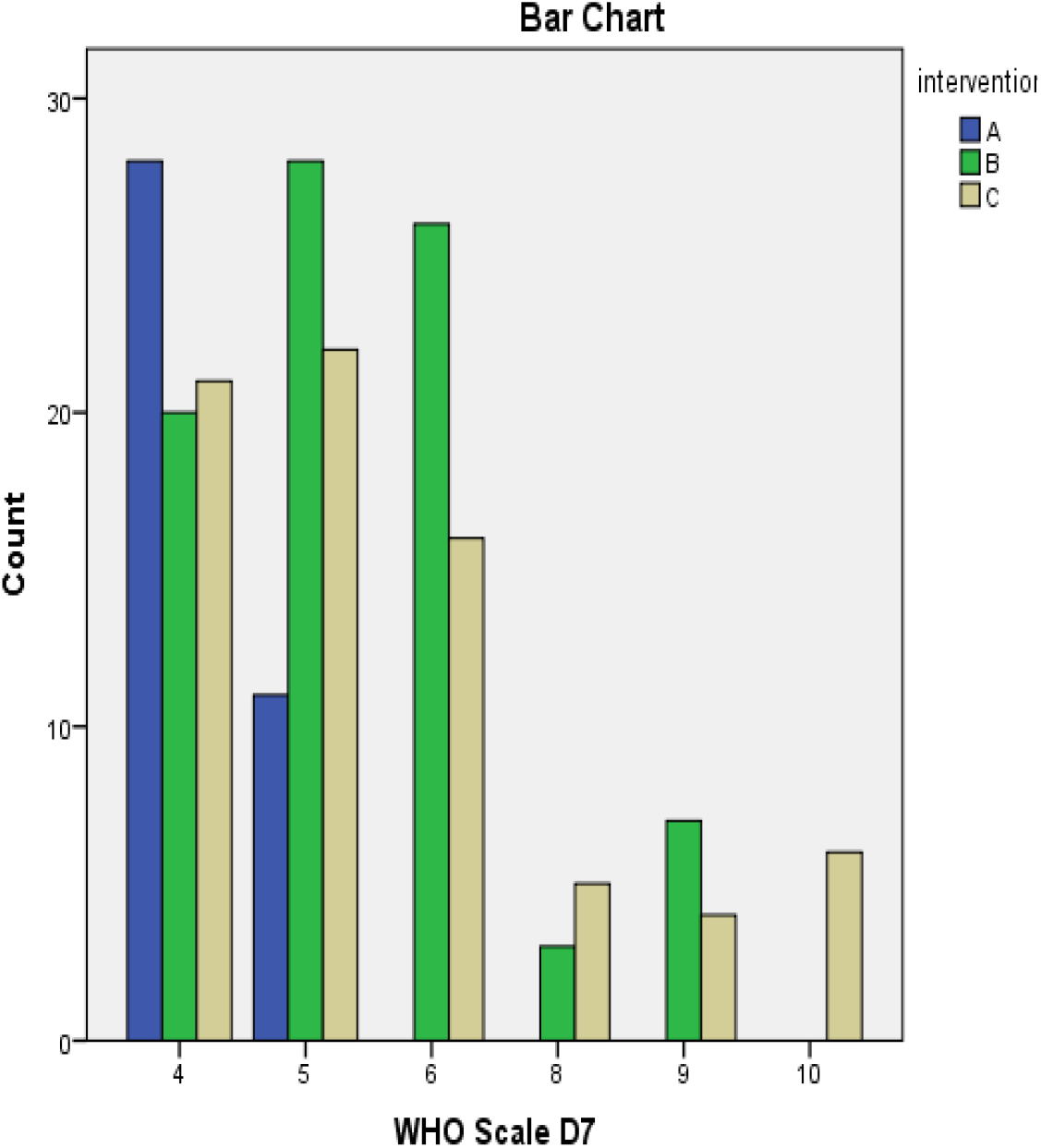
Frequency of WHO score at day 7 across the three groups.

**Figure 77.**
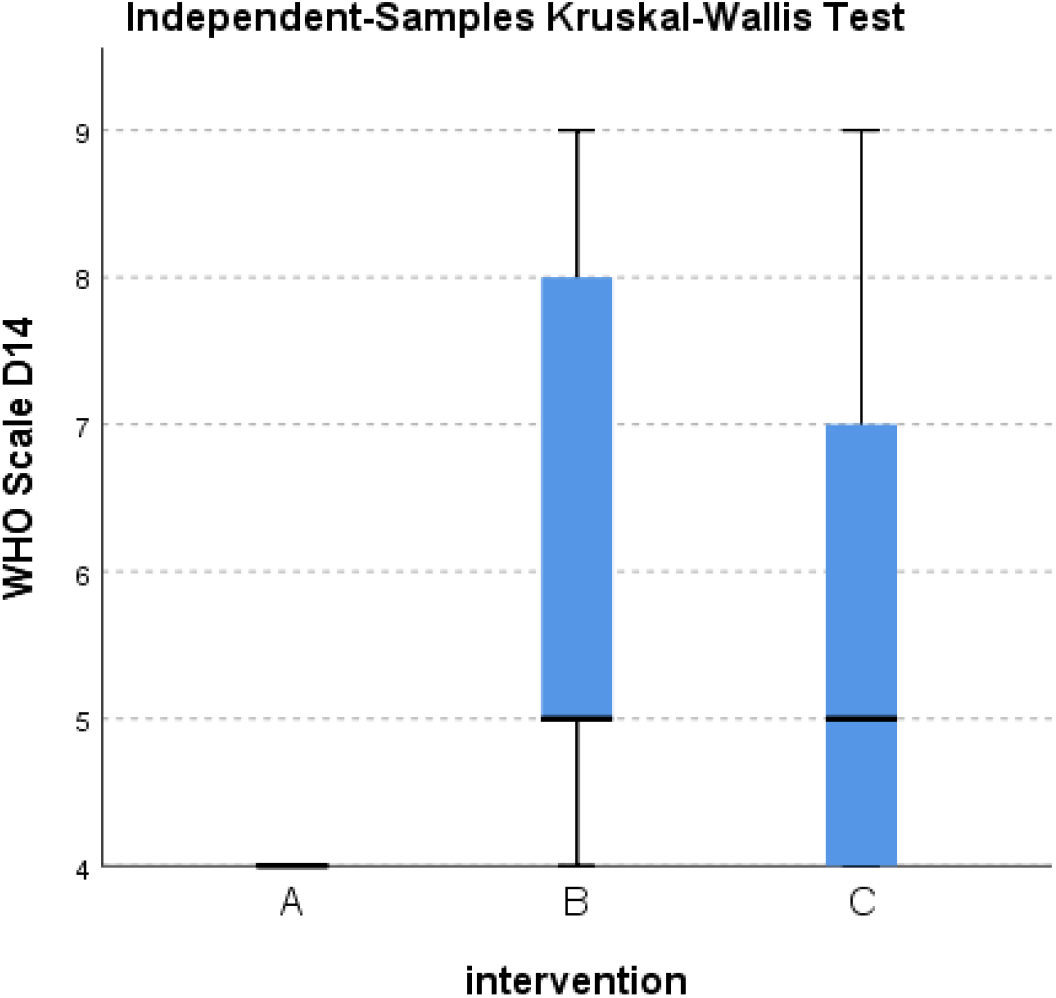
Distribution of WHO score at day 14 across the three groups.

**Figure 78.**
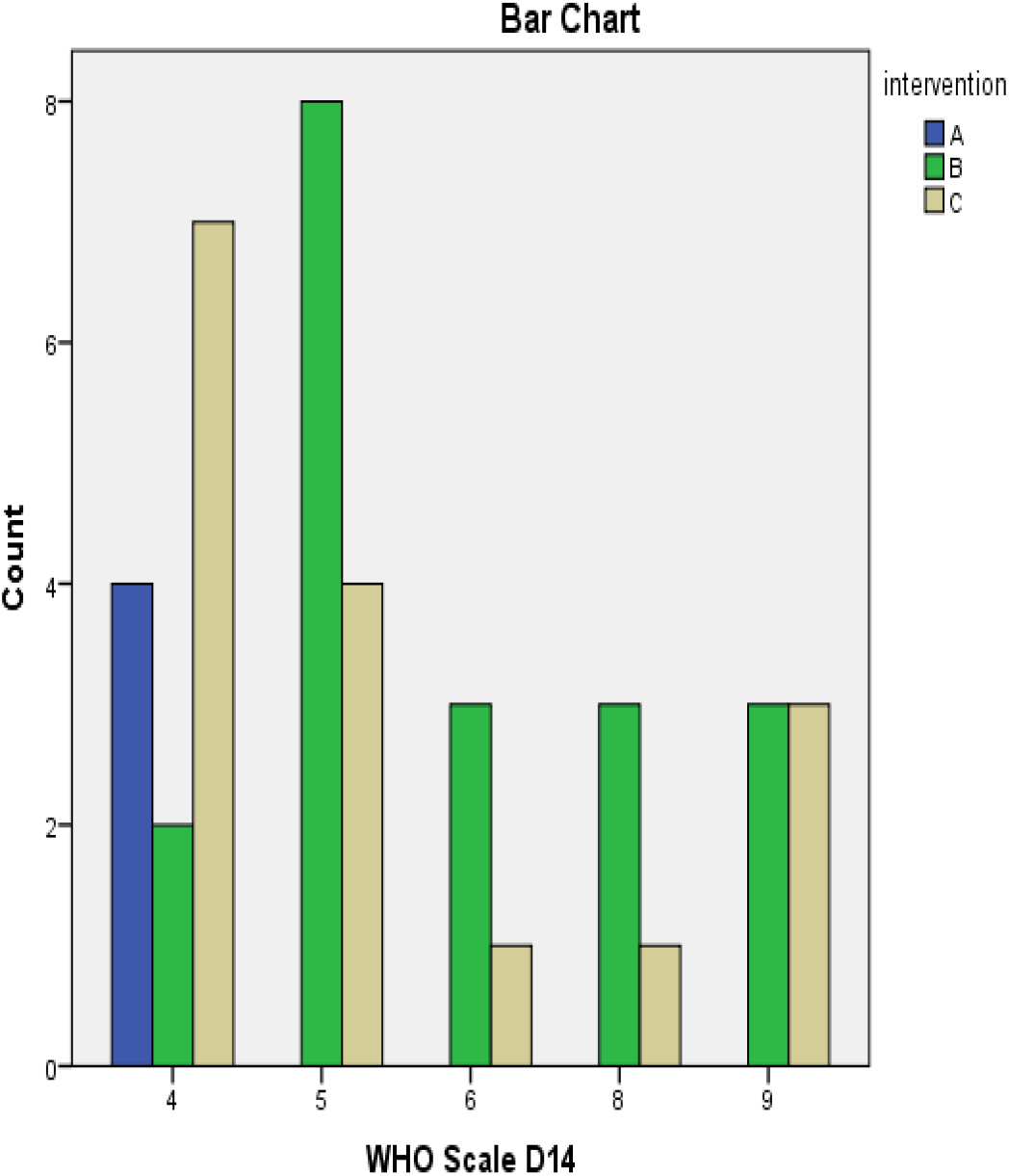
Frequency of WHO score at day 14 across the three groups.

#### 8.2.1. effect of interventions on liver function

there is a statistically non-significant difference between the three groups in liver function tests (AST, ALT, bilirubin) after using the three antivirals with no significant hepatotoxicity in each group.

there is only a statistically significant difference in ALT in day 14 between B-C and in bilirubin in day 7 with albumin in day 14 between A-B & A-C while albumin in day 3,7 between A-C.

#### 8.2.2. effect of interventions on platelet counts

there is only a statistically significant difference between A-C & B-C in platelet count in day 3,7 and there are no other statistically significant differences between groups observed.

#### 8.2.3. effect of interventions on kidney function

there is a statistically significant difference in serum creatinine between A-C & B-C in day 3,7 and between B-C in day 14 and there are no other statistically significant differences between groups observed.

#### 8.2.4. effect on inflammatory markers (CRP, D-dimer, CK, LDH, Ferritin)

There is only a statistically significant difference in CRP at day 3,7, D-dimer, LDH and ferritin at day 7 between A-B & A-C, LDH in day 3 between A-B.

#### 8.2.5. effect on consciousness level

There is a statistically significant difference in GCS in day 3 between A-C & B-C and in day 7 between the three Groups and there are no other statistically significant differences between groups observed.

#### 8.2.6. effect on score of multi-organ functions

There is a statistically significant difference in SOFA score in day 3 between the three group and in day 7,14 between A-B & A-C and there are no other statistically significant differences between groups observed.

#### 8.2.7. effect on oxygen pressure in blood

There is a statistically significant difference in PaO2/FiO2 in day 3,7,14 between A-B & A-C.

#### 8.2.8. effect on 28-day mortality (primary outcome)

There is a statistically significant difference in 28-day mortality between A-B & A-C.

#### 8.2.9. day of death in cases of mortality

There is a statistically significant difference in day of death between A-B & A-C.

#### 8.2.10. mortality at discharge

There is a statistically significant difference in death at discharge between A-B & A-C.

#### 8.2.11. PCR result at hospital discharge (primary outcome)

There is a statistically significant difference in swab PCR result between A-B & B-C.

#### 8.2.12. incidence of any serious adverse effect leading to drug discontinuation (primary outcome)

There is a statistically non-significant difference between the three groups in causing of any serious adverse effect.

#### 8.2.13. incidence of acute kidney injury (AKI) and acute liver damage (ALD)

There is a statistically non-significant difference between the three groups in causing any deterioration on kidneys or liver functions.

#### 8.2.14. need for IMV during hospitalization

There is a statistically significant difference in need for IMV between A-B &A-C.

#### 8.2.15. effect on number of days in which there is need for IMV or O2 therapy

There is a statistically significant difference in number of days with need for IMV or oxygen therapy between A-B & A-C.

#### 8.2.16. effect on time to clinical improvement in days

There is a statistically non-significant difference between the three groups in time to clinical improvement in days

#### 8.2.17. effect on hospital stay duration

There is a statistically significant difference in duration of hospitalization between A-B.

#### 8.2.18. effect on ICU stay duration

There is a statistically significant difference in duration of ICU stay between

A-B & A-C.

#### 8.2.19. effect on WHO scale for COVID cases

There is a statistically significant difference in WHO scale in day 3,7 between A-B & A-C and in day 14 between A-B only.

For more statistical analysis that is performed on clinical data of this study, this is a link to a SPSS output file that contains all statistical analysis of the study. an excel data sheet and a SPSS data file containing all clinical data of the cases of the three groups can be found in this link in addition to an excel data sheet for included and excluded cases with date: https://drive.google.com/drive/folders/1X1dDQwW9vBvusutwMbeebUjN8jJqYxsh?usp=sharing

## IX. DISSCUSION

### 9.1. Regarding baseline characterestics

### 9.1.1. Age

The age in groups A & B is statistically significant less than that in group C.

### 9.1.2. Gender

There is statistically significant more females in group B group than group C.

### 9.1.3. Number of comorbidities

There is statistically significant a greater number of co-morbidities in group C than Group B.

### 9.1.4. Severity of COVID-19

There are statistically significant less severe cases in group A than groups B & C.

### 9.1.5. WHO clinical progression scale

The who scale is statistically significantly lower in group A than groups B & C.

### 9.1.6. Number of symptoms

There is statistically significant a smaller number of symptoms in group A than in groups B & C.

### 9.1.7. Antibiotics use

Although a statistically non-significant difference exists between the three groups in antibiotics use generally, the use of macrolide antibiotics is statistically significant more in group A group than group C.

### 9.1.8. Antiplatelet use

Use of antiplatelet (aspirin) is statistically significant more in group A group than group C.

### 9.1.9. Steroids use

The use of steroids is statistically significant more in group B than group A.

### 9.1.10. Additive therapy use

Use of paracetamol is statistically significant more in group C than group A and use of zinc is statistically significant more in group A than groups B & C.

### 9.1.11. Oxygen therapy use

In general, use of O2 therapy in group A is statistically significant less than groups B & C and O2 therapy using SFM, NIV, IMV in Group A is statistically significant less than Groups B & C, while the use of mask reservoir (MR) as O2 source is more in group B than group C.

### 9.1.12. Vasopressor use

Use of vasopressors in group A is statistically significant less than groups B & C.

### 9.1.13. Oxygen saturation

There is statistically significant more cases in group A who does not need O2 therapy with statistically significant higher O2 saturation on room air than groups B & C.

### 9.1.14. Coagulation profile

PT and INR in group A are statistically significantly less than that in groups B & C.

### 9.1.15. Kidney function test

Serum creatinine is statistically significantly higher in group C than groups A & B.

### 9.1.16. Inflammatory markers

D-dimer is statistically significantly higher in group A followed by group C followed by group B, A>C>B.

### 9.1.17. Blood picture

TLC is statistically significantly higher in group C than group B and hematocrit is statistically significantly higher in group A group than groups B & C.

### 9.1.18. Electrolytes

The potassium level is statistically significantly higher in group C than group B.

### 9.1.19. Blood gases

PaO2 and PaO2/FiO2 are statistically significantly higher in value in group A than group B and PaO2 is statistically significant higher in value in group A than group C.

### 9.1.20. Consciousness level

The consciousness level (GCS) is statistically significantly lower in group C than groups A & B.

### 9.1.21. Multi-Organ Functions Assessment

SOFA score is higher in group C than groups A & B and in group B than group A. A>B>C in multi-organ functions (better multi-organ functions in A than B and B than C)

### 9.2. Regression analysis

After statistical analysis of baseline characteristics of the cases of the three groups and finding that statistically significant differences in some baseline characteristics exist between the three groups. Differences exist between age, gender, number of symptoms, number of co-morbidities, severity of COVID, WHO clinical progression scale, SOFA score, use of antiplatelets, & steroid, & zinc, serum creatinine, PT, INR, TLC, D-dimer, PaO2, PaO2/FiO2 and use of O2 therapy including IMV and NIV.

So, it is necessary to exclude the effect of these variables on the outcomes of the study which represented by the primary outcome and mainly 28-day mortality.

For this reason, regression analysis is performed to explore the effects of these variables on the primary outcome of the study (28-day mortality).

After regression analysis, it is found that all baseline characteristics that differ between the three groups have no effect on the study outcome with exception of need for NIV and IMV, PaO2 and PaO2/FiO2 that show effect on 28-day mortality. This is explained by the need for NIV & IMV and a decline in PaO2 and PaO2/FiO2 can cause an increase in 28-day mortality.

### 9.3. Regarding outcomes of the study after intervention in the three groups

### 9.3.1. effect of interventions on liver function

ALT level is statistically significant higher in group C than group B, bilirubin at day 7 is lower in group A than groups B & C. Albumin in day 14 is higher in group A than groups B & C. while albumin at day 3,7 is higher in group A than group C. From these results, it is concluded that A causes less hepatic damage than B & C.

### 9.3.2. effect of interventions on platelet counts

Platelets count at day 3,7 is statistically significantly lower in group C than groups A & B.

### 9.3.3. effect of interventions on kidney function

Serum creatinine at day 3,7 level is statistically significant higher in group C than groups A & B and at day 14 is higher in group C than group B.

### 9.3.4. effect on inflammatory markers (CRP, D-dimer, CK, LDH, Ferritin)

CRP at day 3,7 and D-dimer LDH & ferritin at day 7 are statistically significant lower in group A than groups B & C. LDH at day 3 is statistically significant lower in group A than group B.

From these results, it is concluded that inflammatory marker levels have been lowered by A than B & C interventions.

### 9.3.5. effect on consciousness level

GCS at day 3 is statistically significant lower in C than groups A & B, while GCS at day 7 is statistically significant higher in group A than groups B & C and in group B than group C(A>B>C).

### 9.3.6. effect on score of multi-organ functions

SOFA score at day 3 is statistically significant higher in group C than groups A & B and in group B than group A, SOFA score at day 7,14 is statistically significant lower in group A than groups B & C.

From these results, it is concluded that the best multi-organ functions are in this arrangement (A>B>C) with group A has the best multi-organ functions (lowest SOFA score).

### 9.3.7. effect on oxygen pressure in blood

PaO2/FiO2 value at day 3,7,14 is statistically significant higher in group A than groups B & C.

From these results, it is concluded that group A has more favorable oxygen level in blood than groups B & C.

### 9.3.8. effect on mortality at day 28 (primary outcome)

Group A has statistically significant lower 28-day mortality rate than groups B & C.

### 9.3.10. day of death in cases of mortality

Day of death is statistically significant less in group A than Groups B & C.

### 9.3.11. mortality at discharge

In addition to lower 28-day mortality, a statistically significant lower mortality rate at hospital discharge with group A than groups B & C.

### 9.3.12. PCR result at hospital discharge (primary outcome)

Group B has a statistically significant greater number of positive cases at discharge than groups A & C and group C has more positive cases than group A which has the highest number of negative cases at discharge followed by group C followed by group B.

### 9.3.13. incidence of any serious adverse effect leading to drug discontinuation (primary outcome)

All three interventions have no significant adverse effect that proves their safety.

### 9.3.14. incidence of AKI and hepatotoxicity

No nephrotoxicity or hepatotoxicity observed with all three interventions that prove their safety.

### 9.3.15. need for invasive mechanical ventilation (IMV) during hospitalization

Group A has a statistically significantly lower need for IMV than groups B & C.

### 9.3.16. effect on number of days in which there is need for IMV or oxygen therapy

Group A has statistically significantly less duration with need for O2 therapy or IMV than groups B & C.

### 9.3.17. effect on time to clinical improvement in days

A statistically non-significant difference exists between the three groups in time to clinical improvement in days.

### 9.3.18. effect on hospital stay duration

Group A has statistically significantly less duration of hospitalization than group B.

### 9.3.19. effect on ICU stay duration

Group A has statistically significant less duration of ICU stays than Groups B & C.

### 9.3.20. effect on WHO scale for COVID cases

Group A has statistically significant lower WHO progression scale than Groups B & C at day 3,7 and statistically significant lower WHO progression scale than Group B at day 14 that proves that less progression of the cases in Group A (lower WHO scale) than Groups B & C.

## X. CONCLUSION

Casirivimab and imdevimab group achieves less 28-day mortality rate, less mortality at hospital discharge, more negative swab cases, less need for O2 therapy and IMV, less duration of this need, less hospital and ICU stay, less case progression as presented by lower WHO scale and better multi-organ functions as presented by lower SOFA score than Remdesivir and Favipravir groups.

From all of these results, it is concluded that group A (Casirivimab & imdevimab) has more favorable clinical outcomes than groups B (remdesivir) & C (favipravir).

## Data Availability

All data produced in the present work are contained in the manuscript

https://drive.google.com/drive/folders/1X1dDQwW9vBvusutwMbeebUjN8jJqYxsh?usp=sharing

